# Impact of rare and common genetic variation on cell type-specific gene expression in human blood

**DOI:** 10.1101/2025.03.20.25324352

**Authors:** Anna S.E. Cuomo, Eleanor Spenceley, Hope A. Tanudisastro, Blake Bowen, Albert Henry, Hao Lawrence Huang, Angli Xue, Wei Zhou, Matthew J. Welland, Samantha J. Bryen, Oscar A. Dong, Arthur S. Lee, Jeffrey M. Pullin, Kristof Wing, Owen Tang, Michael P. Gray, Michael Franklin, Michael Harper, Michael Silk, Katalina Bobowik, Alexander Stuckey, John Marshall, Vivian Bakiris, Caitlin Uren, Bindu Swapna Madala, Amy Miniter, Caitlin Bartie, Drew R. Neavin, Zhen Qiao, Eyal Ben-David, Ling Chen, Kyle Kai-How Farh, Stuart M. Grieve, Tung Nguyen, Chris Wallace, Jennifer Piscionere, Owen M. Siggs, Hannah Nicholas, Katrina M. de Lange, Alex W. Hewitt, Gemma A. Figtree, Daniel G. MacArthur, Joseph E. Powell

## Abstract

Understanding the genetic basis of gene expression can shed light on the regulatory mechanisms underlying complex traits and diseases. Single cell-resolved measures of RNA levels and single-cell expression quantitative trait loci (sc-eQTLs) have revealed genetic regulation that drives sub-tissue cell states and types across diverse human tissues. Here, we describe the first phase of TenK10K, the largest-to-date dataset of matched whole-genome sequencing (WGS) and single-cell RNA-sequencing (scRNA-seq). We leverage scRNA-seq data from over 5 million cells across 28 immune cell types, and matched WGS, from 1,925 individuals, which provides power to detect associations between rare and low-frequency genetic variants that have largely been uncharacterised in their impact on cell-specific gene expression. We map the effects of both common and rare variants in a cell type-specific manner using a recently introduced method that increases power by modelling single cells directly rather than relying on aggregated ‘pseudobulk’ counts. We identify putative common regulatory variants for 83% of all 21,404 genes tested and cumulative rare variant signals for 47% of genes. We explore how genetic effects vary across cell type and state spectra, develop a framework to determine the degree to which sc-eQTLs are cell type-specific, and show that about half of the effects are observed only in one or a few cell types. By integrating our results with functional annotations and disease information, we also further characterise the likely molecular modes of action for many disease-variant associations. Finally, we explore the effects that genetic variants have on gene expression across continuous cell states and functions, and effects that vary cell state abundance directly.

## Introduction

Genome-wide association studies (GWASs) have revealed hundreds of thousands of association signals between genetic variants and human diseases and phenotypic traits^1^, but the modes of action by which most of these variants affect human disease and phenotypes remain to be characterised, especially for variants outside protein-coding regions. The characterisation of the impact of genetic variants on gene expression – known as expression quantitative trait locus (eQTL) mapping – has already uncovered molecular mechanisms for many of these variants by linking them to the putative genes that they regulate^2–10^.

eQTL mapping can help identify the genes and pathways involved in disease pathogenesis, which is a critical step in identifying opportunities for therapeutic intervention^11^. However, important limitations remain. First, eQTL mapping has traditionally been limited to common genetic variation (population minor allele frequency [MAF] >1-5%) due to relatively small sample sizes and the widespread use of genotyping arrays, which capture variants in linkage disequilibrium (LD) with known common genetic variants, but do not accurately genotype most rare genetic variants and a substantial fraction of complex structural variants^12^. Generating a more complete picture of the impact of genetic variation on human molecular phenotypes requires the use of whole genome sequencing approaches.

Second, eQTLs are extremely context-specific, with multiple studies demonstrating that genetic variants frequently impact gene expression levels in ways that can only be detected in specific tissues^3^, sorted cell types^7,13–16^, or upon stimulation^17–19^. The recent introduction of single cell-resolved measures of RNA levels allows expression phenotypes to be measured in these more granular cell types and states, and single-cell eQTL mapping has demonstrated that the effects of many genetic variants – including disease-associated ones – on gene expression can only be detected when mapping eQTLs within specific cell types and even transient cellular states^20–36^.

To overcome these challenges, we introduce phase 1 of the TenK10K (up to 10,000 cells for 10,000 individuals) study. In total TenK10K phase 1 encompasses 5,920,025 cells from 2,103 individuals, from which we selected 5.4 million cells from 1,925 individuals of European ancestry to deeply explore the impact of rare and common genetic variation on gene expression phenotypes. In accompanying papers, we leverage the same dataset to investigate the impact of tandem repeat variation on gene expression [Tanudisastro *et al*. 2025]^37^ and the causal links between gene expression measurements and human diseases and traits [Henry, Senabouth *et al*. 2025]^38^, and an overlapping set of samples to explore the impact of genetic variation on chromatin accessibility [Xue *et al*. 2025]^39^.

Here we use an association method we recently developed, SAIGE-QTL^40^, to map single-cell expression quantitative trait loci across cell types for both common and rare variants called from whole genome sequencing (WGS) data and identify over 150,000 common variant and over 30,000 rare variant effects on gene expression, including common or rare variant signals for nearly 5,000 genes (∼25%) with no previously reported eQTLs in the largest blood eQTL study to date^2^. We propose a framework for quantifying the extent of cell type specificity or sharedness of reported eQTLs and show that about half of the effects are found in only one or few cell types, and almost none are ubiquitously found across all. We perform statistical colocalisation between our eQTLs and 60 disease phenotypes and blood traits, and identify over 60,000 colocalisation events for over 5,000 genes, providing novel potential molecular mechanisms in almost 900 genes associated with human phenotypic variation. We use a novel biologically-driven deep learning method, scDeepID, to identify continuous cell states that relate to cell function and map over 15,000 eQTLs that vary dynamically along those states. Finally, we identify 164 independent genetic variants that regulate the abundance of cell states.

## Results

### High throughput blood scRNA-seq and matched WGS data of 1,925 individuals

After quality control, a total of 5,438,679 peripheral mononuclear blood cells (PBMCs) from 1,925 donors were considered for analysis (**Fig. 1, Supplementary Table 1; Methods**). We collected and sequenced PBMCs from all individuals using the high-throughput 3’ sequencing kit from 10x Genomics, achieving 2,825 cells on average per individual after quality control (**Supplementary Figures 1, 2**; **Methods**).

**Figure 1.**
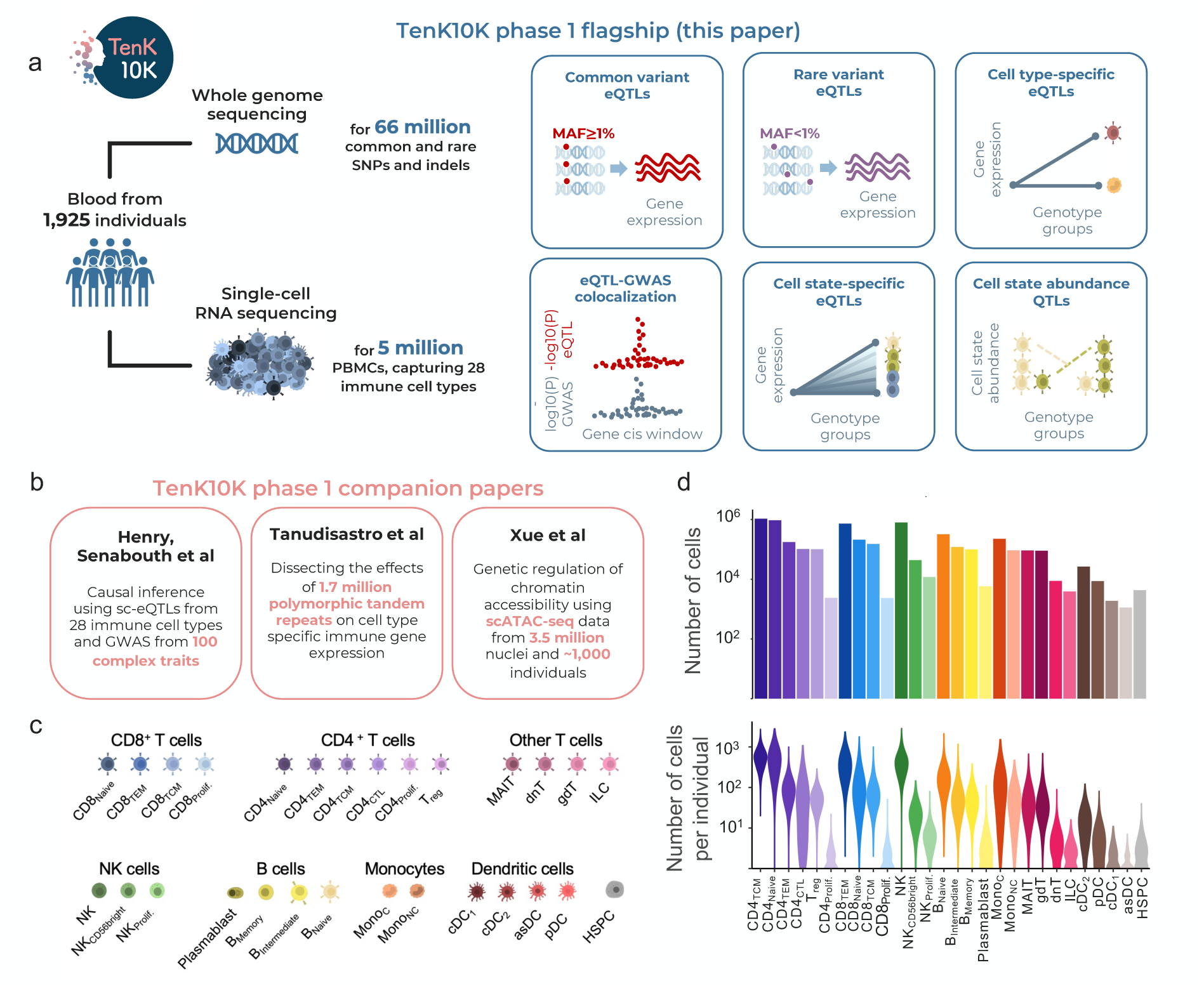
Population-scale single-cell RNA sequencing and matched whole-genome sequencing from 1,925 individuals identified 28 immune cell types and 66 million variants. **a)** Overview of the data used in this paper introducing phase 1 of the TenK10K cohort. High quality matched whole-genome sequencing (WGS) and single-cell RNA sequencing (scRNA-seq) data were generated for 1,925 individuals of European ancestry (after QC). After computational analysis and processing, we conducted both common and rare variant eQTL analysis and identified cell type– and state-specific effects of genetic regulation in immune cells. We also performed colocalisation analysis with GWAS from 60 traits and diseases and mapped QTLs impacting cell state abundance (csaQTLs). **b)** Accompanying manuscripts perform complementary analyses using the TenK10K phase 1 cohort. Briefly, Henry*, Senabouth* *et al*. integrate sc-eQTLs from the present study and 100 GWAS studies to elucidate the causal role of gene expression in immune cell types across complex traits (*=co-first authors); Tanudisastro *et al*. use the WGS data to generate a catalogue of >1.7 million polymorphic tandem repeats and dissect their effects on single-cell resolved gene expression; Xue *et al*. combine common genetic variants called from WGS with scATAC-seq generated for a subset of ∼1,000 individuals to investigate the genetic regulation of chromatin accessibility at cellular resolution. **c)** Schematic of the 28 immune cell types identified (and abbreviations used throughout the manuscript), organised by major cell type. **d)** The number of cells available for analysis by cell type for the 28 immune cell types is summarised in d. Top: bar plot displaying total cell numbers per cell type on a log 10 scale, bottom: violin plots displaying the distribution across individuals on a log_10_ scale.

Each cell was assigned to their donor of origin via demultiplexing, and to an immune cell type using a combined approach of reference-based classification and hierarchical clustering as recommended by the single-cell eQTLGen Consortium^41^ (**Methods**). Briefly, we applied a supervised classification using a reference panel of protein markers from Hao *et al.*^42^ to assign cells to distinct lineages, including myeloid and lymphoid, with the latter further classified into B, natural killer (NK), and CD4+ and CD8+ T cells, and the former into monocytes and dendritic cells. Next, we used unsupervised clustering to split cells into sub-cell types further. Each cell was assigned to one of 28 different cell types, spanning the key expected immune lineages (**Fig. 1c, Supplementary Fig. 3, Supplementary Table 2**).

Additionally, we generated WGS data for all 1,925 individuals, which was processed and quality controlled using established pipelines (**Methods**). We called a total of 66,658,242 sites between single nucleotide polymorphisms (SNPs) and indels, of which 53,784,018 (80.6%) were rare (MAF<1%), approximately 4.5 times as many as called from SNP array data (34.4 compared to 4.6 million when considering SNPs only for 960 individuals with matched data; **Supplementary Fig. 4; Methods**); and 4,317,810 (6.5%) were low-frequency (1%≤MAF<5%).

### Single-cell eQTL mapping reveals common variant impact on gene expression

To assess the effects of common genetic variation on gene expression changes, we mapped *cis* eQTLs, testing for the effects of genotypes at common (MAF>1%) variant sites within a window of +/-100kb around the gene body on the single-cell expression profile of the target gene (testing all genes expressed in at least 1% of all cells for a given cell type; **Methods**). We used SAIGE-QTL^40^ to map eQTL effects using single-cell profiles, modelled as Poisson counts, rather than relying on ‘pseudobulk’ aggregation. We mapped eQTLs separately in each of the 28 cell types and confirmed our Type I error was controlled using permutations (**Fig. 2a**). At FDR<5% after multiple testing correction, we identified a total of 154,932 eQTLs in at least one of the 28 cell types from 17,674 unique eGenes (83% of all genes tested; **Fig. 2b** and **Supplementary Fig. 5**). The number of eQTLs identified varied substantially across cell types, largely due to the number of cells available for each cell type and the resulting number of individuals tested affecting statistical power, as previously shown^27,40^ (**Fig. 2b, Supplementary Fig. 6, Supplementary Table 2**).

**Figure 2.**
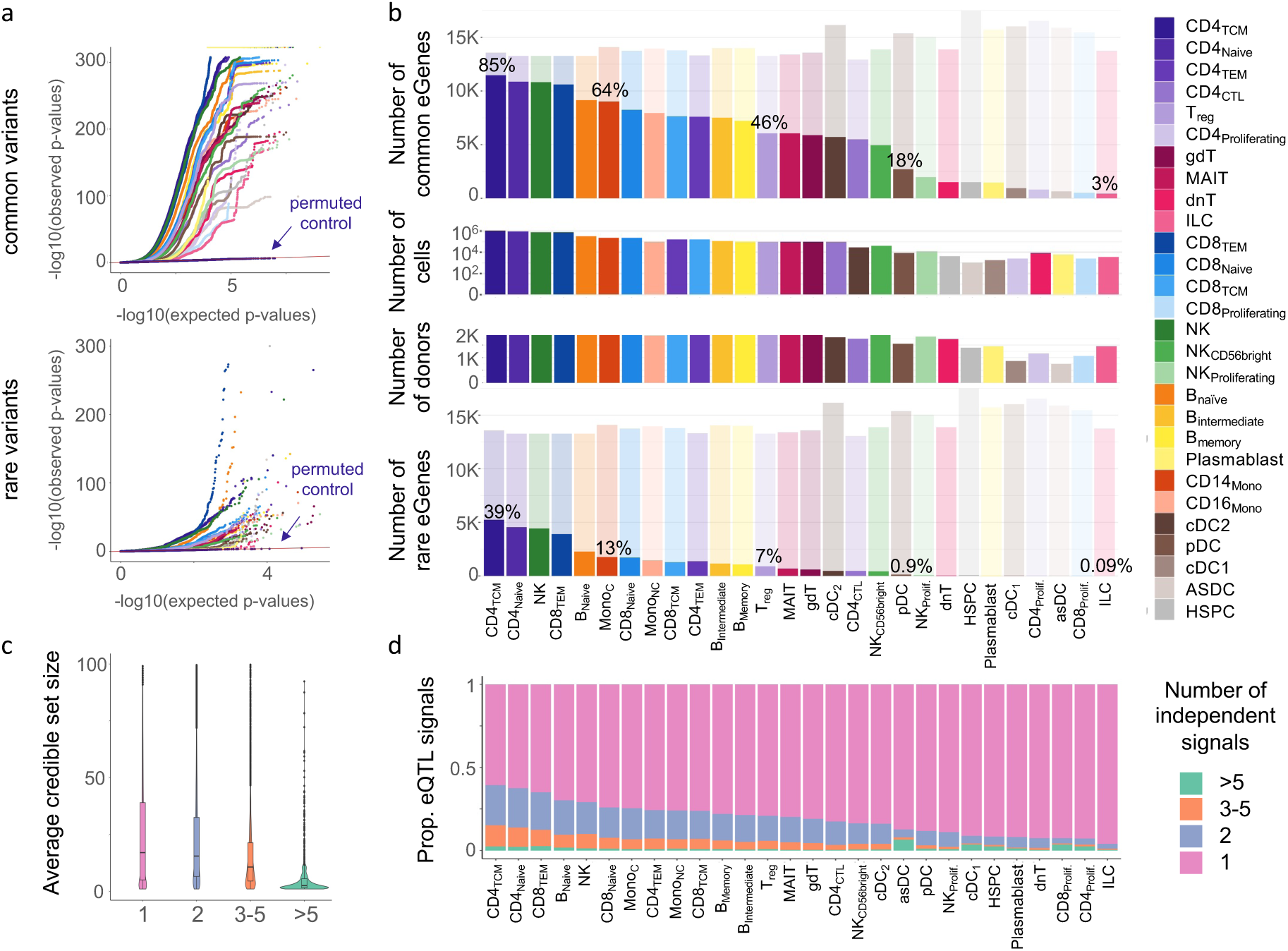
Common and rare eQTL mapping in 28 immune cell types. **a)** Quantile-quantile (QQ) plot for all common (MAF≥1%, top) and rare (MAF<1%, bottom) variant eQTL results across all 28 immune cell types. For the most abundant cell type, CD4_TCM_, we shuffled individual IDs and re-performed the test to assess calibration (labelled ‘permuted control’). Colour represents cell type, as shown in the legend in the top right. **b)** Number of eGenes identified (solid colour) compared to the total number of genes tested (faded) across cell types. Top: common variant eQTL mapping (from single variant tests), bottom: rare variant eGenes using combined *p*-values of the burden and SKAT tests. The middle bar plots represent the number of cells and individuals available for each cell type to demonstrate the former’s (and to some extent the latter’s) key role in the power differences across cell types. For representative cell types, the percentage of eGenes compared to the total number of genes tested is highlighted. Percentages for all cell types are available at **Supplementary Table 2**. **c)** Distribution of SuSie credible set sizes stratified by number of non-primary signals (divided into four bins). **d)** Proportion of primary and non-primary eQTLs identified in each cell type via fine-mapping using SuSie, coloured by the number of independent signals divided into bins as in **c**.

eQTL variants were concentrated around the transcription start and end sites (TSS and TES, respectively) and declined in number proportionally to the distance from the gene body, likely indicating their role in genetic regulation and similarly to what has been shown previously^2^. Indeed, we found that 83% of all 154,932 lead eQTL variants were located within 50kb of either side of the gene, and 94% within +/-80kb.

In order to identify additional independent signals for our eGenes, we performed fine-mapping using SuSie^43^ (**Methods**). This analysis revealed an additional 24,925 (a 16% increase) non-primary (secondary, tertiary, etc.)^44,45^ eQTL signals for 7,131 eGenes (40.4%). We used 95% credible sets from SuSie to quantify numbers of independent signals for each eGene, and explored differences between primary effects (all variants in the credible set containing the top significant variant for each eGene) and non-primary effects (all other credible sets). We find that primary effects contain larger credible sets (**Fig. 2c**), and genes in the higher-proportion cell types have substantially more non-primary effects (**Fig. 2d**). As shown by others^45^, non-primary signals were found on average to be further from the TSS, and more likely in enhancer as opposed to promoter regions (**Supplementary Fig. 7**).

We compared the eGenes we identified with those identified by bulk eQTLs in blood. First, we considered the eQTLGen consortium study^2^ the largest eQTL study to date, which focuses on whole-blood. We identified a comparable number of eGenes compared to eQTLGen (17,674 and 16,987 respectively), despite the 15-fold smaller sample size (1,925 compared to over 30,000 total individuals; **Supplementary Fig. 8a**). Over a quarter of significant eGenes in our study were not identified in eQTLGen (*n*=4,572, 25.9%); those genes were slightly more lowly expressed (**Supplementary Fig. 8b**), and their expression was more cell type-specific (**Supplementary Fig. 8c**) compared to eGenes found by both studies; TenK10K-unique eGenes were also more likely to be eGenes in fewer cell types overall (**Supplementary Fig. 8d**), in rare cell types (*n*<60,000) compared to more abundant ones (**Supplementary Fig. 8e**), or in one cell type only (‘cell type-specific’, **Supplementary Fig. 8f**), demonstrating the importance of mapping eQTLs at cellular resolution. On the other hand, 23% of eGenes identified in eQTLGen were not detected in our study: these genes were extremely lowly expressed across all 28 cell types (average expression 4% as opposed to 30% for shared eGenes and 28% for TenK10K-only eGenes; **Supplementary Fig. 8b**).

An alternative to sc-eQTL mapping to detect cell type-specific eQTLs is to use bulk eQTL mapping on sorted cell types, and many studies have been shown to be extremely well powered, even with modest sample sizes. Here, we compared our eQTLs to four bulk studies (BLUEPRINT^7^, CEDAR^15^, Kasela *et al*^14^, Schmiedel *et al*^13^) using sorted immune cell types, matching cell types like-for-like. Overall, we demonstrate similar to much-increased power to identify cell type-specific eQTLs across cell types and studies, demonstrating the usefulness of single-cell eQTL mapping for this task. Importantly, our study offers assessment of more granular cell types (*n*=28) than previous studies using sorted immune cell types (*e.g.,* 13 cell types in Schmiedel *et al*; **Supplementary Fig. 9**).

We compared our results to the OneK1K study^17^, which was the previously largest sc-eQTL map in blood. We identified nearly three times more eGenes compared to OneK1K (17,674 compared to 6,469). This is due to a number of factors, including the increased power from both larger sample size (about twice the number of individuals: 1,925 compared to 982), the number of cells per individual (2,825 compared to 1,291), the consequent more granular cell type resolution (28 compared to 14 cell types), and the increased power due to modelling single-cell expression profiles rather than relying on pseudobulk. The eGenes found in our study and not OneK1K were more lowly expressed and more likely to be expressed in fewer cell type populations (in our data; **Supplementary Fig. 10**).

Finally, we compared the eQTLs identified separately in each cell type to a new set of eQTLs obtained from pseudobulk expression values aggregated across all of our 28 cell types to quantify the improvement provided by cell type resolution within our own data (**Methods**). At FDR<5%, the all-cell types pseudobulk eQTL analysis identified 11,359 eGenes, similar to the number of eGenes found in our most abundant cell type, CD4_TCM_, and nearly 30% less than when we consider all eGenes identified in at least one of the cell types (**Supplementary Fig. 11**).

We correlated our identified eQTL effect sizes with scores from PromoterAI (a convolutional neural network that predicts promoter activity of variants, their target gene and their effect on gene expression^46^; **Methods**) for the lead variant for each gene across the different cell types, finding good agreement (median correlation=0.66; **Supplementary Fig. 12**), demonstrating concordance of both the magnitude and direction of effects.

### Mapping the effects of rare variation on single-cell gene expression

Most bulk and single-cell eQTL studies to date^3,6,22,27,28^ have focused on common genetic variants (population minor allele frequency; MAF≥ 1-5%) due to their relatively small sample size and the use of array-based genotyping with imputation. Studies investigating the effects of rare variants (MAF < 1%) on tissue-level gene expression^4,47^ have conducted “outlier analyses”, where rare variants are sought specifically in individuals with extreme expression levels for a given gene rather than performing systematic assessments of the effects of *cis* rare variants on expression levels genome-wide across individuals.

By combining whole-genome sequencing and single-cell data at population scale, TenK10K phase 1 is uniquely positioned to assess the effects of rare genetic variation on cell type-specific expression genome-wide. Using SAIGE-QTL, we mapped the effects of rare variation (MAF<1%) within a *cis* window (+/-100kb) around each gene’s body, using gene-set based tests for increased power (**Methods**). For each cell type, we tested for the effects of local rare variants on all sufficiently expressed genes (genes expressed in at least 1% of all cells from the cell type of interest) using two different types of tests designed to capture different regulatory mechanisms: the burden test, and the sequence kernel association test (SKAT)^48^.

Overall, we observe 34,376 rare variant signals across all cell types for 10,005 unique rare eGenes (47% of all genes tested) after FDR correction on the combined *p*-values from the two tests (burden test and SKAT; FDR<5%, **Fig. 2b, Supplementary Fig. 13; Methods**). Type I error was well controlled for the rare variant tests (**Fig. 2a),** and the number of cells for each cell type was highly correlated with the number of eGenes identified (correlation=0.98, **Supplementary Fig. 6**).

Most eGenes were identified by the SKAT test alone (29,260; 85%), followed by both SKAT and burden (4,975; 14.5%) and burden test alone (141; 0.5%). This is consistent with SKAT being the more appropriate test when variants in the set have mixed direction of effects (*i.e.,* some increasing gene expression, some decreasing it) which we expect in a regulatory context such as this (**Fig. 3a**).

**Figure 3.**
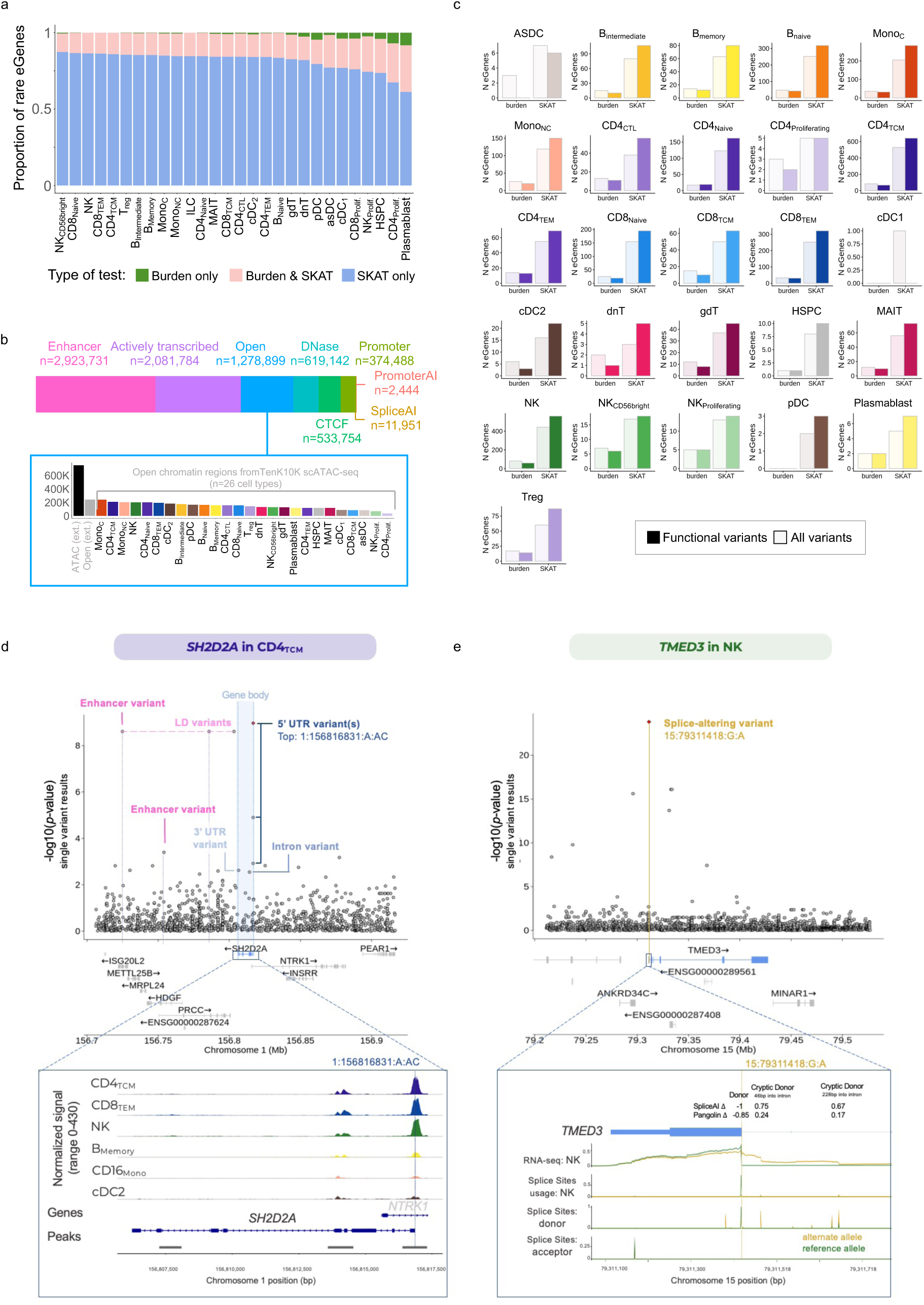
Rare variation shapes cell type-specific gene expression in blood. a) Bar plot showing the proportion of rare eGenes identified using the different set-based tests – identified as eGenes at FDR<5% by the burden test, the SKAT test, or both – across cell types. **b)** Breakdown of number of rare variants overlapping with the functional annotations used. If a variant was annotated to be in more than one category, it is counted for both. The bottom panel splits the “open chromatin” annotation further into external resources and cell type-specific open chromatin regions based on TenK10K scATAC-seq (more information on the publicly available functional annotations used are in **Supplementary Table 3**). **c)** Number of eGenes identified at *p*-value < 10^−6^ identified when testing for association using only functionally annotated (solid colour) or all *cis* (faded colour) rare variants (**Methods**). Each plot represents a different cell type (*n*=26), results for both SKAT and burden are shown. **d)** Locus zoom plot of rare genetic variants associated with the *SH2D2A* gene on chromosome 1 in CD4_TCM_, with the leading genetic variants annotated. “LD variants” are variants in perfect LD with the annotated putatively causal ones. Bottom plot: zoom in on the chromatin accessibility profile across representative cell types for each major group around the *SH2D2A* gene and with the top variant (1:156816831:A:AC) shown as a vertical line. **e)** Locus zoom plot of rare genetic variants associated with the *TMED3* gene on chromosome 15 in NK, with the leading splice-altering genetic variant (15:79311418:G:A) annotated. Bottom plot shows AlphaGenome-predicted RNA-seq coverage, splice sites usage, and splice sites tracks for exon 1 and 2 of the *TMED3* gene for the 15:79311418:G:A variant (ALT, yellow) and reference (REF, green) alleles. Tracks are predicted for natural killer cells (CL:0000623). SpliceAI and Pangolin^62^ splice site prediction scores are annotated for the canonical donor splice site and two cryptic donors 46 and 228 bp into intron 1. LD: linkage disequilibrium; UTR: untranslated region.

When comparing eGenes with a rare variant signal to those with at least one common variant eQTL, we found that rare eGenes were more ubiquitously expressed across all cell types, whereas common eGenes were expressed in a more cell type-specific manner (**Supplementary Fig. 14a**), although this may be due at least in part to power (**Supplementary Fig. 15**). Single-variant test results show the expected relationship between allele frequency and effect sizes with rarer cell types displaying larger effects on gene expression (**Supplementary Fig. 14b**). Both common and rare eGenes were enriched for putative promoter activity as estimated by PromoterAI^46^, with stronger effect variants being enriched at rare allele frequencies (**Supplementary Fig. 14c**).

### Rare variant effects on cell type-specific gene expression reveal novel regulation

While common variant effects on gene expression have been well characterised, rare variation remains less well studied, especially in the non-coding genome. To further characterise our rare variant signals, we overlapped our signals with a combined set of functional annotations from public datasets to characterise genomic regions as putative enhancers, promoters, transcription factor binding sites, DNase sites and others, as well as splice-altering and promoter regions based on SpliceAI^49^ and PromoterAI^46^ scores respectively., Finally, we included cell type-matched open chromatin regions using scATAC-seq for a subset of our samples [Xue *et al*.] (**Fig. 3b, Supplementary Fig. 16, Supplementary Table 3; Methods**).

We performed a second set of gene-level tests (SKAT and burden), focusing only on variants predicted to be either in cell type-matched open chromatin regions or in regulatory regions (promoter or enhancer regions, splice-altering sites, etc; **Methods**). Despite reducing the number of variants included in the tests by more than half, these tests resulted in a greater number of eGenes on average, across cell types (1.2 times as many eGenes on average, and a larger number of 23 of the 26 cell types tested when considering the SKAT test and dTSS weights, **Fig. 3c**; **Supplementary Fig. 16**). This was especially true when weighting variants based on their distance from the gene’s transcription start site (dTSS, the most powerful approach in the SAIGE-QTL publication^40^) likely reflecting the proximity of regulatory regions to the gene, and less pronounced when using equal weights. In contrast, when up-weighting rarer variants using the *Beta*(1,25) distribution, the all-*cis* variant test performed slightly better than the functional-only one (for 24 out of 16 cell types, 1.09 as many eGenes when using all variants instead of functionally annotated ones, **Supplementary Fig. 17**). We hypothesise that this may be due to the very rare variants that do affect gene expression being mostly coding and thus excluded from our regulatory-focused functionally annotated regions.

As an example, we identified the significant downregulation of *SH2D2A* in CD4_TCM_ (Cauchy combined *p*-value=1.48×10^−2^) with the effects driven by several rare variants located in functional regions, including three 5’ UTR variants, one 3’ UTR variant, three putative enhancer variants (likely on a single haplotype) and one intron variant (**Fig. 3d**). None of these variants have been previously reported to affect the expression of *SH2D2A,* likely due to their low frequency (MAF< 0.005), highlighting the value of whole genome sequencing. The top 5’ UTR variant (1:156816831:A:AC, rs561132719) is an indel which additionally overlapped with an open chromatin peak in T and NK cells in the 5’ UTR region of *SH2D2A* using scATAC-seq data generated for a subset of the individuals from our cohort (**Fig. 3d, Supplementary Fig. 18; Methods**). The signal is stronger upon conditioning on the top common variant for the same gene with an opposing direction of effect (conditional *p*-value=2.6×10^−4^). *SH2D2A* encodes the T cell-specific adaptor protein (TSAd) that induces cytokine production in CD4+ T cells specifically, and has been linked to autoimmune conditions including hypothyroidism^50–52^.

We also found that the expression of *PRDM4* was downregulated by the combined effects of multiple rare variants in most T cell subpopulations, most significantly in CD4+ naive T cells (CD4_Naïve_; *p*-value=1.2×10^−17^). While one of these variants (12:107695720:A:G, rs34017446) was previously reported (in whole blood, with consistent direction of effect^2^), we report two novel rare(r) variants here for the first time. We hypothesise that one variant (12:107761146:T:G, rs71454708; MAF=0.0099) in high LD with 12:107695720:A:G is likely causal, as it overlaps a CTCF binding site and likely promoter region according to multiple sources (Encode^53–55^, chromHMM^56^, UCSF TF-ChIP data^57^), had high activity-by-contact (ABC^58,59^) score and was predicted to be in a promoter region and to result in expression downregulation (PromoterAI score=-0.77; **Supplementary Fig. 19**). *PRDM4* encodes a transcription factor that mediates cell proliferation and has been linked to a number of cancers^60^.

In some cases, we identified rare variants that likely affect pre-mRNA splicing. For example, we found an association between 15:79311418:G:A (rs141194056) and *TMED3* in several cell types (most significantly in NK, *p*-value=1.6×10^−24^). The 15:79311418:G:A (NM_007364.4:c.168+1G>A) variant is predicted by SpliceAI and AlphaGenome^61^ to abolish the canonical donor splice site in intron 1 of the *TMED3* gene and increase the strength of two intronic cryptic donors. Use of either cryptic donors would lead to a premature termination codon (PTC) and nonsense mediated decay (NMD), consistent with our observation that the alternative allele of this variant is associated with reduced expression of *TMED3* (both in our data and in AlphaGenome-predicted RNA-seq tracks, **Fig. 3e**).

Another example is the synonymous variant 19:51367458:C:T (rs143501994), which we found to be associated with increased expression of *CLDND2,* with the strongest effects in NK and CD8_TEM_ cells (*p*-value=8×10^−77^ and *p*-value=3×10^−75^ respectively). This variant lies at the penultimate base of exon 3 (NM_152353.3:c.429G>A, p.(Ala143=)). AlphaGenome predicts that it increases the strength of the canonical donor splice site in intron 3, promoting exon 3-4 splicing and increased *CLDND2* expression. This aligns with GTEx data showing frequent intron 3 retention in blood, fibroblasts, and lymphocytes, with retained transcripts expected to undergo non-stop decay (NSD). Enhanced splicing efficiency would therefore reduce NSD-mediated degradation and increase stable *CLDND2* transcript abundance, consistent with the single-cell RNA-seq association (**Supplementary Fig. 20**). Although SpliceAI predicts the wild-type donor splice site to be weak (0.12), it does not predict increased donor strength in the presence of the variant. Instead, SpliceAI predicts gain of a cryptic acceptor site (Acceptor gain = 0.93) that would introduce NSD, but this appears less likely based on the local sequence context.

### Quantifying the cell type specificity of immune cell eQTLs

An important question with single-cell eQTL analysis is the degree to which the loci exhibit genetic effects in specific cell types or subsets; in other words, how ‘cell type-specific’ are they? For several reasons, this is a complex question to address directly: genes are variably expressed across cell types, which can lead to true eQTLs being missed for lowly expressed genes; differences in cell numbers lead to variable power to detect eQTLs in a given cell type, making like for like comparison between cell types difficult; and linkage disequilibrium (LD) between loci means we cannot always directly infer if alternative top eQTL variants for the same gene in different cells are tagging the same or different causal loci. Existing methods to assess the extent of cell type specificity across conditions simultaneously (like mash^63^) can only assess genes that are sufficiently expressed across all conditions, which are only a subset of the total number (**Supplementary Fig. 21**).

Here, we defined five possible scenarios that can occur for a given eQTL identified at FDR<5% in one cell type (cell type A) for any of the pairwise comparisons with another cell type (cell type B; **Figure 4a**). In *scenario 1 (specific eGene)*, the eGene corresponding to the eQTL is not sufficiently expressed (<1% cells) in cell type B. In *scenario 2 (specific eQTL)*, the eGene is sufficiently expressed in cell type B, but no eQTL is identified. *Scenario 3 (distinct eQTL)* represents a case where two eQTL variants in low LD (*r*^2^ <0.5) for the same eGene are identified in cell types A and B. Conversely, we define a case where the same eQTL or an eQTL in LD (*r*^2^ ≥ 0.5) for the same eGene are found in cell types A and B as *scenario 4 (shared eQTL, concordant effect)* when the direction of allelic effect per additional minor allele on gene expression is concordant, or *scenario 5 (shared eQTL, discordant effect)* when the direction is discordant. Noting that an eQTL in *scenario 2* may have an underpowered association signal if cell type B is rarer than cell type A, we performed a pairwise random-effect meta-analysis and reclassified eQTL with potential association in cell type B (meta-analysis *p-*value < 10^−5^ x *p-value* in cell type A) as scenario 4 or 5 depending on the concordance of effect direction (**Methods**).

**Figure 4.**
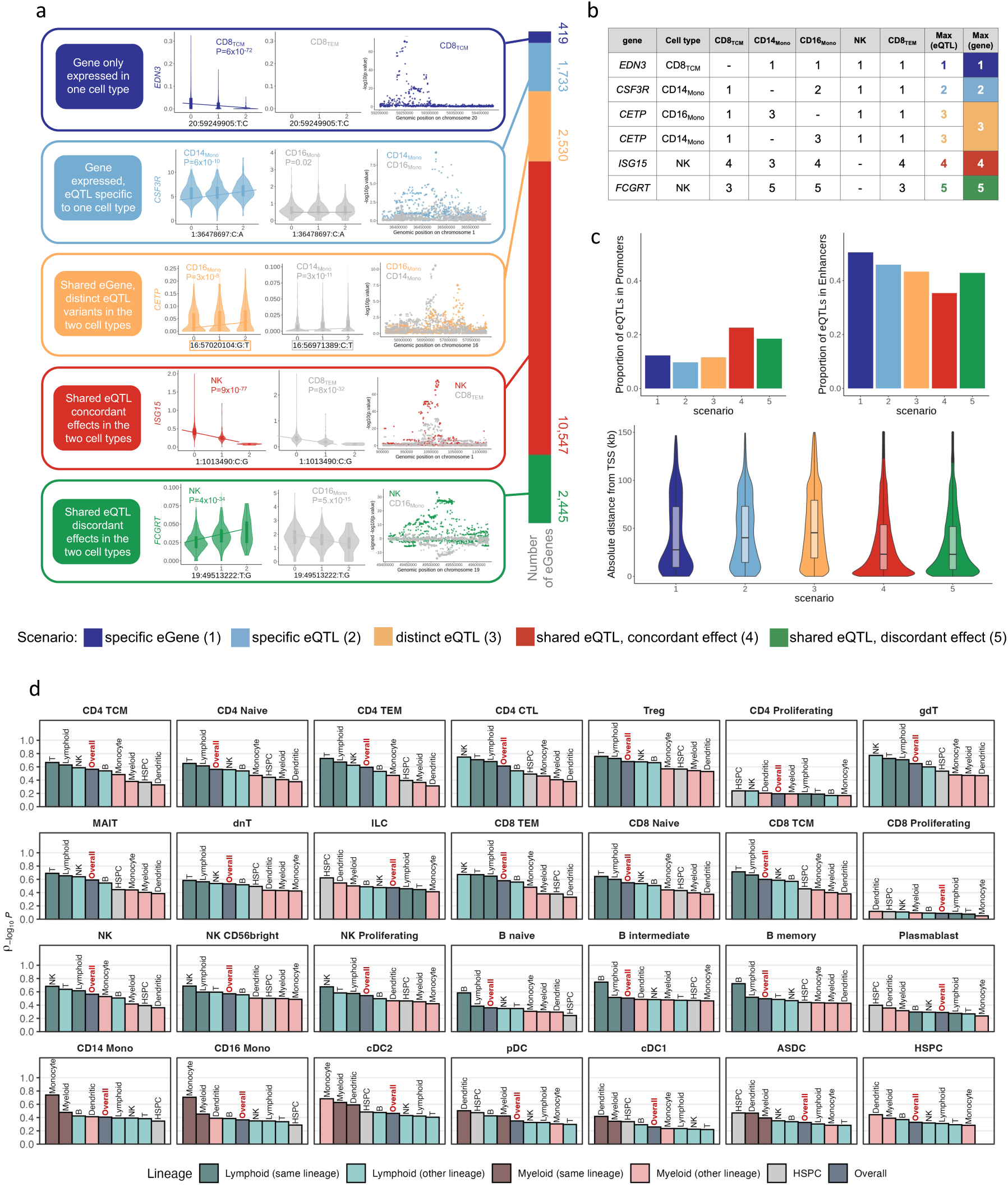
Cell type specificity analysis. **a)** Examples of common variant eQTLs illustrating the five scenarios of cell type specificity between two cell types (cell type A in each scenario’s colour, cell type B in grey). For each scenario, from left to right: brief description of the scenario, box plots showing the mean gene expression levels (y-axis) across genotype groups based on effect allele count (x-axis) for the lead SNPs in each of the two cell types, eQTL association signals measured as –log_10_(*p*-value) (y-axis) for all tested variants around the gene (ordered by base pair position, x-axis), with signal for both cell types overlaid together. Note that for scenario 3 we are highlighting the different eQTL variants in the two cell types. For scenario 5, eQTL association signal is calculated as –log_10_(*p*-value) multiplied by the sign of the eQTL effect, to emphasise the opposite direction of effect. The bar chart at the end shows the count of eGenes for each scenario, aggregated by calculating the maximum scenario number of all pairwise cell type comparisons (see **Methods**). **b)** Schematic to describe how the aggregated scenario is computed from eQTLs to eGenes using the maximum value. The same examples from **a** are used for clarity (full results in **Supplementary Table 4**). **c)** Characteristics of eQTLs in different scenarios, in terms of the portion of lead eQTL variants located in promoter or enhancer regions (top), and their distance from the transcription start site (TSS; bottom). **d)** Bar plots showing average correlation of –log_10_(*p*-value) eQTL association across cell types. The correlation coefficient is calculated as the pairwise Pearson’s correlation between –log_10_(*p*-value) for eQTL given cell type (the panel label) and each other cell type belonging to a cell type group (x-axis). The Overall group represents all 28 cell types under analysis. The bars are sorted from highest to lowest along the x-axis and coloured by lineage, with darker shades representing membership of the group. The Overall and HSPC groups are coloured dark and light grey respectively.

We assigned a scenario to each pair of the primary cell type with the other 27 cell types for each of the identified 154,932 common variant eQTLs (**Supplementary Table 4**). Next, we aggregated the eQTL scenarios for each pairwise comparison into a single score for 17,674 common variant eGenes by taking the *maximum* scenario value across 27 other eQTLs under contrast (as illustrated in **Figure 4b Supplementary Fig. 22; Methods**).

In the overall eGene set aggregation, we found 419 of 17,674 (2.4%) eGenes with a maximum score of 1, representing specific expression in only one cell type. For example, *EDN3* is only expressed in CD8 positive central memory T cells (CD8_TCM_) and has an eQTL at 20:59249905:T:C (rs259962), such that more copies of the C allele results in higher expression in CD8_TCM_ (**Fig. 4a,b**). A higher percentage of eGenes (805 out of 17,674, 4.6%) were expressed in at most two cell types, 7% in at most three cell types, and 15% in at most 7 (1/4 of all cell types). Just over half (9,723 out of 17,674, 55%) of all eGenes were sufficiently expressed in all 28 cell types (**Supplementary Fig. 23**).

Amongst eGenes that are sufficiently expressed in two or more cell types, 1,733 (9.8% of all eGenes) had a maximum score of 2, indicating the presence of an eQTL in only one of 28 cell types. For example, we found rs3917929 (1:36478697) as an eQTL for *CSF3R* only in classical monocytes (CD14_Mono;_ **Fig. 4a,b**). About half (8,506) of eGenes were significant in at most a quarter of all cell types (7/28), with only 35 eGenes (0.2%) found to have a significant eQTL in all 28 cell types (**Supplementary Fig. 24**).

A maximum overall score of 3, representing the presence of at least two distinct eQTLs (LD *r*^2^ < 0.5) for the same eGene in multiple cell types, was observed for 2,530 (∼14%) eGenes. For example, *CETP* is upregulated by different variants in classical (16:57020104:G:T, rs2052880) and non-classical (16:56971389:C:T, rs1532625) monocytes (**Fig. 4a,b**). The remaining eGenes showed evidence of a shared eQTL (same variant or two different variants with LD *r*^2^≥0.5) in two or more cell types, where 10,547 (∼60%) eGenes showed concordant direction of allelic effect on gene expression and 2,445 (∼14%) showed discordance. For example, the *FCGRT* gene was upregulated by 19:49513222:T:G (rs111981233) in NK cells but downregulated in non-classical monocytes (CD16_Mono;_ **Fig. 4a,b**), whereas *ISG15* was downregulated by 1:1013490:C:G (rs4615788) in both NK and CD8_TEM_ cells (**Fig. 4a,b**). The proportion of eGenes in different cell type specificity scenarios did not change dramatically when excluding the rarer cell types (**Supplementary Fig. 25**).

Next, we investigated the characteristics of eQTLs across different scenarios. Similar to observations by others^35^, cell type-specific eQTLs (scenarios 1-2) were generally found further away from the gene, and are more likely in enhancer regions as opposed to shared eQTLs (scenarios 4-5) which are more often in promoter regions (**Fig. 4c**). Distinct eQTL variants across cell types for the same eGene (scenario 3) resemble cell type-specific eQTLs more than shared ones according to these features.

As expected, eQTL sharedness between cell types was grouped by cell lineage and broad cell type categories (**Fig. 4d, Supplementary Fig. 26**). Sharedness within both lymphoid and myeloid lineages and major cell types (T, NK, B, Monocyte, Dendritic) was higher than outside of the lineage and major cell type, with the highest sharing in Monocytes (average correlation 0.72), and the lowest in dendritic cells (average correlation 0.49; **Supplementary Fig. 27a**). Excluding rare cell types (number of cells < 6,000; **Methods**), which are underpowered and skew the distributions, we found higher correlations for B and T cells (average correlations 0.82 and 0.75 respectively, **Supplementary Fig. 27b**).

An equivalent analysis on rare variants identified 2,560 cell type-specific effects (26%), either cell type-specific expression (*n*=75, ∼1%) or true cell type-specific regulation (*n*=2,485, 25%) and 7,445 shared (or likely shared; **Methods**) effects across cell types (74%; **Fig. 4, Supplementary Fig. 28; Methods; Supplementary Table 5**). Since rare variant analysis is gene-based, whereas common variant analysis is variant-based, the two results are not directly comparable. However, we observe a similar proportion (26%) of eGenes of which genetic regulation is more likely to be cell type-specific in both common variant analysis (scenario 1, 2, and 3) and rare variant analysis (scenario 1 and 2) analyses.

### Colocalisation analysis with disease phenotypes and blood traits

We used colocalisation analysis to identify the relationship between the cell type-specific eQTLs identified and genetic risk loci for several diseases and blood traits. We performed colocalisation with 16 diseases, including asthma^64^, autoimmune conditions (Crohn’s disease (CD)^65^, IgA nephropathy^66^, inflammatory bowel disease (IBD)^65^, rheumatoid arthritis (RA)^67^, systemic lupus erythematosus (SLE)^68^, type 1 diabetes (T1D)^69^ and ulcerative colitis (UC)^65^), cancer (breast^70^, lung^71^, lymphoma^50^ and prostate^72^), COVID-19^73^, and neuroinflammatory conditions (Alzheimer’s disease (AD)^74^, multiple sclerosis (MS)^75^ and Parkinson’s disease (PD)^76^) as well as 44 blood counts and traits from the UK BioBank^77^ (**Methods**). We restricted the analysis to common eGenes only (FDR<5%) and to genomic regions where at least one variant was genome-wide significant in the respective GWAS (*p*-value < 5×10^-8^).

We found strong colocalisation, indicating that the trait and cell type-specific gene expression are driven by the same variant (posterior probability of hypothesis 4, that the same variant drives both traits: PP H4 ≥ 0.8), for 5,287 genes in 60,458 gene-cell type-trait combinations. Of these, 5,491 colocalisation events (1,469 unique genes) were found for disease traits, 27,585 (3,572 genes) for blood serum traits, and 27,382 (3,331 genes) for full blood counts (**Supplementary Table 6**). Different cell types contributed to different traits and diseases, for example B cells most contributed to SLE, CD4+ T cells to rheumatoid arthritis, and dendritic cells to Alzheimer’s disease (**Fig. 5a; Supplementary Fig. 29**).

**Figure 5.**
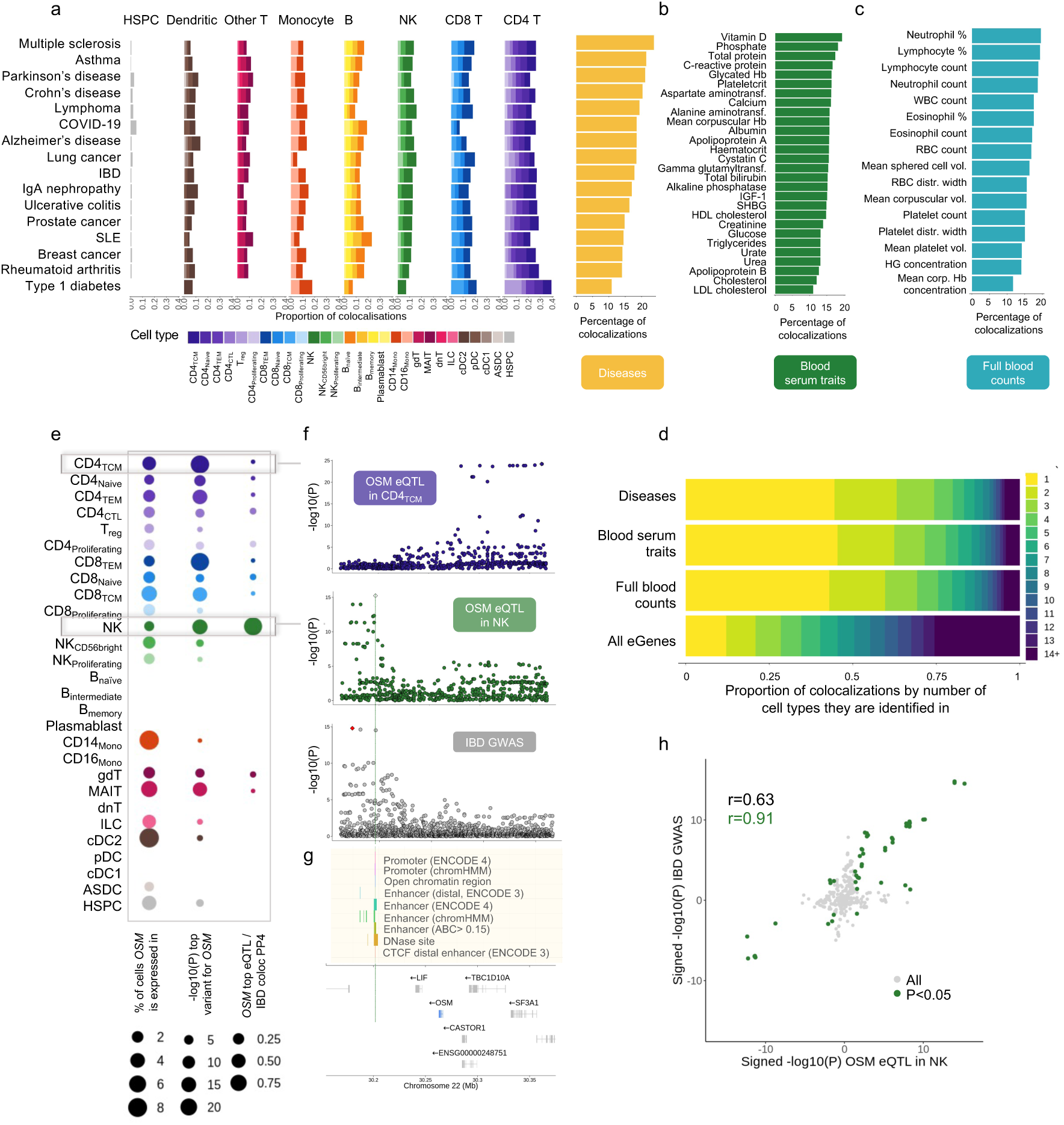
Colocalisation analysis with disease phenotypes and blood traits. **a)** Different cell types contribute to different diseases. Left: bar plots showing the proportion of colocalisation events for each trait, across the 28 cell types, coloured by cell type. Right: bar plots showing the proportion of disease-associated loci for which a colocalisation event (PP4>0.8) with at least one cell type was identified. **b,c)** Equivalent to the right most plot in a but for blood serum traits, and full blood counts respectively. **d)** Number of cell types each colocalisation event was found in, stratified by trait category. All eGenes are included for reference: the colocalisation events identified are very cell type-specific, especially the disease ones, much more than eQTLs without shown disease relevance. **e-h)** Cell type-specific colocalisation between an eQTL signal for *OSM* in NK cells and inflammatory bowel disease (IBD). **e)** Patterns of expression, eQTL signal, and colocalisation with IBD across cell types. *OSM* is broadly expressed and has eQTLs for many cell types, but the colocalisation with IBD is specific to NK cells. **f)** Locus zoom plots for *OSM* eQTL signal for CD4_TCM_ (top), *OSM* eQTL signal for NK (middle), IBD GWAS (bottom). **g)** Annotations for the likely causal *OSM* eQTL in NK, are highlighted, and the location of *OSM* within the region is shown at the bottom. **h)** Scatter plot of negative log_10_(*p*-values) multiplied by the direction of effect for the *OSM* eQTL in NK and the IBD GWAS signals, confirming the relationship between increased *OSM* expression in NK cells and increased IBD risk. IBD: inflammatory bowel disease, SLE: systemic erythematosus lupus, SSNS: steroid sensitive nephrotic syndrome, Hb: haemoglobin, IGF-1: Insulin-like Growth Factor 1, SHBG: Sex Hormone-Binding Globulin, aminotransf.: aminotransferase, RBC: red blood cell, WBC: white blood cell, vol.: volume, distr.: distribution, corp.: corpuscular.

These colocalisation events were remarkably cell type-specific: out of 16,993 unique gene-trait colocalisation pairs, 7,521 (44%) were uniquely found in one of the 28 cell types, and 13,073 (77%) identified in at most four cell types. This was consistent for disease traits (44%), blood serum traits (45%) and full blood counts (43%); when considering all eGenes the percentage is only 12% (**Fig. 5d; Supplementary Fig. 30**). While these cell type-specific colocalisation events were not always found for cell type-specific eGenes, the proportion of those (eGenes in scenarios 1,2 and 3 to some extent) for which we found a colocalisation event was significantly higher than for scenario 4 and 5 genes (**Supplementary Fig. 31**). Out of 5,288 genes with a significant colocalisation event, 893 (17%) were not previously reported as eGenes by the eQTLGen consortium, and of those, 49% were specific to a single cell type (and 68% to at most 2/28), highlighting the importance of cell type-specific maps of genetic regulation to both identify novel putative molecular mechanisms of disease variants, and identify the cell types that are most likely to contribute. We ran colocalisation with the same settings using our all-cell-type pseudobulk eQTLs and found a remarkable 5.2-fold increase in the number of colocalisation across traits (bringing the percentage of GWAS loci explained by an eQTL from 6.6 to 16.1% on average across traits; **Supplementary Fig. 32**).

For example, we identified a novel colocalisation event between expression levels of *OSM* and inflammatory bowel disease (IBD; PP4=0.99) that is specific to natural killer (NK) cells. The *OSM* gene encodes the Oncostatin M, a cytokine that contributes to many inflammatory diseases^78^. Additionally, the role of *OSM* in modifying the risk of IBD, specifically in NK cells, has been shown recently^79^. The top eQTL identified (22:30196498:C:G (rs713875)) leads to overexpression of *OSM* in NK cells in individuals with the IBD risk allele, consistent with previous observations that *OSM* levels are higher in IBD patients^80,81^. We show that *OSM* is expressed broadly (20/28 cell types), and has evidence of genetic regulation of expression in over a third of all cell types (11/28), yet only the regulatory variants active in NK (downstream of the gene) colocalise with the disease-associated variant (PP4 for all other cell types < 0.06), with the other signal affecting *OSM* expression in other cell types is located several kbp away, upstream of the gene and not in LD (*r*^2^=0.0005 between top NK and top CD4_TCM_ variant (22:30300755:C:A (rs9619096)), an example of a *scenario 3* eGene in our cell type specificity framework). Effect sizes were broadly concordant between eQTL and GWAS signal, confirming that increased expression of *OSM* in NK cells was associated with increased IBD risk (**Fig. 5e-h**).

We also identified a colocalisation event between Alzheimer’s disease (AD) and changes in expression levels of *IGHG4* specific to memory B cells (PP4=0.99; lead eQTL variant: 14:105721893:C:T (rs1134590)). The *IGHG4* gene encodes the Immunoglobulin heavy constant gamma 4 (IGHG4); while the involvement of the immune system in AD is relatively recent, it has been shown that significantly higher levels of immunoglobulin occur in late-stage AD patients^82^. *IGHG4* is expressed in a subset of memory B cells, which have been linked to AD: a possible mechanism is that depleted memory B cells may initiate Aβ cascade-related neuropathological events leading to AD^83^. In our data, *IGHG4* is expressed in B_intermediate_ and B_memory_ cells as well as in plasmablasts – consistent with the observation that class-switch recombination from IgM/IgD to IgG/A/E only happens after B cell activation^84^ – and the colocalisation with AD stronger in B_memory_ compared to B_intermediate_ (PP4=0.69). The direction of effect is consistent, indicating that higher *IGHG4* expression corresponds to higher AD risk (**Supplementary Fig. 33**).

Additionally, we identified a colocalisation event between IBD and changes in expression levels of *TNFSF15* in dendritic cells, specifically Plasmacytoid dendritic cells (pDC, PP4=0.99; lead eQTL variant: 9:114796423:C:T (rs6478108)), type 2 conventional dendritic cells (cDC2, PP4=0.98) and AXL+SIGLEC6+ dendritic cells (ASDC, PP4=0.95). *TNFSF15* encodes the TNF-like cytokine 1A (TL1A)^85^, which is a key regulator of inflammation and the only known ligand of the death-domain receptor 3 (DR3, encoded by *TNFRSF25*). *TNFSF15* was one of the first risk loci defined by the GWAS approach for IBD^86^, and has the largest effect size in populations of East Asian ancestry^87^. In addition, a *trans*-ancestry meta-analysis has implicated a GTEx *cis*-eQTL for *TNFRSF25* (rs2986751; which we also identify as an eQTL for *TNFRSF25* – and no other gene – in our own data), in IBD risk^88^. TL1A inhibition is an active area of drug development^89^, with the anti-TL1A monoclonal antibody *tulisokibart* having recently shown efficacy in phase II trials for moderate to severe ulcerative colitis^90^. In our data, *TNFSF15* is uniquely expressed (>1% of cells) in the four dendritic cell types, and has an eQTL in three out of four. The effect size of the top variants is negative, indicating that lower expression of *TNFSF15* corresponds to higher IBD risk, consistent with similar results previously reported in monocytes^91^ **Supplementary Fig. 34**).

Other examples of novel colocalisation events identified were between the expression of the *PYCARD* gene in CD8_TEM_ (lead variant: 16:31258718:A:ACACAC (rs58732039)) and both SLE (PP4=0.91) and IgAN (PP4=0.9); and a colocalisation between a non-classical monocyte (CD16_Mono_)-specific eQTL (lead variant: 5:149822705:C:A (rs1078324)) for *PPARGC1B* and SLE (PP4=0.94; **Supplementary Fig. 35**).

### Identification of cell state eQTLs

We aimed to identify eQTL effects that were modulated by continuous cellular states beyond discrete cell types. A challenge with these analyses is the definition of cell states, with existing studies typically going one of two ways: an unsupervised approach to determine axes of variability (often using principal components or related methods^21,22^) or using one or few very well-defined variables, such as disease state^92,93^ or sex and age^92^. In the first case, these factors tend to be difficult to interpret; the latter approach misses out on novel cell states and contexts that are yet to be characterised. Here, we used an in-house deep learning method – scDeepID – to identify sub cell types, states and phenotypes that are biologically informed yet general. Collectively, scDeepID is a multi-task transformer capable of cell type annotation, cell function identification and cell simulation (additional detail in **Methods**).

We took NK cells as an example, because sub cell type heterogeneity and function have been characterised at cellular resolution in a recent publication^94^. First, we performed label transfer with scDeepID from their integrated scRNA-seq data of NK cells^94^ which allowed us to identify three subtypes within the macro-NK cells defined previously: NK_adaptive_ (most involved in cell activation), NK_mature_ (most cytotoxic) and NK_intermediate_ (showing signatures of both NK_mature_ and NK_CD56bright_ subtypes), with good match to the original publication based on the expression of key markers (**Supplementary Fig. 36**). Together, we have five different subtypes for NK cells, including the NK_CD56bright_ and NK_Proliferating_ cells (**Fig. 6a**). Next, we sought to identify transient and functionally meaningful states within and across these cell subtypes by training a scDeepID model with GO biological process (GOBP) database including the terms previously identified as key functions in NKs. They are: cell killing, leukocyte cell cell adhesion, myeloid leukocyte differentiation, regulation of cytoskeleton organisation, and canonical NF-κB signal transduction to identify the cell state variation in NK cells (**Fig. 6b; Methods**).

**Figure 6.**
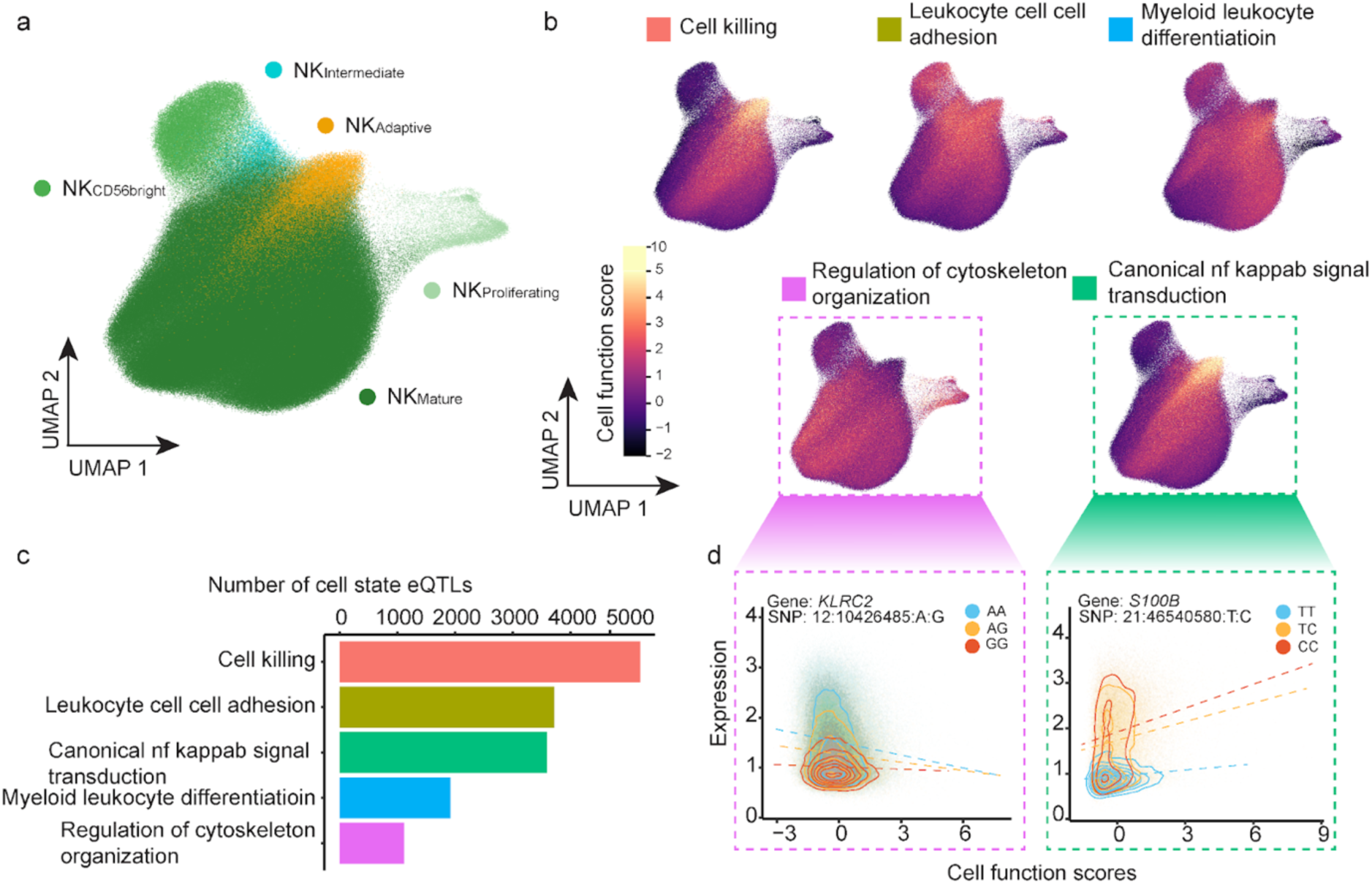
Identifying cell state eQTLs in NK cells. **a)** Cell function score-based UMAP, coloured by different subtypes within NK cells, broadly matching those described in Rebuffet *et al*^94^**. b)** Visualisation of five representative cell functions as continuous cell states, UMAP representation from **a. c)** Number of cell state eQTLs identified with significant interaction with the five cell states from **b. d**) Examples of cell state-specific sc-eQTLs**. left** The relationship between genotype groups at 21:46540580:T:C and NK expression of *S100B* is modulated by the NF-kappaB signaling function, with activated cells expressing higher values of *S100B* for C/C individuals (red) but not T/T (blue). **right** Similarly, relationship between *KLRC2* expression and the regulation of cytoskeleton organisation function when stratifying cells based on genotype group at 12:10426485:A:G.

To identify genetic regulation of gene expression mediated by these cell states, we mapped dynamic eQTLs using a linear mixed model with an interaction term and the cell function scores described above, which represent biologically-meaningful continuous cell states (**Methods**). We confirmed controlled type 1 error using permutations (**Supplementary Fig. 37**). We consider all NK cells (all five subtypes described above, *n*=850,833); all gene-SNP pairs that displayed evidence of eQTL effects (FDR<5%) in any of NK, NK_Proliferating_ or NK_CD56bright_ (16,759 unique gene-SNP pairs; **Methods**) and the five cell states described above to identify gene-by-context (GxC) dynamic effects. In total, we identify 15,535 cell-state eQTLs in NK cells (for 7,709 unique gene-SNP pairs, 46% **Supplementary Fig. 38**), across the five selected cell functions, with cell killing, leukocyte cell cell adhesion and myeloid leukocyte differentiation having the most dynamic eQTLs (**Fig. 6c**).

For example, an eQTL for *S100B* (21:46540580:T:C, rs9306156) is modulated in strength by the positive regulation of the canonical NF KAPPAB signal transduction function, with higher value of the function correlating with higher *S100B* expression for individuals carrying the C allele, but not for T/T homozygous individuals (**Fig. 6d**). 21:46540580:T:C was previously identified as an eQTL for *S100B* in NK cells in the OneK1K study^27^, is associated with changes in lymphocyte counts^95^, and is found to be in an NK cell-specific enhancer by ENCODE^96^. When secreted, *S100B* acts as a signaling molecule that can bind to the receptor RAGE (Receptor for Advanced Glycation End-products), influencing inflammation. This result may indicate that individuals with the C allele at this locus might mount stronger inflammatory responses by producing more extracellular S100B when NKs are activated, possibly leading to heightened NF-κB/S100B feed-forward loops via a cell type-specific enhancer mechanism.

Another example is a dynamic eQTL for *KLRC2*, associated with regulation of cytoskeleton organisation (12:10426485:A:G, rs12318583). We find that *KLRC2* expression is highest in low-function cells and declines as function increases in A/A carriers, whereas it remains low and stable in G/G individuals (**Fig. 6d**). *KLRC2* encodes NKG2C, an activating receptor predominantly expressed on NK cells, which signals through adaptor proteins such as DAP12 to trigger cytoskeletal rearrangements necessary for NK cell activation, immune synapse formation, and target cell killing. The cell state-specific dynamic suggest that in A carriers, higher *KLRC2* levels in low-function cells may prime NK cells for rapid cytoskeletal remodeling upon activation, whereas the G/G genotype maintains consistently low receptor levels, potentially limiting cytoskeleton-dependent functional responses.

An additional example is an eQTL for *RPS26* (12:56042145:C:G, rs1131017) which is modulated in strength by the cell killing function, with the gene regulatory effect of allele G increasing *RPS26* expression levels being strongest in more cytotoxic cells (**Supplementary Fig. 39**).

### Cell state abundance QTLs

We sought to identify genetic variants that alter the composition of states directly by applying Genotype–Neighborhood Associations (GeNA^97^) to test for cell state abundance QTLs (csaQTLs). We mapped csaQTLs in 8 major cell type categories (B cells, CD4+ T cells, CD8+ T cells, other T cells, NK cells, monocytes, dendritic cells, and HSPCs, **Supplementary Fig. 40**). We confirmed Type I error was controlled using permutations after filtering to common genetic variants at MAF>5% (as we found some inflated results for variants between 1-5% MAF, **Supplementary Fig. 41,42; Methods**).

In total, we identified 164 independent csaQTLs passing genome-wide significance (*p*-value < 5×10^−8^), with the highest numbers discovered in monocytes, dendritic cells, and NK cells (**Fig. 7a**). The discordance between csaQTL discovery power and cell type abundance may reflect differential contribution of environmental and genetic factors to cell state abundance traits within immune subpopulations. NK, dendritic, and monocyte populations have been previously described as displaying high heritability for cell state abundance traits compared to other PBMC subsets^98–102^. Our analysis replicated four out of five total csaQTLs previously identified in the OneK1K dataset^97^ (**Supplementary Fig. 43**). We found three csaQTL variants significant in both analyses and one locus significant in the OneK1K analysis that was suggestive in our analysis (*p-*value = 7.1×10^−8^), in moderate LD with a lead variant (*r*^2^=0.46) displaying a matching association with depletion of NK_CD56bright_ cells (**Supplementary Table 7**).

**Figure 7.**
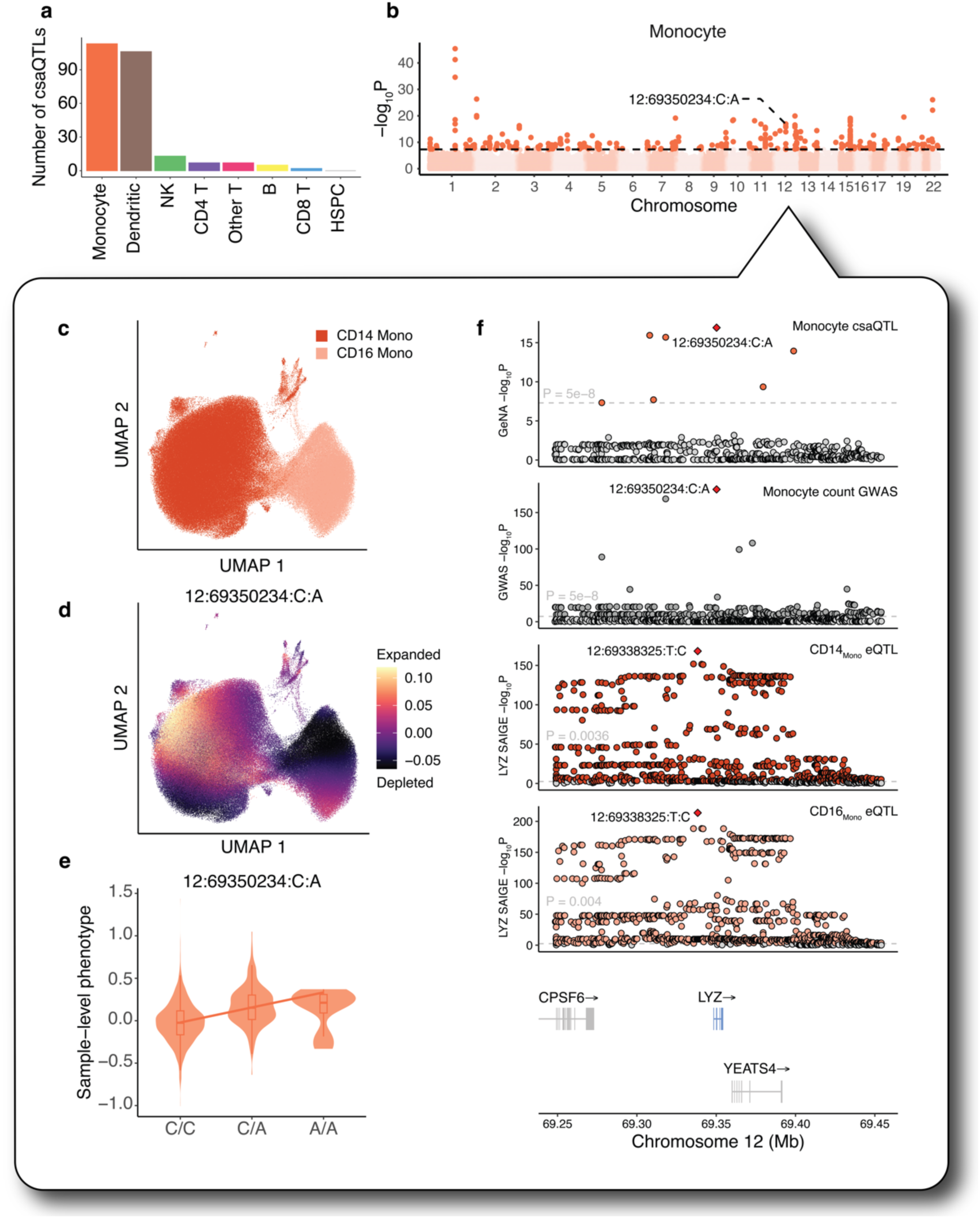
Identifying cell state abundance QTLs. **a)** Number of significant (*p*-value < 5×10^−8^) lead csaQTL loci per major cell type. **b)** Manhattan plot illustrating csaQTLs identified with GeNA for monocytes. Dashed lines indicate the genome-wide significance threshold (*p*-value < 5×10^−8^), and loci passing this threshold are coloured according to the cell type they were significant in. The *LYZ* missense variant 12:69350234:C:A is highlighted. **c)** UMAP projection of monocytes, coloured by cell type. **d)** UMAP projection of monocytes, coloured by GeNA neighborhood-level phenotype values (correlation between cell abundance and alternative allele dose per neighborhood) for 12:69350234:C:A. These values describe which cell populations are expanded or depleted. Colours are scaled between the 5th and 95th percentile. **e)** Sample-level phenotype values (linear combination of sample loadings on NAM-PCs used in the fitted model for the lead SNP) for the *LYZ* variant (12:69350234:C:A). **f)** Locus zoom plots for the csaQTL in monocytes, the monocyte count GWAS and the SAIGE-QTL results for *LYZ* in both classical and non-classical monocytes. The bottom plot shows the location of *LYZ* and other genes in the region on chromosome 12.

We discovered novel csaQTLs associated with abundance shifts in both discrete cell types, and continuous cell states. One example is the monocyte csaQTL 12:69350234:C:A (*p*-value = 1.2×10^−17^, **Fig. 7b-e**), which has also been associated with monocyte counts in a GWAS by Vukovic *et al,*^103^, and which we confirmed to share a causal signal via colocalisation (PP4 > 0.99; Methods) This variant is a missense change in *LYZ* (T88N), overlaps an sc-eQTL for *LYZ* in both CD14⁺ and CD16⁺ monocytes, and additionally colocalises with an LYZ eQTL in HSPCs (**Supplementary Fig. 44a**). *LYZ* encodes lysozyme, an antimicrobial effector secreted by phagocytes that hydrolyses peptidoglycan in bacterial cell walls, and shows myeloid-specific expression (**Supplementary Fig. 44b**). To further characterise this csaQTL, we performed gene-set enrichment analysis on the top genes correlated with the associated cell state. Enriched pathways included inflammatory response (BH-adjusted *p*-value = 9.03×10^−8^) and TNFα signalling via NF-κB (BH-adjusted p-value = 7.87×10^−8^), suggesting that this csaQTL modulates the abundance of a pro-inflammatory monocyte/macrophage state (**Supplementary Fig. 45**). We also identified a monocyte csaQTL 12:9953308:T:TG (*p*-value = 7.0×10^−14^, **Supplementary Fig. 46a,b**) that intersected a *CLEC12A* eQTL in CD16 monocytes **Supplementary Fig. 46c**). *CLEC12A* encodes a C-type lectin which inhibits monocyte and granulocyte activation^104^, and may regulate inflammation in response to cell death^105^. *CLEC12A* has also been described as a marker for leukemic stem cells and is a possible drug target in acute myeloid leukaemia^106^. The cell state associated with this variant exhibited higher expression of gene sets including inflammatory response, TNFα signalling via NF-κB, and IL2 and IL6 signalling (**Supplementary Fig 45**).

## Discussion

In this paper and accompanying manuscripts, we present the largest map to date of the effects of genetic variation on gene expression and chromatin accessibility at cellular resolution in human immune cells. In this flagship paper, we mapped the effects of common and rare SNPs and indels called from whole-genome sequencing in 1,925 individuals and 28 immune cell types and identified more than 150,000 common variant eQTLs for 83% of the genes tested and over 30,000 gene-level rare variant signals for nearly 50% of all genes. For common variants, especially for the most abundant cell types, we find an eQTL for the majority of genes tested, noting that the expected relationship between number of cells and number of eGenes begins to plateau for the four most abundant cell types (number of cells >500,000) suggesting that this may be the number necessary to begin saturating common variant effects within a cell type. For rare variants, the relationship between number of cells and number of eGenes is linear, suggesting further increases in sample size and more sophisticated selection of variants based on functional annotation will continue to yield additional discoveries (**Supplementary Fig. 6**).

For common variants, we identify nearly three times more eGenes compared to OneK1K^27^, the previously largest single-cell eQTL study – due to a combination of factors: larger sample size, more cells per individual, more granular resolution of cell types, and improved single cell-specific association methods. When comparing to eQTLs from sorted immune cell types followed like bulk transcriptomics (*e.g.,* the BLUEPRINT^7^ study) we found that these studies could be similarly powered to us despite the smaller sample size for very well-defined cell type populations (*e.g.,* CD14+ monocytes) but much less well powered for more heterogeneous cell types (like CD4+ T cells, which can be separated into six subpopulations in TenK10K). It is worth noting that these studies are limited to known cell types with robust cell surface markers and, even then, are labour-intensive.

Finally, we identified almost exactly the same number of unique eGenes as the eQTLGen program^2^, the largest eQTL map using whole blood ‘bulk’ transcriptomics from >30,000 people, despite a 15-fold lower number of individuals. About 25% of eQTLGen eGenes were not detected in our study: these were very lowly expressed in all of our cell types; in contrast, ∼25% of TenK10K eGenes were not found in eQTLGen, and these were more cell type-specific, emphasising the value of single cell approaches for identifying cell type-specific molecular variation that is difficult or impossible to observe in bulk transcriptome methods.

When mapping eQTLs across increasing numbers of cell types, an important and often non-trivial question is the extent to which these eQTLs are specific or shared. We propose a framework to distinguish between five possible scenarios that start with pairwise comparisons between two cell types (**Fig. 3**). Aggregating all pairwise combinations yields a specificity score for each identified eQTL, which will be a useful framework to quantify cell type specificity in future studies. Unlike existing approaches, this framework is not limited to genes tested in all cell types. It could be improved by addressing some limitations: the threshold used to define cell type specificity is based on data-driven observations (**Supplementary Fig. 47**) and conservative, but no independent dataset of comparable or larger sample size was available to evaluate the winner’s curse bias; we used the heuristic cutoff of LD=0.5 to determine whether an eQTL was shared or not across cell types in case of distinct variants – different cutoffs may be more appropriate for different applications.

In this study, about 50% of eQTLs were specific to one or a subset of cell types, and very few were ubiquitous to all, even when accounting for differences in power, demonstrating the importance of eQTL maps across different cell types – especially rare ones that would be missed in tissue-level maps. Cell type-specific eGenes were also more likely to have a colocalisation event with at least one trait (8-fold increase between *scenarios 2* and *4*, **Supplementary Fig. 31**), suggesting that eQTL maps in increasingly granular and harder to access cell types (and states) may be key to bridging the gap between eQTL and GWAS variants^107^. We confirmed this by mapping eQTLs in an all-cell-type pseudobulk we constructed including aggregate expression across all cells regardless of cell type. This analysis showed that the single-cell approach identifies ∼1.5 times as many eGenes and, an average, 2.6 times as many colocalisation events across our 60 traits.

An important advancement compared to existing single-cell eQTL studies was our ability to map the effects of rare and low-frequency genetic variants on gene expression using genotypes called from WGS and set-based tests implemented in SAIGE-QTL^40^. Running collapsing analyses with different settings provides a glimpse into which variants may have a greater impact on genetic regulation. However, larger sample sizes and more specific, better annotations will be crucial for a more in-depth understanding. First, we found that the SKAT test was significantly better powered than burden, likely reflecting the expectation that *cis* variants have mixed effect directions on gene expression changes.

Second, we did not have sufficient power to test different sets of functional annotations separately, although restricting to variants with at least one of these annotations did improve our detection power. More cell type-specific regulatory annotations are also needed to improve this analysis. While we have scATAC-seq from matched cell types^39^, we are currently under-powered for a straightforward analysis of variants in cell type-specific open chromatin regions and had to incorporate them with other functionally annotated regions that were not cell type-specific, potentially diluting the signal.

This dataset also allows us to explore genetic effects that are modulated not only by cell type but also cell states. Using natural killer cells and a biologically-driven deep learning method, we first define cellular states corresponding to specific functions (for example, cell killing) and then test for the effects that these have on eQTLs, identifying over 15,000 cell state eQTLs in NK cells. Approaches like this are promising to identify eQTLs that vary dynamically across biologically relevant contexts and functions, which are likely widespread^10^; however, existing complex methods do not scale well, so we used a simpler model and limited our analysis to regular eQTLs, potentially missing dynamic-only eQTLs.

In addition to exploring the impact of genetic variation on the expression of genes *within* defined cell states, we mapped the effects of genetic variants on the composition of cell states within each major cell type, which have been associated with disease risk^103,108–109^. Using a recently proposed method^97^, we identify 164 cell state abundance QTLs (csaQTLs) across 8 major cell types. We detected most of our csaQTLs for monocytes, dendritic cells, and NK cells, consistent with previous reports^97^: this may be due to the higher estimated heritability of abundance of NK and myeloid cell types compared to other blood cell populations, or T and B cell states may be better represented by cell surface markers, which scRNA-seq does not capture. We explored validation of our csaQTLs by colocalising them with cell count GWAS^77,103^. This comparison revealed limited overlap, likely explained by low statistical power for csaQTLs, as well as limited biological overlap: the vast majority of the csaQTLs detected do not align with discrete cell types but rather continuous cell states, whereas the cell type categories measured in the cell count GWAS are quite broad (*e.g.*, ‘Lymphocyte’, which includes all T, B and NK cells in our data), or are not PBMCs and thus not present in our dataset (*e.g.*, ‘Neutrophil’). In summary, it is likely that the csaQTLs we detect have an effect on the composition of granular cell states within these broad categories but do not substantially affect their overall count.

In accompanying TenK10K manuscripts, we explore additional aspects of human cell type-specific molecular variation. We investigate the impact of tandem repeat length polymorphism on cell type-specific gene expression^37^, identifying over 60,000 eQTLs, many of them independent of the eQTLs described in this manuscript. In other manuscripts we further characterise the putative direction of causal relationship between molecular QTLs and human phenotypic associations^38^, and the impact of genetic variation on measures of cell type-specific chromatin accessibility, a critical upstream marker of regulatory impacts on gene expression^39^. Together, these publications demonstrate the power of population-scale cellular genomics^20^. We show that sequencing-based approaches to characterising genetic variation, ideally whole-genome sequencing, are critical for accessing both low-frequency variants and complex classes of structural variation. Single-cell methods for studying both gene expression and chromatin accessibility phenotypes provide a massive increase in discovery power and the opportunity to access cell type-specific effects, which constitute most genetic impacts on molecular phenotypes.

Population cellular genomic datasets provide an important lens on the impact of gene perturbation on whole-organism molecular physiology, complementing high-throughput experimental perturbation approaches, and will be critical to build increasingly accurate models of human biology. Further expansions in the sample size of population cellular genomic studies and increasing the diversity of participants and tissues – including individuals of diverse ancestry and from a wide variety of active disease states, and a much wider variety of tissue and cell types – will be important for building a comprehensive understanding of the molecular origins of human disease.

## Methods

### Cohorts

Blood samples were collected from 2,339 consenting individuals from two Australian biomedical research cohorts: the Tasmanian Ophthalmic Biobank (TOB) [ethics protocol 2020/ETH02479] and the BioHEART study [ethics protocol 2019/ETH08376] cohort.

The TOB cohort consists of 983 individuals (562 females, 421 males) recruited between January and February 2018 by the Royal Hobart Hospital and Menzies Institute of Medical Research in Tasmania, Australia. The selection criteria for inclusion in the cohort were as follows: (1) age 18 years and over, (2) no signs of ocular disease, and (3) self-reported British, Scottish, or Irish ancestry.

The BioHEART^110^ cohort included 1,356 individuals (561 females and 795 males) recruited between March 29th, 2016 and May 2nd, 2023, presenting to study sites for clinically indicated cardiovascular imaging or acutely with ST elevation myocardial infarction. The selection criteria for inclusion in the cohort were as follows: (1) age 18 years and over, and (2) presenting with cardiac disorders (or for the investigation of suspected cardiac disorders) including: acute myocardial infarction, suspected coronary artery disease, patients undergoing cardiothoracic or vascular surgery, or plastic surgery involving resection of vascularised tissue.

After donor, variant, and cell QC, including selection to individuals of European ancestry only, 1,925 individuals remained: 950 from TOB and 975 from BioHEART (**Supplementary Fig. 1**).

### Blood collection and DNA extraction

For the TOB cohort^27^, blood samples were collected from each participant and stored in 1 x 8mL heparin cell preparation tubes (CPT) and 1× 9mL EDTA tubes. For the BioHEART cohort, aliquots were stored at –80C.

DNA extraction from BioHEART buffy coat samples was performed by the Australian Genome Research Facility (AGRF) using QIAamp96 DNA QIAcube HT kit. DNA extraction from TOB samples was performed at the Menzies Institute of Medical Research (University of Tasmania) using the QIAmp Blood Mini Kit. For a subset of TOB samples that failed initial quality control checks during sequencing, DNA was extracted from PBMCs a second time using the Qiagen DNeasy Blood and Tissue Kit and eluted in an AE buffer by the Garvan Molecular Genetics lab service. DNA concentration was measured for all samples using fluorometric quantitation with the Quantifluor from Promega prior to sequencing.

### Whole-genome sequencing data

Whole-genome sequencing (WGS) data was generated for all individuals from the TOB cohort (*n*=983) and BioHEART (*n*=1,356).

#### Sequencing library preparation and whole-genome sequencing

Sequencing library preparation was performed using the KAPA PCR-free v2.1 kit (Roche) for the TOB samples, and the DNA PCR-free kit (Illumina) for the BioHEART samples. Short-read whole genome sequencing was performed using Illumina 2 x 150 bp chemistry on the NovaSeq 6000 instrument to achieve mean 30x coverage. Library preparation and sequencing were performed by the Garvan Sequencing Platform.

#### Alignment and variant calling

Alignment and variant calling were performed in line with GATK-DRAGEN Best Practices. Reads were aligned from FASTQ (BioHEART) or CRAMs (TOB) against the GRCh38 reference genome (masked version, as available at <u>gs://gcp-public-data--broad-references/hg38/v0/dragen_reference/</u>) using a fork of DRAGMAP v1.3.1 (https://github.com/populationgenomics/DRAGMAP). When aligning from CRAMs, bazam (https://github.com/ssadedin/bazam) was used to first convert the reads back to FASTQ format. Calling of SNPs and indels was then performed using GATK4 HaplotypeCaller in DRAGEN mode. The resulting gVCF files were combined using hail’s VDS Combiner and stored in the variant dataset (VDS) format.

#### Sample QC

Hard sample filtering was applied to remove individuals with: high rates of contamination (FREEMIX ≥ 5%); an elevated percentage of chimeras (PCT_CHIMERAS ≥ 6.3%); median insert size < 250bp; and sex aneuploidy or discordance between reported and observed sex. The dataset was also inspected for outlier samples based on the number of singletons, het/hom ratio, Ts/Tv ratio, and ins/del ratio, taking care not to exclude ancestry outliers based on these metrics.

A set of high-quality sites, defined as an LD-pruned subset of autosomal, biallelic, allele frequency > 0.01, call rate > 99% and InbreedingCoeff > –0.25, were extracted from 1000 Genomes and HGDP. Samples with call rate < 0.85 across these sites were also excluded.

Following hard sample filtering, related individuals were identified using hail’s implementation of PC-Relate and confirmed using somalier relate v0.2.15 (https://github.com/brentp/somalier). For family sets consisting of second-degree or closer relatives, as defined by a kinship coefficient > 0.088, the sample with the highest quality single cell RNA-seq data was retained, as determined by the number of cells and cell type composition. When equal quality single cell data was available, the sample with the highest mean genome sequencing depth on chromosome 20 was retained.

Ancestry inference was then performed on the high-quality, unrelated set of samples, by combining this set with background samples from 1000 Genomes and HGDP, then computing principal components using gnomAD’s run_pca_with_relateds method and the same set of high-quality sites defined for call rate estimation above. Ancestry labels were assigned based on genetic similarity, as identified through principal component analysis, to those of known ancestry in the background datasets. For association testing, only samples assigned to the European cluster were retained (**Supplementary Fig. 48**).

#### Variant QC

Allele-specific Variant Quality Score Recalibration or AS-VQSR (GATK v4.2.6.1) was run using all QC-pass, unrelated samples (regardless of ancestry), using the default training resources and priors. For SNPs this is hapmap (prior=15), omni (prior=12), 1000G (prior=10) and dbsnp (prior=7). For indels this is mills (prior=12), axiomPoly (prior=10), and dbsnp (prior=2). The following features were included for all SNPs and indels: AS_ReadPosRankSum, AS_MQRankSum, AS_QD, AS_FS, AS_SOR. AS_MQ was used for SNPs only. All sites failing on the site-level filter calculated by AS-VQSR were removed.

Within the European-only association callset, we then removed any variants that had no high-quality genotype calls (defined as GQ>=20, DP>=10 and allele balance > 0.2 for heterozygotes) or with an excess of heterozygotes at that site compared to Hardy-Weinberg expectations (InbreedingCoeff < – 0.3). A final filter to remove sites with missingness > 15% was then applied prior to association testing.

### Single-cell RNA sequencing data

Single-cell RNA-seq data was collected from peripheral blood mononuclear cells (PBMCs) from samples from individuals from the Tasmanian Ophthalmic Biobank (TOB) (OneK1K^27^; *n*=1,036) and BioHEART^110^ (*n*=1,012) cohorts. **Supplementary Fig. 49** illustrates the experimental workflow and key steps.

#### Sample pooling and cell count estimation

TOB samples were previously pooled during preparation of the OneK1K project, with pools made up of 12-14 individuals^27^. In order to capture more cells per individual (aiming for up to 5,000 cells per individual), two pools (*n*=24-28) at a time were further pooled (weighted by cell viability) and loaded into 3 sequencing lanes (aiming for 40,000 cells each, so 120,000 in total). Additionally, “maxi pools” of 4 combined pools (96-112 individuals) were sequenced in two separate lanes to compensate for possible low cell numbers.

In contrast, BioHEART samples were pooled (weighted by cell viability) for the first time for this project, each pool comprises 8 individuals and is sequenced in a separate sequencing lane, aiming for 40,000 cells (∼5,000 cells per individual). In some cases, pools with low cell viability were sequenced again to rescue cell numbers and get closer to the target.

#### Capture and sequencing

Cells were captured using the single-cell 3’ gene expression high throughput (3’ GEX HT) kit v3.1 from 10X on a Chromium X machine. For the TOB cohort, cells were re-captured starting with the original biomaterial (OneK1K individuals) that we had biobanked and using the new 10x chemistry and with the new high-throughput kit. The long-term cryopreservation of these samples led to a smaller proportion of monocytes in the TOB cohort compared to BioHEART (**Supplementary Note 1**). Library preparation and short-read sequencing were performed using illumina’s NovaSeq 6,000 machine.

#### Alignment

scRNA-seq reads were aligned to a genome reference (GRCh38, Gencode release 44, Ensembl 110) using STAR^111^, as implemented within 10X’s CellRanger software (v7.2.0) which also estimates the number of cells captured (**Supplementary Fig. 50**). CellBender^112^ (v0.3.0) estimated numbers of cells differed significantly from those estimated by CellRanger, leading us to discard these results (**Supplementary Note 2**).

#### Demultiplexing and doublet detection

To assign cells to their donor of origin, as well as to remove doublets (droplets containing more than one cell), we used Demuxafy^113^ (version: 2.1.0), a tool to runs a number of demultiplexing and double detection software and outputs a consensus call for each individual cell. Using *Demuxafy*, we ran *vireo*^114^ to demultiplex cells to their donor of origin within each sequencing library, using genotypes extracted from either SNP array data (available as part of the OneK1K data; for TOB) or using common (MAF>5%) biallelic SNPs from WGS (for BioHEART). A handful of individuals did not have available genotype data, but *vireo* allows us to include the expected number of individuals in each pool with or without genotypes, which makes the donor assignment and doublet detection more accurate. For doublet detection, we used *Demuxafy* to run scds^115^ and *ScDblFinder*^116^, and used majority voting between all three tools (*vireo*, *scds* and *scDblFinder*) to determine the final set of doublets for each sequencing library (**Supplementary Fig. 51**).

#### Cell typing

To assign individual cells to specific immune cell types we follow the recommendations of the single-cell eQTLGen consortium^41^. We use a combined approach that uses *scPred*^117^, hierarchical progressive learning, scHPL^118^, and Azimuth as the reference dataset^42^ as detailed in https://powellgenomicslab.github.io/WG2-pipeline-classification-docs/index.html. To run this step, we converted our single-cell Scanpy object into Seurat^119^ objects. We also used CellTypist^120,121^ (v=1.6.2) to confirm cell typing of individual cells, with good concordance (**Supplementary Fig. 52**). Since the former approach provided more granularity (28 cell types vs 18 identified by CellTypist), we proceeded with the 28 cell types for the remainder of the manuscript.

#### Quality Control

Next, we performed quality control (QC) with Scanpy (v1.8.2)^122^. For each library, we removed cells with >20% mitochondrial reads, <800 reads detected, and <1000 or >10,000 genes detected (**Supplementary Fig. 53**). We also removed individuals with abnormal cell type composition and individuals with fewer than 100 cells.

Overall, after cell and sample QC (based on scRNA-seq and WGS), a total of 5,438,679 cells from 1,925 individuals were left for consequent analyses (**Supplementary Fig. 1**).

### Association testing

#### SAIGE-QTL

To test for the effects of sets of both common and rare variant associations with single-cell gene expression, we used the recently proposed SAIGE-QTL method^40^. Briefly, SAIGE-QTL implements a Poisson mixed model, thus effectively modelling single-cell counts and repeat cells for the same individual. Additionally, several computational and statistical speed-ups are implemented to improve speed, scalability, and storage usage. Finally, implementation of both single-variant tests (where effects of the genotype at a single locus are assessed), as well as set-based tests (where signals from multiple variants within a region are assessed together in their combined effect), is available, as briefly described below.

We mapped eQTLs separately in each of the 28 cell types. We confirmed that our tests were calibrated by shuffling the individual IDs and re-performing the test for our most abundant cell type, CD4 positive central memory T cells (CD4_TCM_), the results of which show no evidence of false positives for both common and rare variant tests (**Fig. 2a**).

#### Fitting the null

For every gene and cell type, we fit a single null model using SAIGE-QTL, the same for both subsequent tests (common and rare variant tests alike). Briefly, we fit a Poisson model using the raw counts from the single-cell expression data as the outcome variable (phenotype), and the following as covariates: sex and age of the individuals, 7 genotype PCs, 5 cell type-specific expression PCs, cohort status (TOB or BioHEART), percentage of counts from mitochondrial genes and total counts. We provide all additional parameters and flags as part of the available code in GitHub. We only test genes that are expressed in at least 1% of cells in each cell type. For a subset of gene-cell type combinations the model failed to converge (0.27%) and these were excluded from the analysis (**Supplementary Note 3**).

#### Common variant eQTLs

We test for the effects of common genetic variants on single-cell expression using the single-variant test as implemented in SAIGE-QTL. We consider all SNPs and indels with MAF>=1% and missingness < 15% in our sample. We test for *cis* (variants within a +/-100kb window around the gene body) effects only. We apply a two-step multiple testing correction as recommended in the SAIGE-QTL manuscript^40^, using the Aggregated Cauchy Association Test (ACAT^123^) at gene-level, and Storey’s q-value method (*qvalue* package in R) across genes. At FDR<5%, we identify a total of 154,932 eQTLs in at least one of the 28 cell types, from 17,674 unique genes (83% of all 21,404 genes tested).

The number of eQTLs identified varied substantially across cell types, ranging from 11,491 eQTLs in CD4_TCM_ (85% of all genes tested in this cell type) to 449 eQTLs in innate lymphoid cells (ILC; 3%) as shown in **Fig. 2b**. This was likely explained by differences in the statistical power across cell types, largely due to the number of cells available for each cell type (correlation between log_10_ number of cells and number of eGenes detected = 0.98, **Supplementary Fig. 6**).

The same analysis was run again for all cell types (common variants only) using a +/-1Mb *cis* window around the gene instead. These results are not used for any of the analyses described in the main paper (except **Supplementary Figure 54**), but we made the summary statistics available, see ‘Data Availability’ for details.

#### Rare variant eQTLs

We test for the effects of rare variants on single-cell expression using the set-based test as implemented in SAIGE-QTL. We consider all SNPs and indels with MAF<1% and missingness < 15%. We test for the combined effects of all variants within a *cis* region around the gene (+/-100kb around the gene body). As described in the SAIGE-QTL publication, we consider a burden test and the SKAT test, and use the Cauchy method to combine *p*-values at the gene-level. Similar to the approach used for the common variant results above, we use the *q*-value method and report significant results at FDR<5%.

Overall, we observe 34,376 rare variant signals across all cell types, for 10,005 unique genes after FDR correction using Storey’s *q-*value on the Cauchy combined *p*-values from the two tests (burden test and SKAT). The percentage of genes with a rare-variant combined signal out of all genes tested ranged from 0.09% (ILC) to 39% (CD4_TCM_; **Fig. 2b**). Once again, the tests were well calibrated when shuffling individual IDs (**Fig. 2a**) and the number of cells for each cell type is highly correlated with the number of eGenes identified (**Supplementary Fig. 6**). We discarded results from the ACAT-V test because we found them to be not calibrated in this setting (**Supplementary Note 4**).

An equivalent analysis was performed after restricting the association to only (*cis* rare) variants that were functionally annotated (see details below).

### Single-cell ATAC-sequencing processing

#### Open chromatin status annotation of case examples

Single-cell ATAC-seq data was obtained from a subset of the TOB cohort which includes 30,829 nuclei from 8 individuals [Xue *et al.* 2025]^39^. Sequencing reads were aligned to the Human Genome Reference GRCh38-1.2.0 (gencode v44) using Cell Ranger ATAC (v2.1.0) and the chromatin peaks were called using the built-in MACS2^124^. Cell type labels were annotated based on a CITE-seq reference^42^ with a combined Multiome bridging dataset [Xue *et al.* 2025], consistent with the 28 PBMC cell types annotated for scRNA-seq data. After quality control, 17,099 nuclei were retained to annotate the chromatin peaks in each cell type for the eQTL signals.

#### Open-chromatin status annotation of all rare variants for association set tests

To annotate all rare variants prior to association testing (in the run using only variants in likely-to-be-functional regions), we used the TenK10K scATAC-seq data to annotate variants found to be in open-chromatin regions for the relevant cell type. We first obtained the open chromatin peak list for each of 26 cell types (2 removed due to super rarity; ILC; *n*=33 and CD8_Proliferating_; *n*=4) from [Xue *et al.* 2025]. For each cell type, we re-called peaks from the raw fragment files using the MACS3 tool (v3.0.3) integrated into SnapATAC2 (v2.8.0). The number of peaks identified (*q*-value < 0.05) per cell type ranged from 43,014 in CD4_Proliferating_ to 269,371 in CD16_Mono_, with a median of 181,702. Then we set up a searching function to check if the rare variant is located within any peaks in the corresponding cell type.

### Functional annotation of variants

#### Publicly available resources

We used publicly accessible resources to annotate our variants, including ENCODE^53^ (v3) *cis* candidate regulatory elements (cCREs), UCSC genome annotation database^125^, Activity-by-Contact (ABC)^59^ scores, FANTOM5^126^ enhancer annotations, functional assay results from a deep literature search, chromHMM^127^ predictions, and enhancers from the VISTA Enhancer Database^128^ with positive signal. Any annotations not from genome build GRCh38 were converted using the UCSC LiftOver tool. Any LiftOver segments mapping to non-canonical chromosomes and/or fragmented < 100 bp in GRCh38 were dropped. All overlapping intervals within categories were merged using *bedtools merge*^129^. Detailed annotation information and references are summarised in **Supplementary Table 3**.

#### PromoterAI and SpliceAI to predict promoter and splicing activity of variants

PromoterAI is a convolutional neural network that uses ∼20 kb of sequence context around a promoter variant to predict its effect on gene expression. It is pretrained on histone modifications, DNA accessibility, TF occupancy, and strand-specific CAGE value around TSS at base-pair resolution and fine-tuned with a curated set of rare non-coding promoter variants associated with unusually high or low expression in GTEx v8. PromoterAI scores range from –1 to 1, with negative values indicating a predicted under-expression effect and positive values indicating a predicted over-expression effect. More details on PromoterAI can be found in the publication^46^ and common variant lead eQTLs are annotated with their PromoterAI score (when applicable) in **Supplementary Table 4**.

SpliceAI^49^ was run on all coding tested variants (common and rare) and the scores are available in Zenodo (see Data Availability). AlphaGenome^61^ and Pangolin^62^ were additionally used to analyse the possible mis-splicing outcomes of two variants.

#### Annotations for rare variant set-test analyses

We used an aggregated “functional and open” flag to label any variant sitting in a region considered to be in any of a promoter, enhancer, or actively transcribed region, in a transcription factor binding site (**Supplementary Table 3**), predicted to be splicing-altering or found to reside in an open-chromatin region in the relevant cell type based on our own scATAC-seq (details above).

### Fine-mapping of common variant eQTLs

We ran SuSie^43^ (R package ‘susieR’, v=0.14.2) for fine-mapping. Specifically, we used the ‘susie_rss’ function using L=10, Z scores from the SAIGE-QTL summary statistics, effective N (number of individuals tested, which varied across cell types, see **Fig. 2b**), and correlation matrices we calculated for all *cis* variants for each gene (noting that SAIGE-QTL using a Poisson model violates SuSie’s linear assumption, **Supplementary Note 5**). We considered each credible set as an ‘independent signal’, and we defined as a ‘primary’ signal the credible set containing the variant with the smallest *p*-value, which is the same as the main eQTL for each eGene reported throughout the rest of the manuscript. For genes with more than one independent signal, all other credible sets were defined as ‘non-primary’. The credible set size in **Fig. 2c** refers to the number of variants contained in each credible set. For the comparison between primary and non-primary signals (**Supplementary Fig. 7**) all variants with PIP>0.9 in either the primary credible set or any of the non-primary credible sets for every gene were considered in terms of both their distance from the transcription start site and their functional annotations.

### All cell types pseudobulk analysis

We aggregated expression values by taking the mean value at the level of gene and individual for each cell type. We then used each cell type pseudobulk as a biological replicate and ran SAIGE-QTL (Poisson) using all the same parameters as for the main results presented in the manuscript. In the comparisons to the single-cell cell type-specific analyses (**Supplementary Figures 11** and **32**), only genes tested in the pseudobulk analysis as well were retained (*n*=21,192).

### Cell type specificity analysis

To quantify cell type specificity, for each of the 154,932 identified eQTLs, we first performed pairwise comparisons of association estimates in the primary cell type and in each of the other 27 cell types. We classified these pairwise comparisons into five scenarios based on eQTL association, eGene expression, and direction of allelic effect on gene expression in the two cell types: 1) eGene is expressed only in the primary cell type, 2) eGene is expressed in both cell types, but eQTL identified only in the primary cell type, 3) independent eQTL variants (LD *r*^2^ < 0.5) for the same eGene are identified in each cell type, 4) same eQTL variants or different variants in LD *r*^2^ ≥ 0.5) for the same eGene are identified in each cell type with concordant direction of effect, and 5) same eQTL variants or different variants in LD r2 ≥ 0.5) for the same eGene are identified in each cell type with discordant direction of effect. These scenarios are summarised in the table below:

| Scenario | eQTL detection | eGene expression | Direction of effect on gene expression |
| --- | --- | --- | --- |
| 1 | Found in cell type A only | Expressed in cell type A only | – |
| 2 | Found in cell type A only | Expressed in cell type A & B | – |
| 3 | Independent eQTL variants (LD $r^2 < 0.2$) found in cell type A and B | Expressed in cell type A & B | – |
| 4 | Shared eQTL (same variant or different variants with LD $r^2 \geq 0.2$) found in cell type A and B | Expressed in cell type A & B | Concordant |
| 5 | Shared eQTL found in cell type A and B | Expressed in cell type A & B | Discordant |

Further, we used a pairwise meta-analysis approach for all eQTLs that are initially classified as in *scenario 2* to take into account potential type 2 error due to insufficient statistical power for association analysis in rarer cell types. This was done for each significant eQTL (FDR<5%), by comparing its effect size in the primary cell type to its effect sizes in each of the other cell types. Meta-analysis was performed using a random-effects model with the *metafor* package in R^130^ (v4.0.0). We compared the resulting meta-analysis *p*-value to the primary *p*-value, denoting an empirically chosen threshold of 10^5^-fold reduction (**Supplementary Fig. 47**) as evidence of sub-discovery eQTL signal in cell type 2, in which case the eQTL in question would be reclassified as scenario 4 / 5 depending on concordance of allelic effect direction.

Next, we calculated an aggregate score for each of the 17,674 common variant eGenes by taking the maximum value of the scenarios above across all pairs of cell types for each eGene (results presented in **Figure 4a, bar plot on the right**). The following table described interpretation of the scenario / aggregated values in each of the comparison:

| Scenario / Score | Pairwise comparison (considering two cell types only, A and B) | Aggregated eGene score (aggregated across all 27 other cell types) |
| --- | --- | --- |
| 1 | eQTL found in the cell type A and eGene not expressed in cell type B | eQTL found and eGene expressed only in 1 cell type |
| 2 | eGene expressed in cell type A and B, but eQTL identified in cell type A only | eGene expressed in $\geq 2$ cell types, but eQTL found only in 1 cell type |
| 3 | Independent eQTL variants (LD $r^2 < 0.5$) for the same eGene found in cell type A and B | $\geq 2$ independent eQTL variants (LD $r^2 < 0.5$) for the same eGene found in $\geq 2$ cell types, without evidence of sharedness across cell type pairs |
| 4 | Shared eQTL (same variant or different variants with LD $r^2 \geq 0.5$) found in cell type A and B with concordant allelic effect direction on gene expression | eQTL is shared between $\geq 2$ cell types (LD $r^2 \geq 0.5$), all of which show concordant allelic effect direction |
| 5 | Shared eQTL found in cell type A and B with discordant allelic effect direction on gene expression | eQTL is shared between $\geq 2$ cell types (LD $r^2 \geq 0.5$), and $\geq 2$ cell types show discordant allelic effect direction |

Given that correlation of effect estimates are proportional to LD *r*^2^ between variants, we note that variants in lower LD are more likely to exhibit a discordant allelic effect direction and therefore may result in overestimation of aggregated count for scenario 5. We repeated this approach using the gene-level rare variant association results. For each gene in a given cell type identified at FDR <5% based on the Cauchy-aggregation *p*-value (from burden test and the SKAT test), we compared the estimates for the same gene against those derived from each of the other 27 cell types. Similar to the common variant results above, each pairwise comparison is initially classified into one of the following scenarios: 1) gene is expressed in the primary cell type only, 2) gene is expressed in both cell types, but rare variant association signal is identified only in cell type 1, and 3) rare variant association signal is identified in both cell types. To account for potential misclassification of scenario 2 genes where rare variant association signal is absent due to lack of statistical power in the association test in the secondary cell type, we meta-analysed effect estimates from Burden test results of both cell cell types using a random-effects model. Genes with meta-analysis *p*-value 10^5^-fold smaller than the original meta-analysis were reclassified as *likely shared* eGenes. Unlike the common variant results, we did not perform comparison of direction of effect sizes given that the discovery was done using unsigned *p*-values from Cauchy aggregation of Burden and SKAT tests.

### Statistical colocalisation

We use the COLOC package^131^ to perform statistical colocalisation between our eQTLs and a number of traits and diseases using GWAS summary stats from publicly available sources.

Specifically, we downloaded GWAS summary statistics for the following 16 disease traits: Alzheimer’s disease (AD)^74^, asthma^64^, breast cancer^70^, colorectal cancer^72^, COVID-19^73^, Crohn’s disease (CD)^65^, IgA nephropathy^66^, inflammatory bowel disease (IBD)^65^, lung cancer^71^, malignant lymphoma^50^, multiple sclerosis (MS)^75^, Parkinson’s disease (PD)^76^, prostate cancer^72^, rheumatoid arthritis (RA)^67^, systemic lupus erythematosus (SLE)^68^, type 1 diabetes (T1D)^69^ and ulcerative colitis (UC)^65^.

Additionally, we downloaded UKBB GWAS summary statistics for 44 blood cell count and serum traits^77^ (SNPs and indels only) from (https://gymreklab.com/science/2023/09/08/Margoliash-et-al-paper.html).

Colocalisation was performed using the coloc.abf function from coloc (v5.2.3)^131^ Colocalisation was performed using SNP GWAS summary statistics and common variant eQTLs in our dataset with eQTL type set to ‘quant’ and providing N and MAF rather than sdY as single-cell expression was not normalised and modelled as a Poisson distribution by SAIGE-QTL.

### Cell state eQTL analysis

#### Cell state definition

We computed cell function scores as cell states using scDeepID a multi-task transformer, where a biologically-informed masked-encoder is used to generate cell function embeddings along the supplied database, followed by combining a <cls> token [CLS] for representing each cell’s identity. A set of transformer blocks are then used to learn the high-level data representation of the data, followed by two task heads for cell type identification and cell simulation. The attention-based latent space is extracted from the model by attention rollout^132^, and cell function scores are calculated by attribution methods for deep learning models. scDeepID can be flexibly used with different supplied databases such as Reactome, gene ontology (GO), and more.

To define the cell states in the major NK cells of the TenK10K cohort, we first train our scDeepID model on a publicly available dataset that contains different NK subtypes^94^, then perform label transfer by the trained model on our data. Specifically, we use Scanpy to automatically select ∼600 highly variable genes (min_mean=0.0125, max_mean=3, min_disp=0.5) in the reference dataset, then only keep the shared genes between the reference and TenK10K cohort to train the model (*i.e.,* 560 genes). In particular, we further define NK_intermediate_, NK_adaptive_ and NK_mature_ cells in the NK cell type. Then we train a separate scDeepID model with the three fine-grained cell types as well as NK_CD56bright_ and NK_proliferating_ cells (with 2,315 highly variable genes, selected by Scanpy with min_mean=0.0125, max_mean=3, min_disp=0.5). We use the human GOBP database as a guide to train the scDeepID models above. We then extract the attention-based latent space and normalise each cell function score with a zero mean and unit variance.

#### Cell state eQTL mapping

We use the cell states defined above to test for genotype x cell state interactions. We focus on all eQTLs mapped in any of the NK cells subpopulations (NK, NK_Proliferating_, NK_CD56bright_, *n*=16,759 unique variant-gene pairs) and use lme4^133^ to test the significance of the interaction term *cell state * genotype* for each eQTL and each of the five cell states, while correcting for age, sex, cohort (BioHEART or TOB), total counts as fixed effect covariates, and including an individual term as a random effect.

The alternative model tested was:

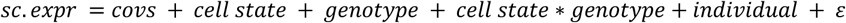

Where *sc.expr* is the single-cell expression profile of the target eGene, *cell state* is one of the cell state functions estimated using scDeepID, *genotype* is the genotype vector at the variant tested, encoded as the number of minor alleles for each individual (0,1,2; expanded such that each cells from a given individual has the genotype of its individual of origin), *individual* is a random effect term to account for the fact that each cell is not an independent observation, and ε is a Gaussian error.

### csaQTL analysis

#### csaQTL mapping

We performed csaQTL associations using GeNA^97^ v1.0.0. We generated single cell objects for each major cell type group and applied the following processing.to each object. Following a standard Scanpy pipeline, we normalised read counts per 10,000, log-transformed normalised counts, and selected approximately 3000 highly variable genes per major cell type (min_mean=0.0125, max_mean=3, min_disp=0.5). The scanpy.pp.regress_out function was used to obtain residualised counts, removing technical variation associated with total counts (library size) and percentage of mitochondrial reads. Expression for each gene was scaled to unit variance across cells. We then created MultiAnnData objects containing the processed counts and donor covariate information. We applied principal component analysis with scanpy using default parameters. Harmony^134^ (harmonypy v0.0.10) was run with default parameters, using sequencing library pools as batch assignment to correct principal components for technical batch effects between sequencing libraries. The harmonised principal components were used to construct a nearest neighbor graph and UMAP using scanpy default parameters.

Next, we generated a neighbourhood abundance matrix (NAM) and its principal components (NAM-PCs) with CNA^135^ (v0.1.6) using default parameters. We then applied GeNA, controlling for age, sex, cohort and, the top seven genotype principal components. We tested associations for all common variants (MAF >= 0.01 and percent missing genotypes < %15). To calibrate the model, we also ran GeNA on permuted genotypes generated by random label swapping (one permutation performed per cell type). Based on this permutation testing, we observed poor calibration for variants below 0.05 MAF, and removed them from the analysis. We considered variants passing the genome-wide significance threshold (*p*-value < 5×10^−8^) as statistically significant. Lead SNPs were defined using the clumping method as described in the GeNA publication^97^. Briefly, at each locus the most significant SNP was considered the lead SNP, and all SNPs with LD > 0.8 to the lead SNP or within a 1 MB window were considered LD-SNPs.We considered the number of unique lead-SNPs for csaQTL loci as the total independent csaQTLs discovered. To visualise cell counts adjusted for covariates, we calculated the fraction of cells within each major subtype belonging to each minor subtype. Using these cell fractions, we fit a linear model with the lm function in R, including age, sex, cohort, and the first 7 genotype principal components as covariates, and visualised the residuals from the fitted model.

#### Characterisation of csaQTL-associated cell states

To characterise the cell state phenotypes detected by GeNA, we used methods previously described by Rumker *et al.*^97^ briefly, we calculated the Pearson correlation between GeNA neighborhood-level phenotypes and expression of highly variable genes. Highly variable genes were then ranked by Pearson’s *r* for each csaQTL and used as input to GSEA. The fgsea (v1.32.2) R package^136^ was used to run GSEA for all Molecular Signatures Database hallmark gene sets. Gene sets with BH-adjusted *p*-value < 0.05 were considered statistically significant. To visualise the expression of enriched gene sets, module scores were calculated with the *scanpy score_genes* function, and overlayed on to UMAP plots.

#### Colocalisation analysis between csaQTLs and blood cell count GWAS

To obtain GeNA summary statistics compatible with colocalisation analysis, we calculated fixed-phenotype summary statistics for each significant lead csaQTL using methods described by Rumker *et al.*^97^. Colocalisation was run with the coloc.abf function from the COLOC R package^131^, using default parameters. We provided COLOC with these fixed-phenotype summary statistics, as well as GWAS from two blood cell count and serum trait GWAS studies. We used the UKBB GWAS summary statistics for 44 blood cell count and serum traits described above, supplemented with an additional 26 blood trait GWAS performed by Vuckovic *et al.*^103^. Colocalisations with PP4 > 0.8 were considered significant.

## Acknowledgements

We acknowledge Vladislav Savelyev and Leonhard Gruenschloss for contributions in prototyping the whole-genome sequencing production pipeline, and Garvan Genomics Platform for providing sequencing services and technical support. We acknowledge Michael Geaghan from the Garvan Data Science Platform for support. The costs of WGS for most samples in this program were supported by the Centre for Population Genomics. Illumina provided discounted reagents for scRNA-seq, and 10X Genomics provided discounted reagents for single cell capture.

## Author contributions

A.S.E.C., J.E.P., and D.G.M. designed the study. A.S.E.C. designed and performed the single-cell processing and eQTL mapping pipelines and led the writing of the manuscript. E.S. and C.B. designed and performed the single-cell RNA sequencing experiments. H.A.T. first established the colocalisation pipeline. A.X. and O.A.D. ran the colocalisation analysis. A.S.E.C. performed the fine-mapping analysis. A.S.E.C. and B.B. processed the single-cell RNA sequencing data. B.B. designed and performed the cell state abundance QTL analysis. A.S.E.C, A.H. and J.E.P. designed and performed the cell type specificity analysis. H.L.H. designed and applied the scDeepID tool for cell state identification. H.L.H., A.S.E.C and J.E.P. designed and performed the cell state-specific eQTL mapping. A.X. contributed scATAC-seq to support annotations of selected variants. S.J.B. provided interpretation for the splicing rare variant examples. J.M.P. and C.W. provided guidance around the fine-mapping and colocalisation analyses. A.X., A.H., H.A.T., K.M.dL. contributed to downstream interpretation. W.Z. provided support for running SAIGE-QTL. M.J.W, H.A.T., M.F., M.H., M.S., A.S., J.M. and V.B provided support with the eQTL mapping pipeline. A.S.L. contributed a curated list of regulatory annotations used to annotate variants. K.W., O.T., M.P.G., S.M.G., and T.N. assisted with participant recruitment and sample collection. M.H., K.B., M.S. and K.M.dL. processed the whole genome sequencing data. B.S.M. and C.U. developed the ethics protocol for the study and coordinated the generation of WGS data. A.M. and M.S. processed the metadata needed to process the WGS data. D.R.N. assisted with the scRNA-seq sample demultiplexing. Z.Q., O.M.S, and H.R.N. contributed to the study design. K.M.dL. and J.P. led the processing of whole genome sequencing data. E.B.D., L.C., and K.K.H.F. designed and performed the annotation of variants using PromoterAI and SpliceAI. A.W.H. designed the Tasmanian Ophthalmic Biobank study. G.A.F. designed the BioHEART study.

All authors contributed to editing the manuscript. J.E.P and D.G.M conceived the project and supervised all aspects of the work.

## Data availability

Individual-level data (genotypes and scRNA-seq count matrices) were deposited to EGA (upload underway). All summary statistics, fine-mapping and colocalisation results were uploaded to Zenodo (10.5281/zenodo.17474113).

## Code availability

All scripts to reproduce the analyses presented here are available in the following GitHub repositories:

● Single-cell expression processing and downstream analyses: https://github.com/powellgenomicslab/tenk10k_phase1/
● WGS data processing: https://github.com/populationgenomics/production-pipelines/tree/main/cpg_workflows/large_cohort
● Processing scripts and association testing pipeline using SAIGE-QTL: https://github.com/populationgenomics/saige-tenk10k
● scDeepID model: https://github.com/powellgenomicslab/scDeepID

