## Supplementary Information for "Impact of rare and common genetic variation on cell type-specific gene expression in human blood"

**Supplementary Figures**……………………………………………………………….…………….4

**Supplementary Figure 1:** Summary of cell, donor and variant QC.
**Supplementary Figure 2:** Number of cells per individual.
**Supplementary Figure 3:** scRNA-seq cell type marker gene expression.
**Supplementary Figure 4:** WGS finds more rare variants than SNP array data.
**Supplementary Figure 5**: Manhattan plots for all 28 cell types (common variants).
**Supplementary Figure 6**: Relationship between number of cells available and number of eGenes identified in each cell type.
**Supplementary Figure 7:** Characterising primary and non-primary signals from fine-mapping.
**Supplementary Figure 8:** Comparison with eQTLGen.
**Supplementary Figure 9:** Comparison with sorted-cell bulk eQTL studies.
**Supplementary Figure 10**: Enrichment of common variant eQTLs that were not found in OneK1K genes with lower and more cell type specific expression.
**Supplementary Figure 11:** Number of eGenes identified by all-cell-type pseudobulk compared to single-cell results stratified by cell type.
**Supplementary Figure 12**: eQTL effect size sign is concordant with PromoterAI scores for the corresponding variant-gene pairs.
**Supplementary Figure 13**: Manhattan plots for all 28 cell types (rare variants).
**Supplementary Figure 14**: Characterisation of common and rare eGenes.
**Supplementary Figure 15:** Rare eGenes being more likely to be expressed in all cell types is likely related to higher overall expression.
**Supplementary Figure 16:** Scatter plots of SKAT and burden *p*-values when testing all variants compared to functional only variants
**Supplementary Figure 17:** Number of eGenes from SKAT and burden tests when testing all variants compared to functional only variants
**Supplementary Figure 18**: Open chromatin peaks around example rare variant signals for *SH2D2A* and *PRDM4.*
**Supplementary Figure 19**: Rare variant eGene example: *PRDM4.*
**Supplementary Figure 20**: Rare variant eGene example: 19:51367458:C:T affecting splicing of *CLDND2*.
**Supplementary Figure 21:** The overlap in number of genes tested across cell types.
**Supplementary Figure 22:** Cell type specificity patterns of common eQTL variants.
**Supplementary Figure 23:** eGenes expression pattern across cell types.
**Supplementary Figure 24:** Distribution of shared eGene across cell types
**Supplementary Figure 25:** Number of eGenes by cell type specificity scenario when excluding rare cell types.
**Supplementary Figure 26:** Correlation of eQTL effect across groups of cell types.
**Supplementary Figure 27:** Pairwise Pearson's correlation of $-\log_{10}(p\text{-value})$ eQTL association between major groups of cell types.
**Supplementary Figure 28**: Cell type specificity patterns of rare variant eGenes.
**Supplementary Figure 29**: Cell type specificity of colocalisations for selected diseases.
**Supplementary Figure 30**: Cell type specificity of colocalisation events.
**Supplementary Figure 31:** Cell type specificity of eGenes with evidence of colocalisation.
**Supplementary Figure 32:** Number of eGenes with at least one colocalisation event found using an all-cell-type pseudobulk compared to single-cell results stratified by cell type.
**Supplementary Figure 33**: Colocalisation between Alzheimer's disease and *IGHG4* in memory B cells.
**Supplementary Figure 34**: Colocalisation between inflammatory bowel disease and *TNFSF15* in pDC.
**Supplementary Figure 35**: More colocalisation event examples.
**Supplementary Figure 36:** Relative marker expression of three representative sub-NK cell types.
**Supplementary Figure 37**: Cell state eQTL tests are calibrated.
**Supplementary Figure 38**: Overlap of cell state eQTL mapping in NK cells across cell functions.

**Supplementary Figure 39:** NK dynamic eQTL example: *RPS26* eQTL is modulated by the cell killing function.
**Supplementary Figure 40:** Cell state abundance QTL Manhattan plots.
**Supplementary Figure 41:** Cell state abundance QTL QQ plots.
**Supplementary Figure 42:** Cell state abundance QTL QQ plots MAF>=5%.
**Supplementary Figure 43:** Validation of csaQTLs from GeNA original publication.
**Supplementary Figure 44:** Supplementary info about *LYZ* csaQTL.
**Supplementary Figure 45:** Characterisation of csaQTL-associated cell states with gene set enrichment analysis.
**Supplementary Figure 46:** Monocyte csaQTL example: *CLEC12A*.
**Supplementary Figure 47:** Shared and cell type specific eQTLs using different meta-analysis p-value to the primary p-value delta thresholds.
**Supplementary Figure 48:** Principal component analysis (PCA) of TenK10K Phase 1 cohort samples overlaid with global reference populations.
**Supplementary Figure 49**: Experimental workflow.
**Supplementary Figure 50**: Number of cells identified by CellRanger.
**Supplementary Figure 51**: Number of cells identified as doublets or unassigned.
**Supplementary Figure 52**: Cell typing concordance between methods.
**Supplementary Figure 53**: Cell QC metric distributions.
**Supplementary Figure 54.** Window size comparison 100kb vs 1Mb.
**Supplementary Figure 55:** Cellbender-Cellranger mismatch in the estimated number of cells.
**Supplementary Figure 56.** Cellbender background fraction distributions.
**Supplementary Figure 57:** Inflated *p*-values when regressing out covariates.
**Supplementary Figure 58:** Inflated *p*-values for ACAT-V results.
**Supplementary Figure 59:** Fewer eGenes identified by SuSie.

### Supplementary Tables……………………………………………………………………….47

*All supplementary tables are provided as supplementary files.*

**Supplementary Table 1**: Donor summary.
**Supplementary Table 2**: Summary of immune cell types identified.
**Supplementary Table 3:** Summary of functional annotations.
**upplementary Table 4**: List of common variant single-cell eQTL (FDR<5%) with cell type specificity scenario information included.
**Supplementary Table 5**: List of rare variant single-cell eQTL (FDR<5%) with cell type specificity scenario information included.
**Supplementary Table 6:** List of colocalization events at PP4>0.8.
**Supplementary Table 7.** Replication of OneK1K csaQTLs in TenK10K phase 1.

### Supplementary Notes………..…………………………………………………………….47

**Supplementary Note 1**: Monocyte proportion differences across cohorts.
**Supplementary Note 2:** Cellbender tool limitations with high-throughput scRNA-seq.
**Supplementary Note 3.** Convergence fails of SAIGE-QTL.
**Supplementary Note 4.** ACAT-V is not calibrated in our data.
**Supplementary Note 5.** SuSie limitations with SAIGE-QTL outputs.

Supplementary Figures

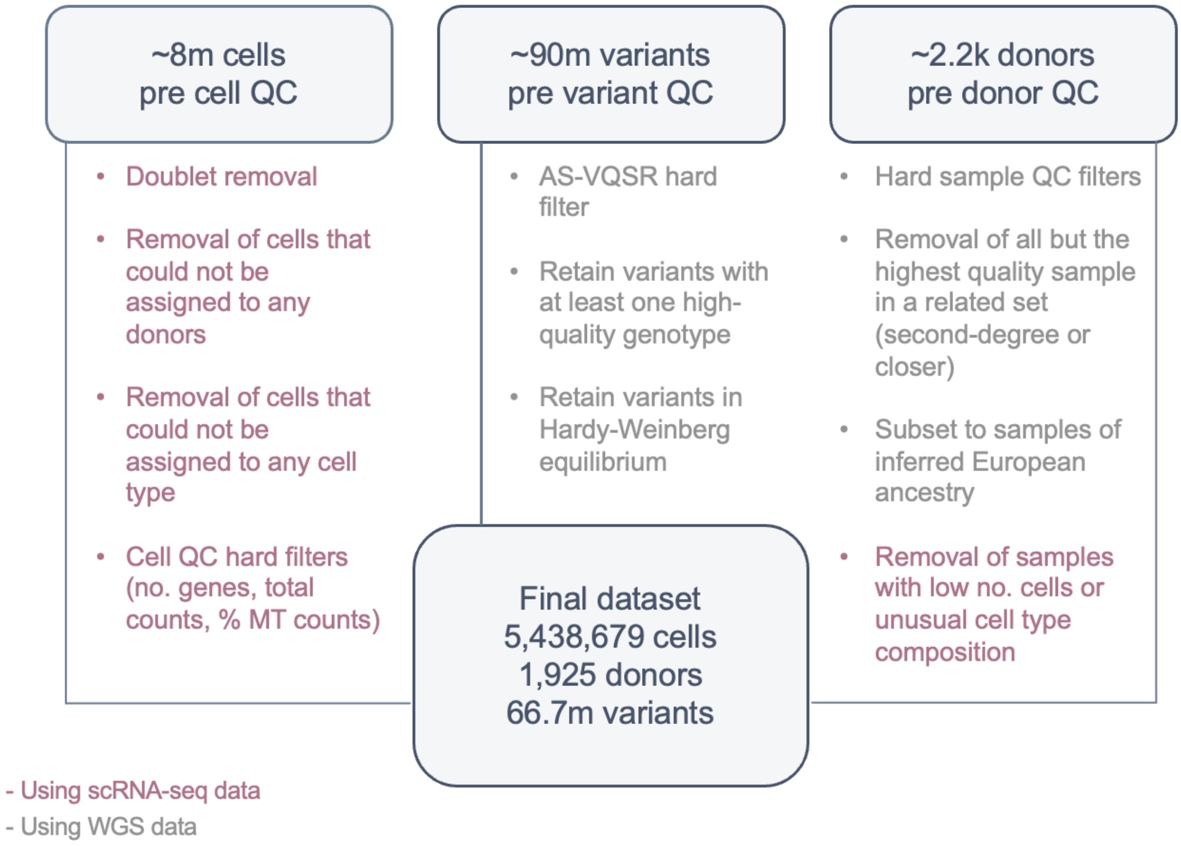


**Supplementary Figure 1. Summary of cell, donor and variant QC.** Pink are QC steps applied based on scRNA-seq data, grey using whole-genome sequencing (WGS) data. **Left)** Cell QC. Doublets were classified using majority voting of three tools (vireo, scDblFinder and scds) and removed. Cells were also excluded if they could not confidently be assigned to any individuals by vireo. We removed all cells that could not confidently be mapped to any of the 28 cell types using our hierarchical scPred implementation. Finally, we applied hard filters on additional cell QC metrics: cells were removed if they had counts from less than 1,000 and more than 10,000 genes, less than 800 total counts, or more than 20% counts from mitochondrial reads. **Middle)** Variant QC. Variants were retained if they passed the Allele-Specific Variant Quality Score Recalibration (AS-VQSR) filter. AS-VQSR is a machine learning technique trained on a control set that filters out probable artefacts from a given callset. Additionally, variants were retained if they had at least one high-quality genotype (GQ≥20, DP≥10, allele balance > 0.2 for heterozygotes) and if there was no evidence for an excess of heterozygotes at the site compared to Hardy-Weinberg expectations (InbreedingCoeff ≥ -0.3). **Right)** Donor QC. Individuals were excluded based on the following hard QC filters: high rates of contamination (FREEMIX ≥ 5%); an elevated percentage of chimeras (PCT_CHIMERAS ≥ 6.3%); median insert size < 250bp; and sex aneuploidy or discordance between reported and observed sex. Additionally, for family sets of second-degree or closer relatives, the sample with higher quality based on WGS + scRNA-seq data was retained. For association testing, only samples with genetic similarity to those of known European ancestry (as identified via PCA analysis on the genotypes) were retained. Finally, any individuals with less than 100 QC-passing cells or with extremely unusual cell type composition based on scRNA-seq were also excluded. **Bottom)** Overall numbers post all three layers of QC are reported in the bottom panel. More detail can be found in Methods.

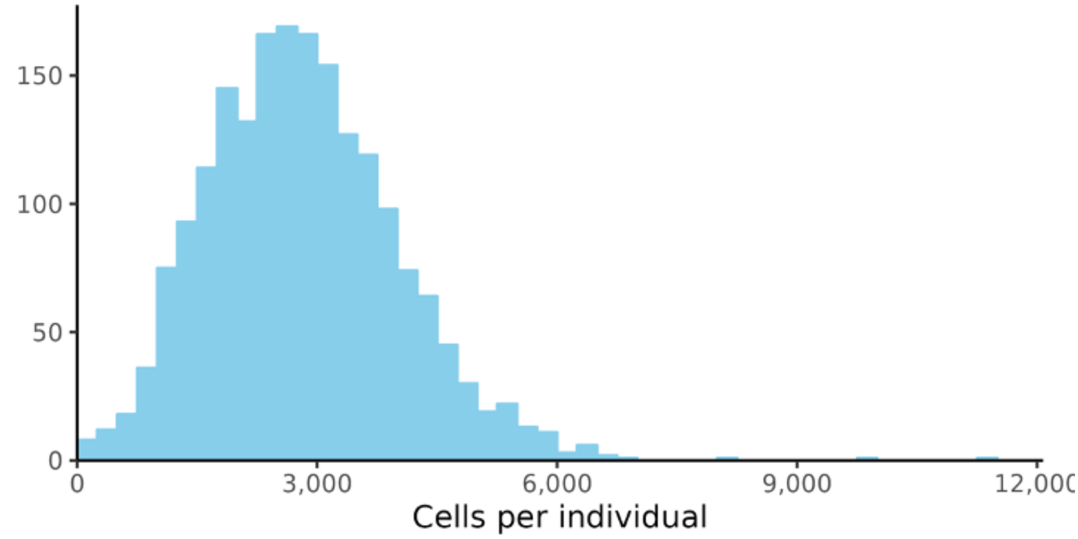


**Supplementary Figure 2. Number of cells per individual.** Distribution of the number of cells per donor after scRNA-seq, demultiplexing, and quality-control filtering (mean=2,825, range=124-11,348).

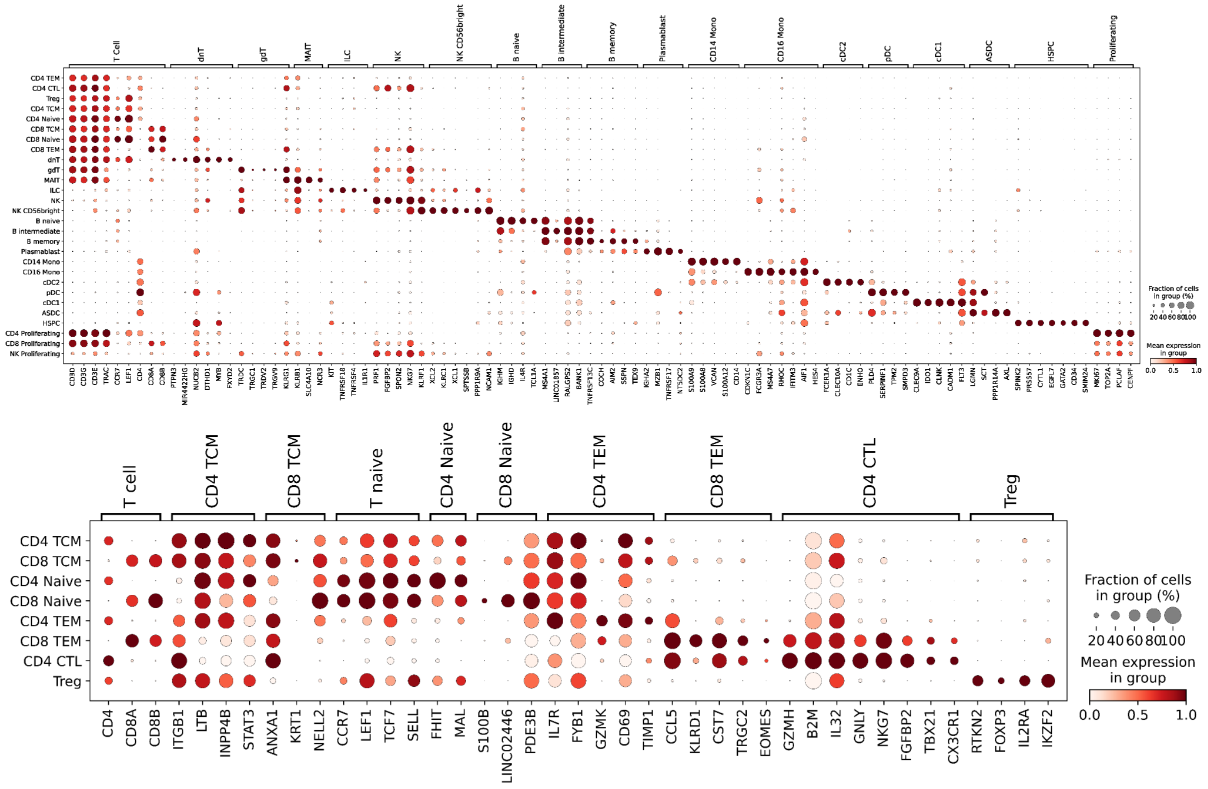


**Supplementary Figure 3. scRNA-seq cell type marker gene expression. Top.** Dot plot of scRNA-seq expression for canonical immune marker genes. Dot size corresponds to the percentage of cells in the cell type expressing the gene and dot colour indicates the mean expression within each cell type. **Bottom.** Dot plot of scRNA-seq expression for canonical T cell marker genes. Similar to a but focussing on CD4+ and CD8+ T cell subpopulations.

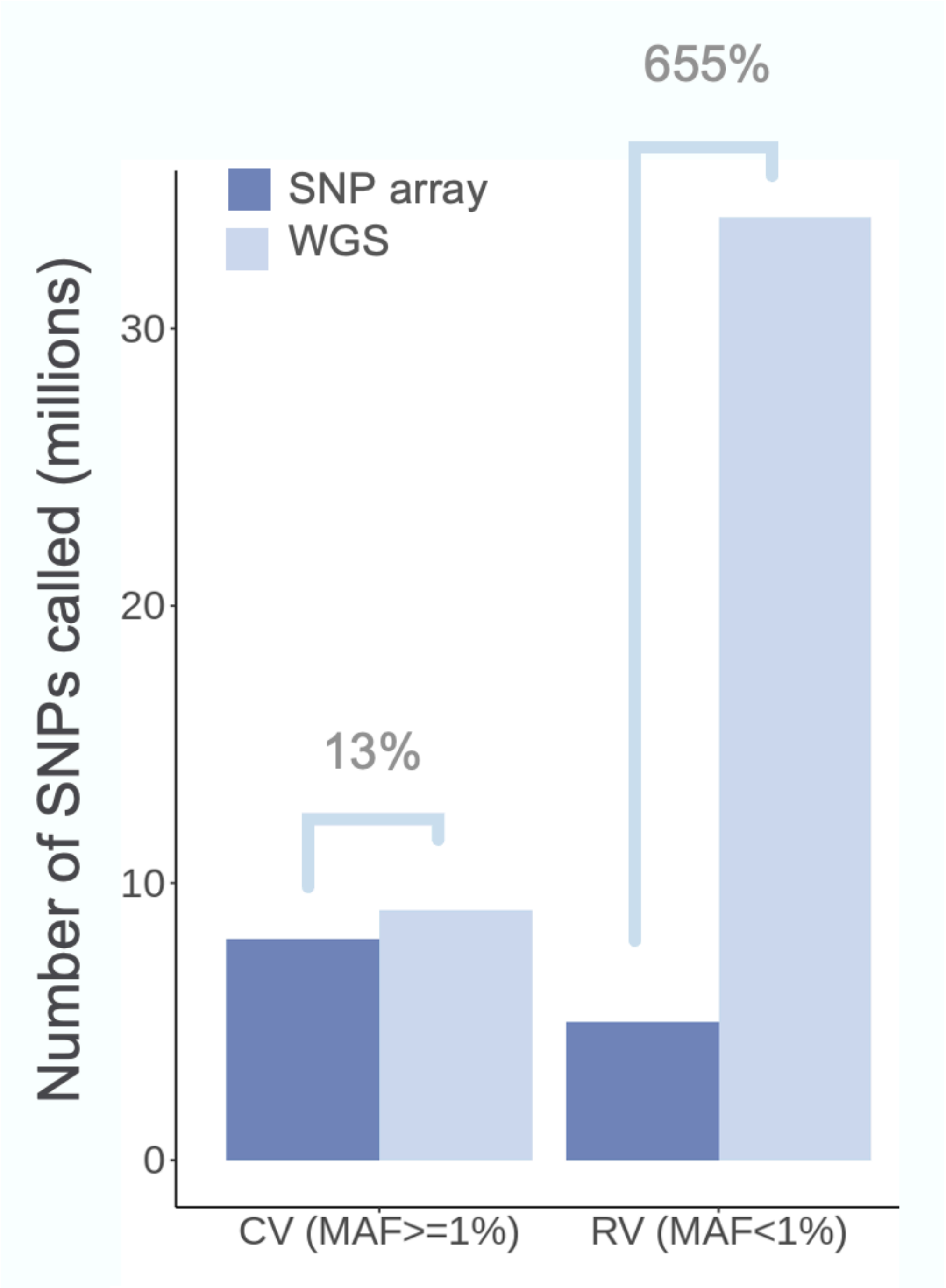


**Supplementary Figure 4. WGS finds more rare variants than SNP array data.** For a subset of 960 individuals, SNP array data and WGS were available. Bar plots representing the number of biallelic SNPs called using the two protocols, split by frequency groups. CV: common variant (minor allele frequency; MAF≥1%), RV: rare variant (MAF<1%).

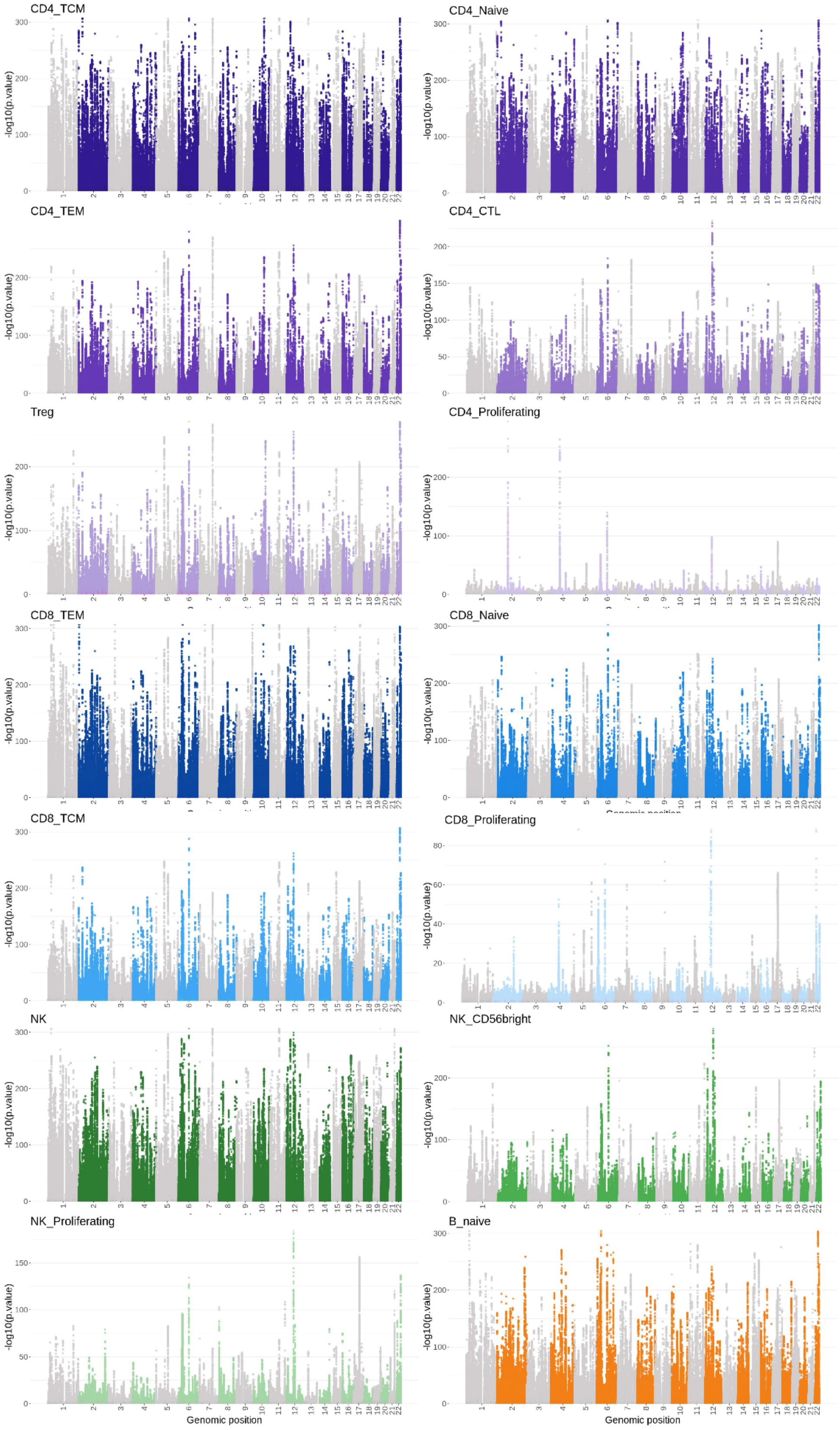


**Supplementary Figure 5. Manhattan plots for all 28 cell types (common variants; part 1)** Manhattan plots for common variant eQTLs (single-variant test results) for all 28 cell types. All variants tested for any of the genes are included.

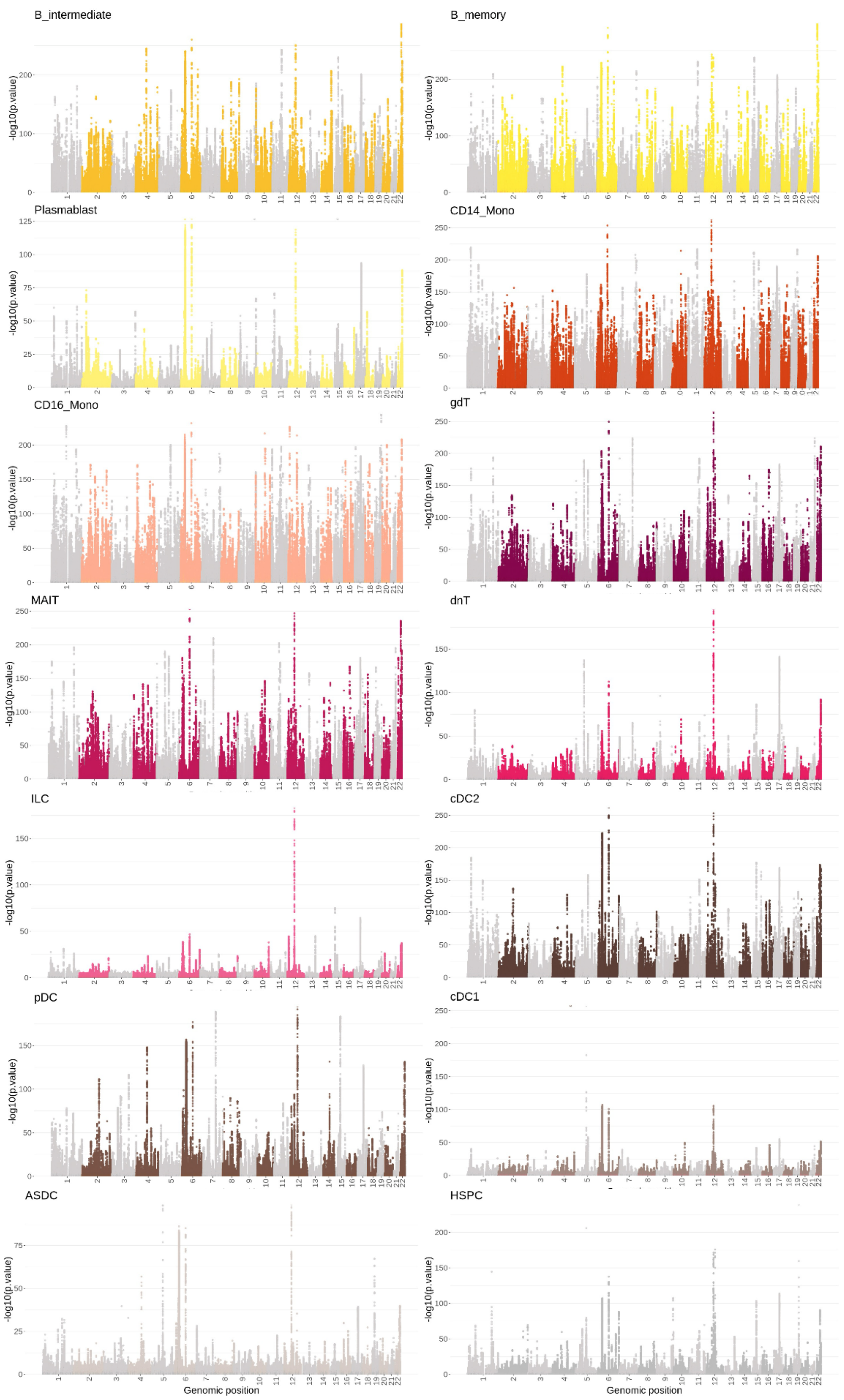


**Supplementary Figure 5. Manhattan plots for all 28 cell types (common variants; part 2)** Manhattan plots for common variant eQTLs (single-variant test results) for all 28 cell types. All variants tested for any of the genes are included.

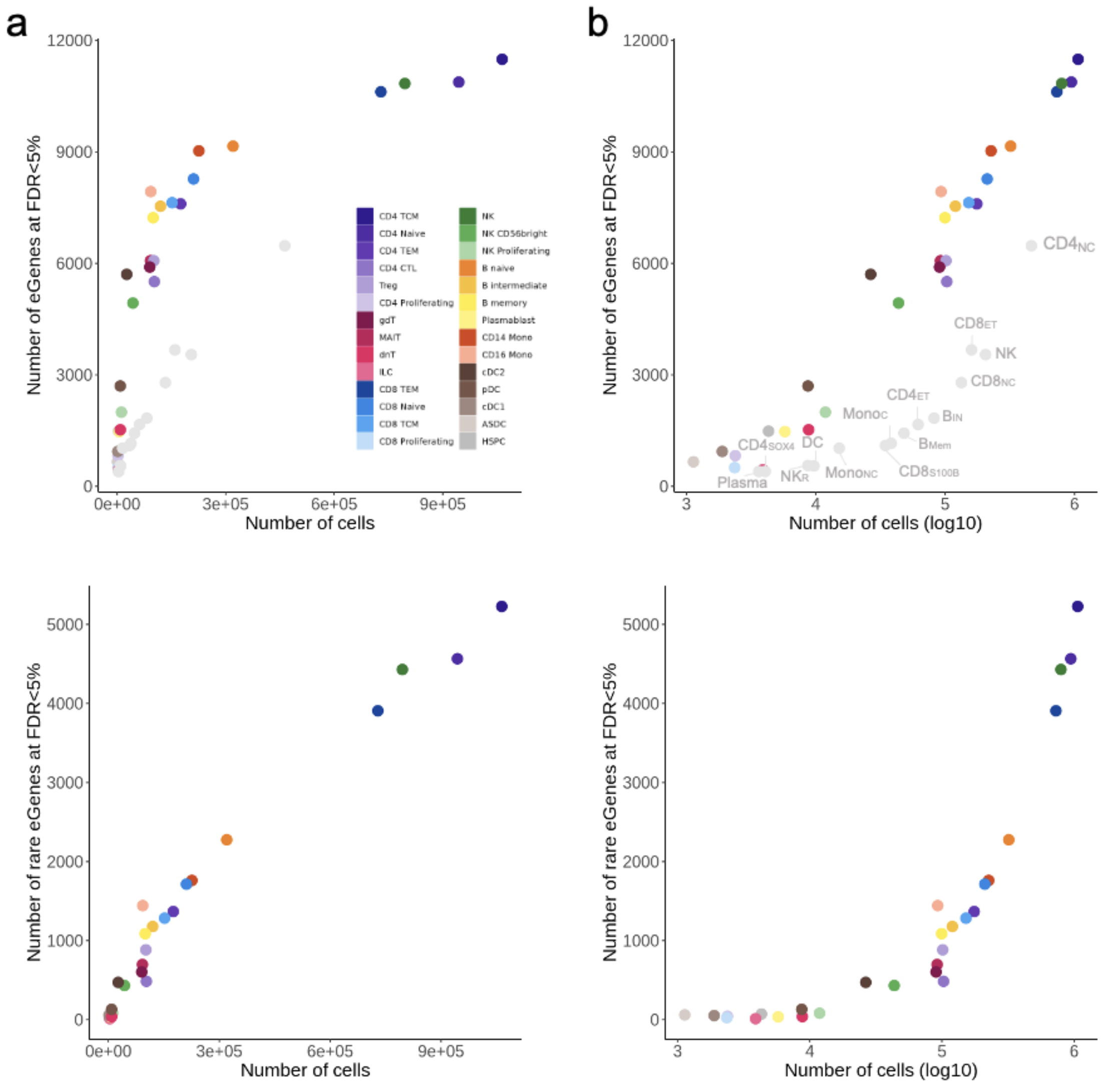


**Supplementary Figure 6. Relationship between number of cells available and number of eGenes identified in each cell type.** Before **(a)** and after **(b)** $log_{10}$ transformation for both common (top) and rare (bottom) variant tests, all coloured by cell type (legend for all in the top left plot). eGene = gene with at least one eQTL (common variant test) or a significant gene-level combined rare signal (rare variant test), in both cases at FDR < 5%. For the common variant plots (top), equivalent dots are included for OneK1K[1] results for comparison (in grey, cell types from the original Yazar *et al.,* 2022 annotated in the right-hand plot).

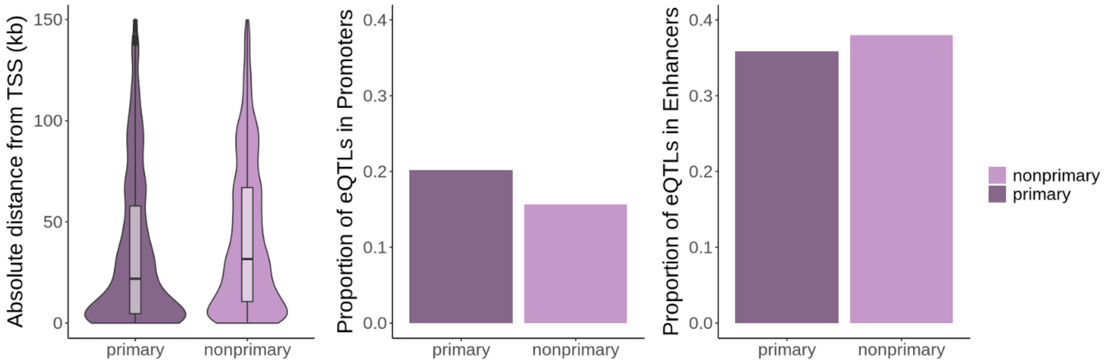


**Supplementary Figure 7. Characterising primary and non-primary signals from fine-mapping.** For all genes with more than one independent signal identified confidently by fine-mapping (PIP>0.9, using SuSie[2], more detail in Methods) signals (=credible sets) were categorised as either primary (including the top most significant variant) or non-primary. Non-primary signals are on average found further away from the gene's transcription start site (TSS), less likely to be found in promoter regions, and more likely to be found in enhancer regions compared to primary signals.

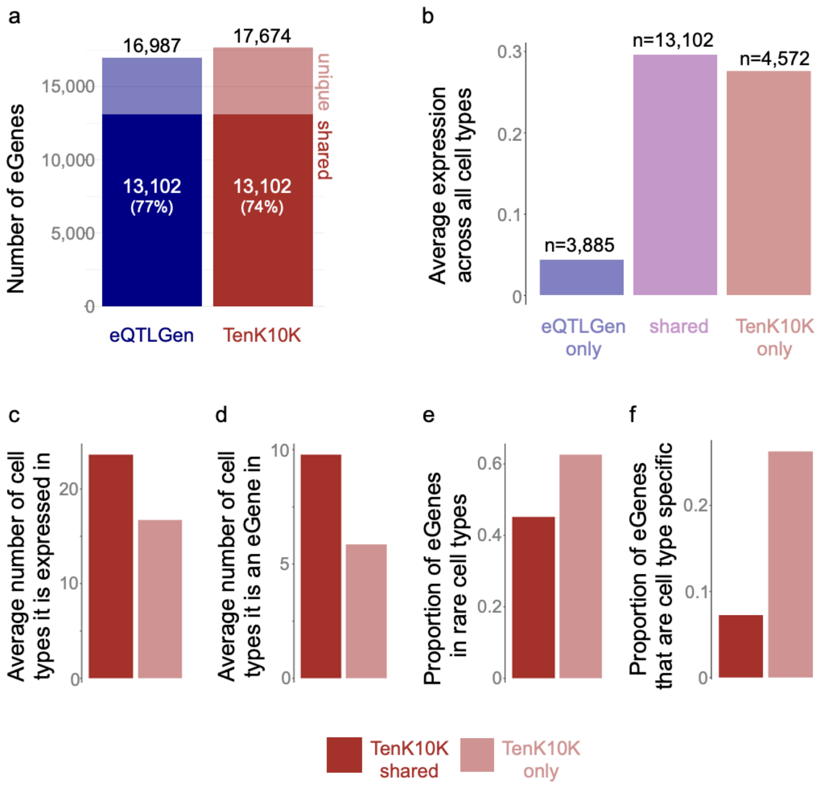


**Supplementary Figure 8. Comparison with eQTLGen. a)** The number of unique eGenes reported at FDR<5% by eQTLGen[3] or identified at FDR<5% in any of the 28 cell types in TenK10K phase 1 are reported, and further split by whether they are found in the other study or not (solid colour:shared, faded:unique). **b)** Characterisation of the shared and unique eGenes from a, in terms of their overall expression levels across all cell types in TenK10K. The eGenes found uniquely by eQTLGen (missed by TenK10K) are expressed at very low levels likely explaining why we lack the power to identify them. **c-f)** Characterisation of eGenes found uniquely in TenK0K (missed by eQTLGen). They are **c)** expressed and **d)** identified as eGenes in fewer cell types; **e)** more likely to be found as eGenes in a rare cell type ($n < 6{,}000$) and **f)** more likely to be cell type specific eGenes.

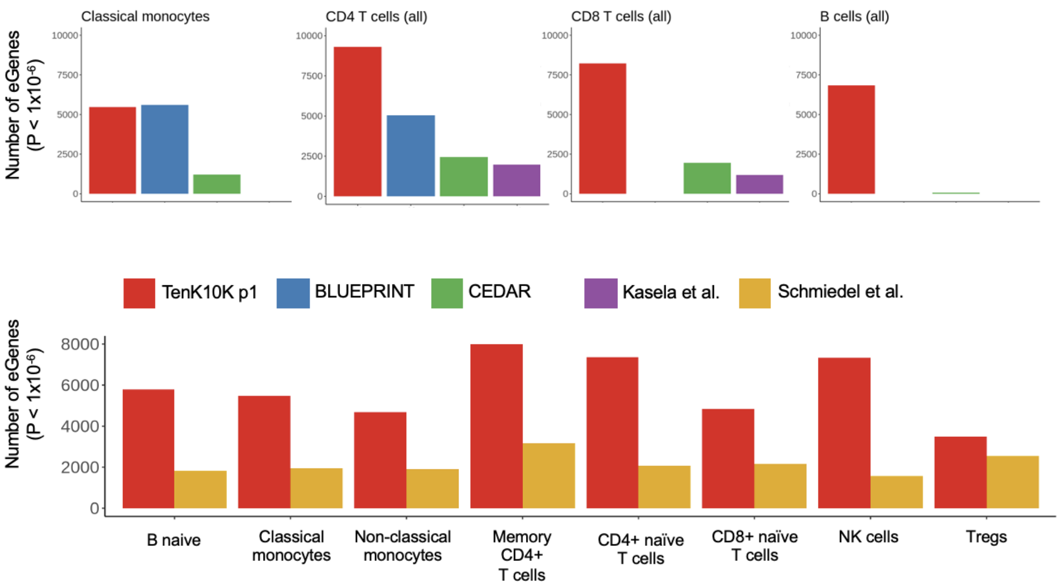


**Supplementary Figure 9. Comparison with sorted-cell bulk eQTL studies.** We compared the number of unique eGenes found at the conservative threshold of $p$-value<$10^{-6}$ in TenK10K and other bulk eQTL studies using sorted immune cells, comparing like-for-like cell types. Top row: CD4 T cells (all) included all eGenes found in at least one of $CD4_{CTL}$, $CD4_{Naive}$, $CD4_{Proliferating}$, $CD4_{TCM}$, $CD4_{TEM}$ and $T_{reg}$ cells; CD8 T cells (all) included all eGenes found in at least one of $CD8_{Naive,}$ $CD8_{TCM}$, $CD8_{TEM}$, $CD8_{Proliferating}$; B cells (all): $B_{naive}$, $B_{intermediate}$, $B_{memory}$, Plasmablast. Bottom row: Memory CD4+ T cells were either $CD4_{TCM}$ or $CD4_{TEM}$ in TenK10K, and any of Tfh, Th1-17, Th1, Th17, Th2from Schmiedel *et al*[4]; Treg were both naive and memory Treg from Schmiedel *et al*. Maximum sample sizes are $n$=197 for BLUEPRINT[5], $n$=323 for CEDAR[6], $n$=313 for Kasela *et al*[7], $n$=91 for Schmiedel *et al*, and 1,925 for TenK10K, with some variation from cell type to cell type.

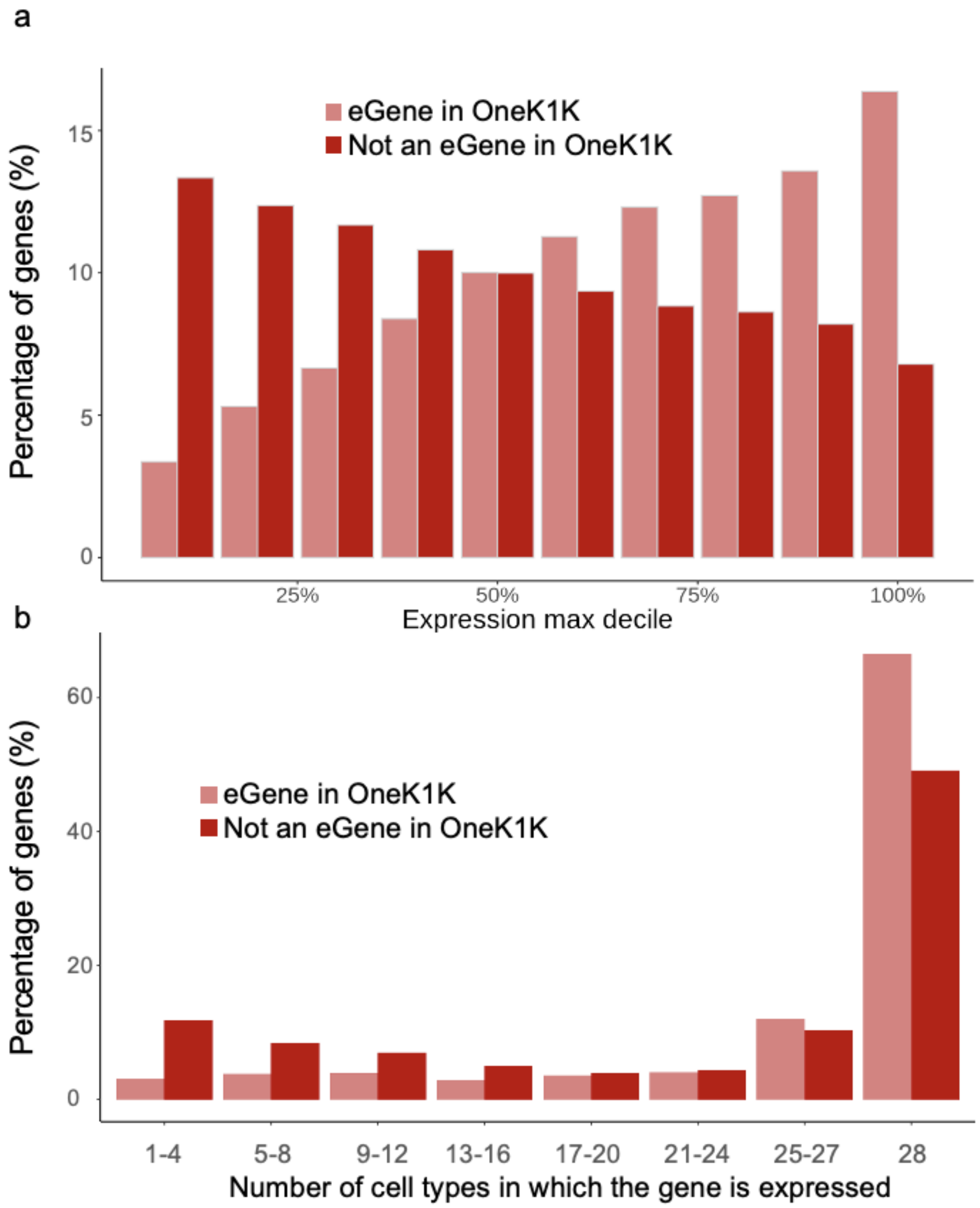


**Supplementary Figure 10. Enrichment of common variant eQTLs that were not found in OneK1K in genes with lower and more cell type specific expression. a)** Common variant eGenes from this study that are found or not found to be eGenes in the OneK1K study[1], stratified by max expression across cell types (in our data). eGenes novel to this study are more lowly-expressed. **b)** Common variant eGenes from this study that are found or not found to be eGenes in the OneK1K study, stratified by the number of cell types in which they are sufficiently expressed (>1% of all cells). eGenes novel to this study are expressed in a more cell type specific manner when considering the list of eGenes identified here and the list of eGenes reported by Yazar *et al.*, and then comparing their expression across the 28 cell types in the TenK10K phase 1 dataset (not OneK1K).

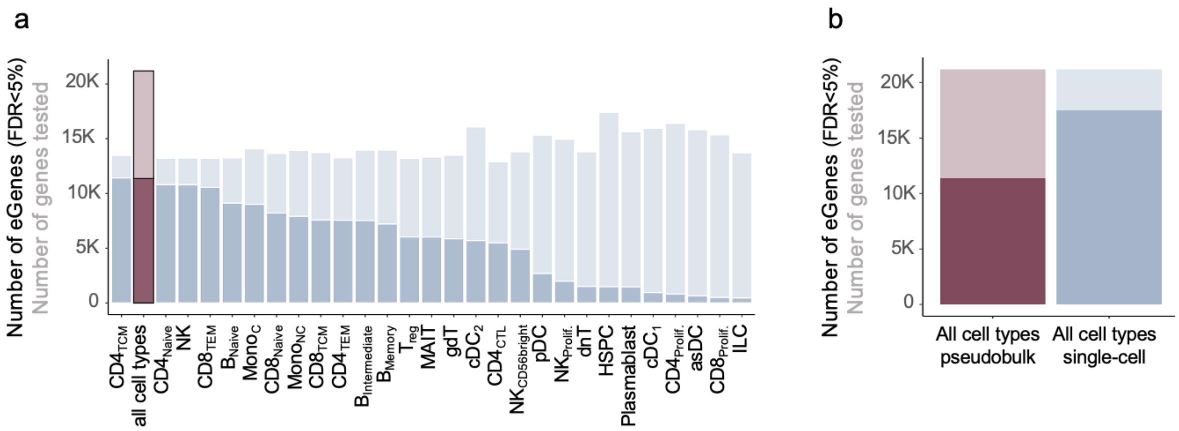


**Supplementary Figure 11. Number of eGenes identified by all-cell-type pseudobulk compared to single-cell results stratified by cell type. a)** Number of eGenes (FDR<5%, solid colour) out of all genes tested (faded colour) identified in each cell type separately (main set of results described in this paper; blue), and in an artificial pseudobulk approach combining all cells from all cell types (red): many more genes could be tested, and the number of eGenes is comparable with the most abundant cell types. **b)** When combined across all cell types, 53.6% of genes tested were identified by the pseudobulk approach as opposed to 82.3% which were found as eGenes in at least one of 28 cell types.

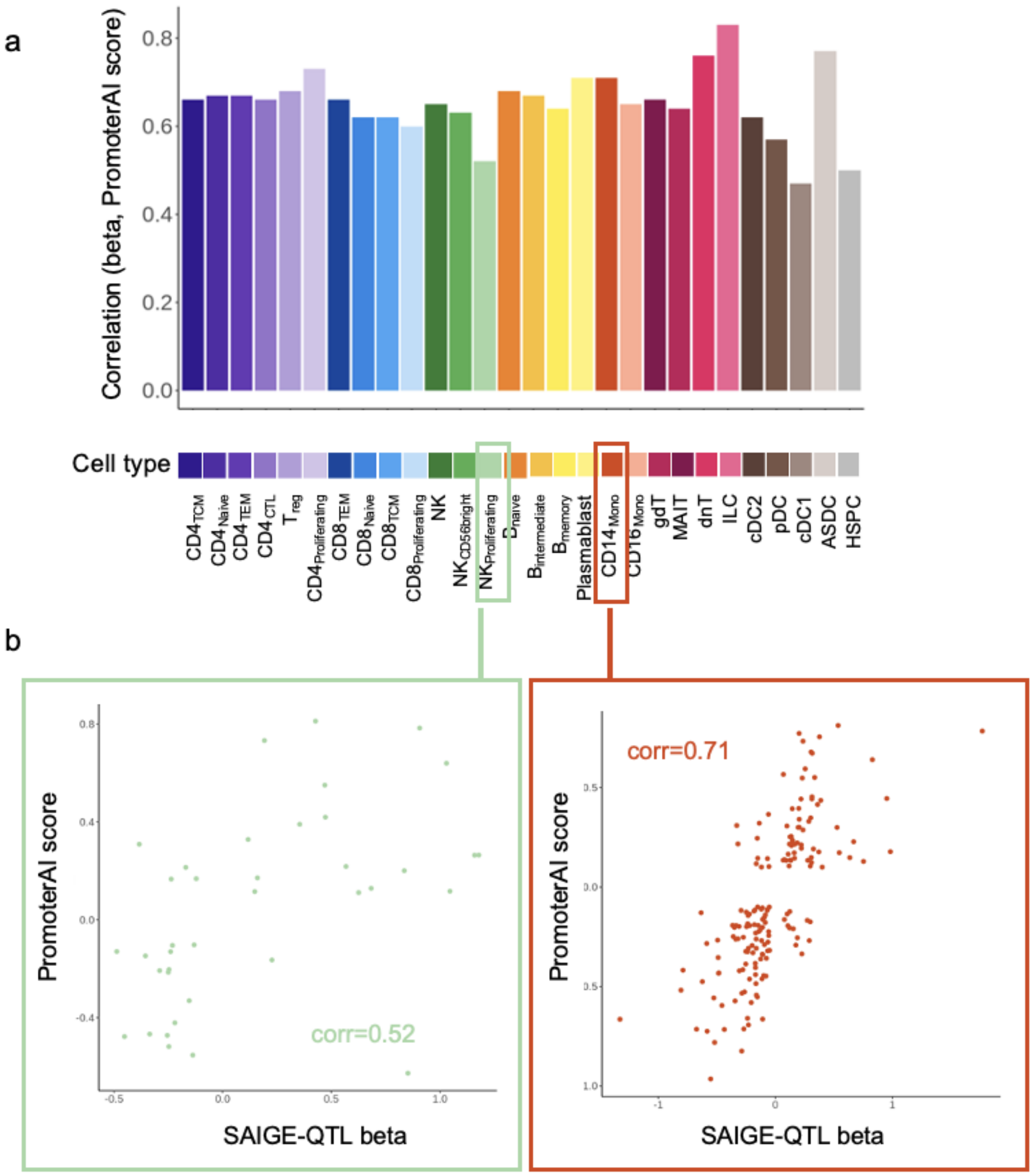


**Supplementary Figure 12. eQTL effect size sign is concordant with PromoterAI scores for the corresponding variant-gene pairs. a)** Bar plot of correlations between SAIGE-QTL effect sizes and PromoterAI[8] scores for each cell type, coloured by cell type. Only significant variants (*p*-value < $5x10^{-8}$) and variants with some evidence of promoter activity (absolute PromoterAI score > 0.1) were included. **b)** Scatter plot of SAIGE-QTL effect sizes and PromoterAI scores for two example cell types ($NK_{Proliferating}$ and $CD14_{Mono}$).

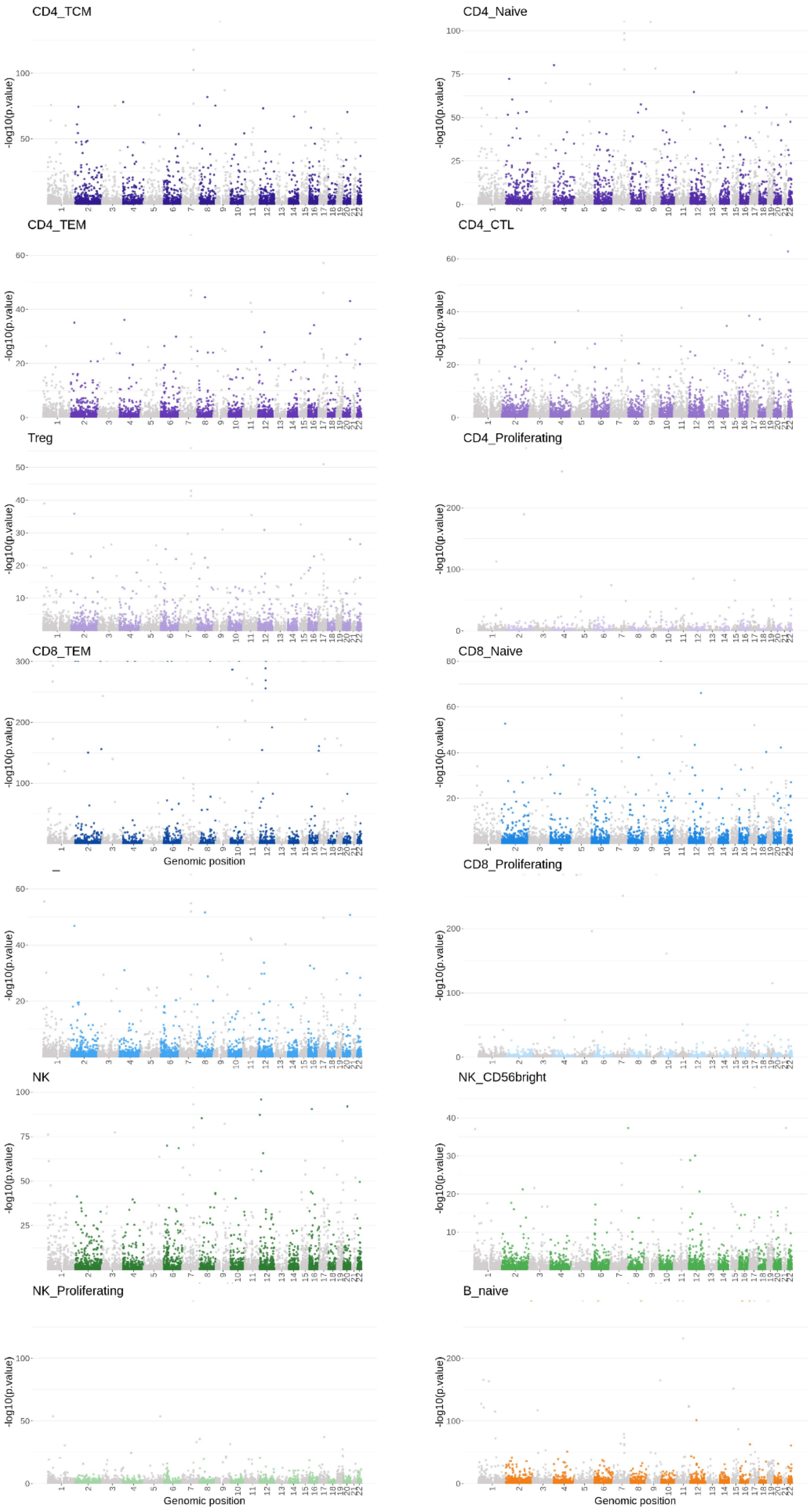


**Supplementary Figure 13. Manhattan plots for all 28 cell types (rare variants; part 1)** Manhattan plots for rare variant eQTLs (gene-level set test results) for all 28 cell types. All genes tested are included.

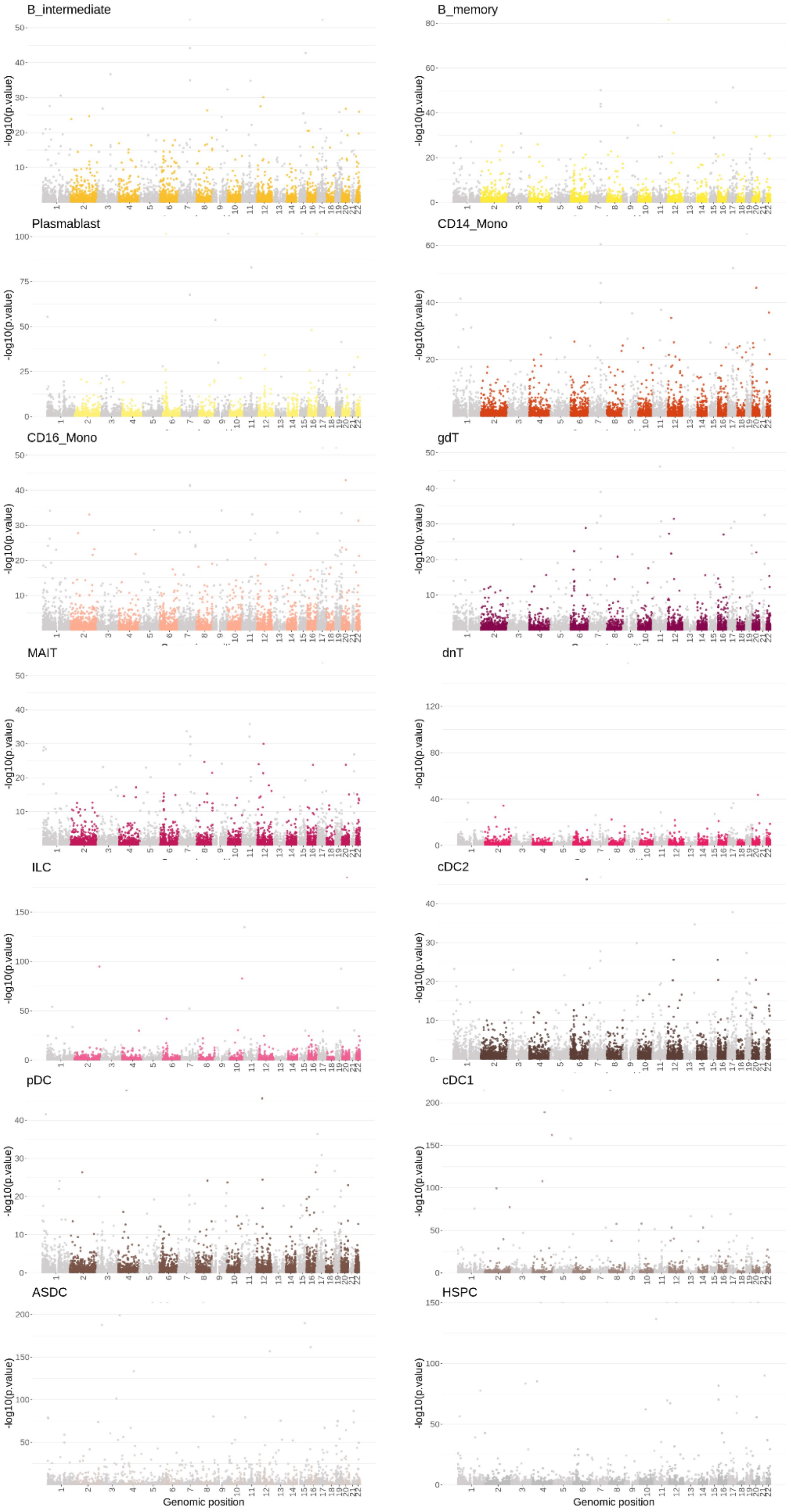


**Supplementary Figure 13. Manhattan plots for all 28 cell types (rare variants; part 2)** Manhattan plots for rare variant eQTLs (gene-level set test results) for all 28 cell types. All genes tested are included.

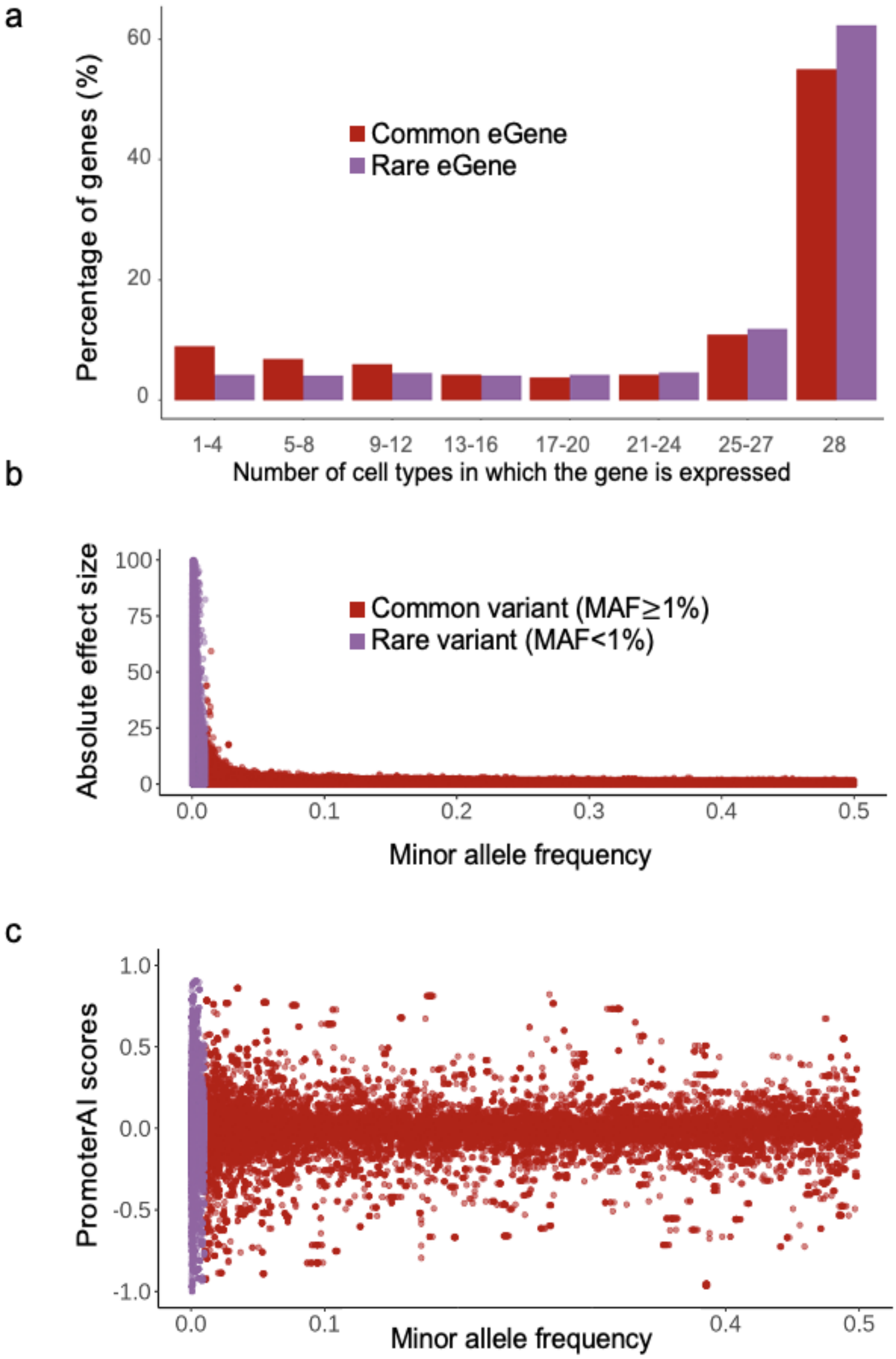


**Supplementary Figure 14. Characterisation of common and rare eGenes. a)** The percentage of rare eGenes (FDR<5%, $n$=10,005; purple) and common eGenes ($n$=17,674; red) in each decile of cell type specific expression (number of cell types a gene is sufficiently expressed in / 28). Rare eGenes are enriched for more ubiquitously expressed genes whereas common-only eGenes are more cell type specific. **b)** Single variant tests for both common (red, MAF>=1%) and rare (purple, MAF<1%) variants, quantifying the effect size as the absolute value of beta against the minor allele frequency (MAF, in $\log_{10}$ scale). The $y$-axis is capped at 100. Note that for rare variants, shown here are single variant test results, as opposed to the main set-based test results (burden, SKAT), reported and shown in **Fig. 2**. **c)** Strength of variants as measured by PromoterAI[8] are enriched for rare variants (common in red, rare in purple).

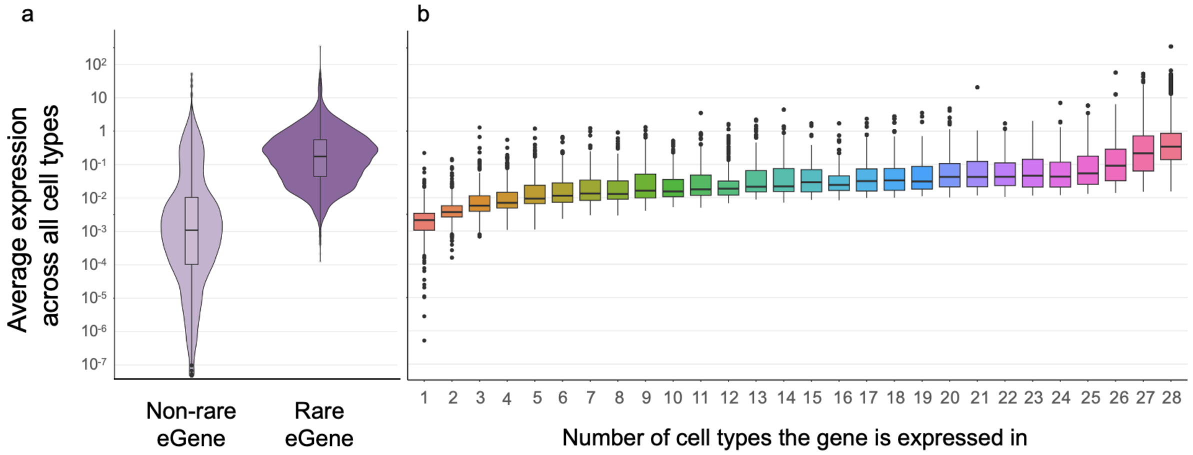


**Supplementary Figure 15. Rare eGenes being more likely to be expressed in all cell types is likely related to higher overall expression. a)** Rare eGenes are on average more highly expressed (average expression across all 28 cell types). **b)** Number of cell types each gene is expressed in as a function of average expression across cell types.

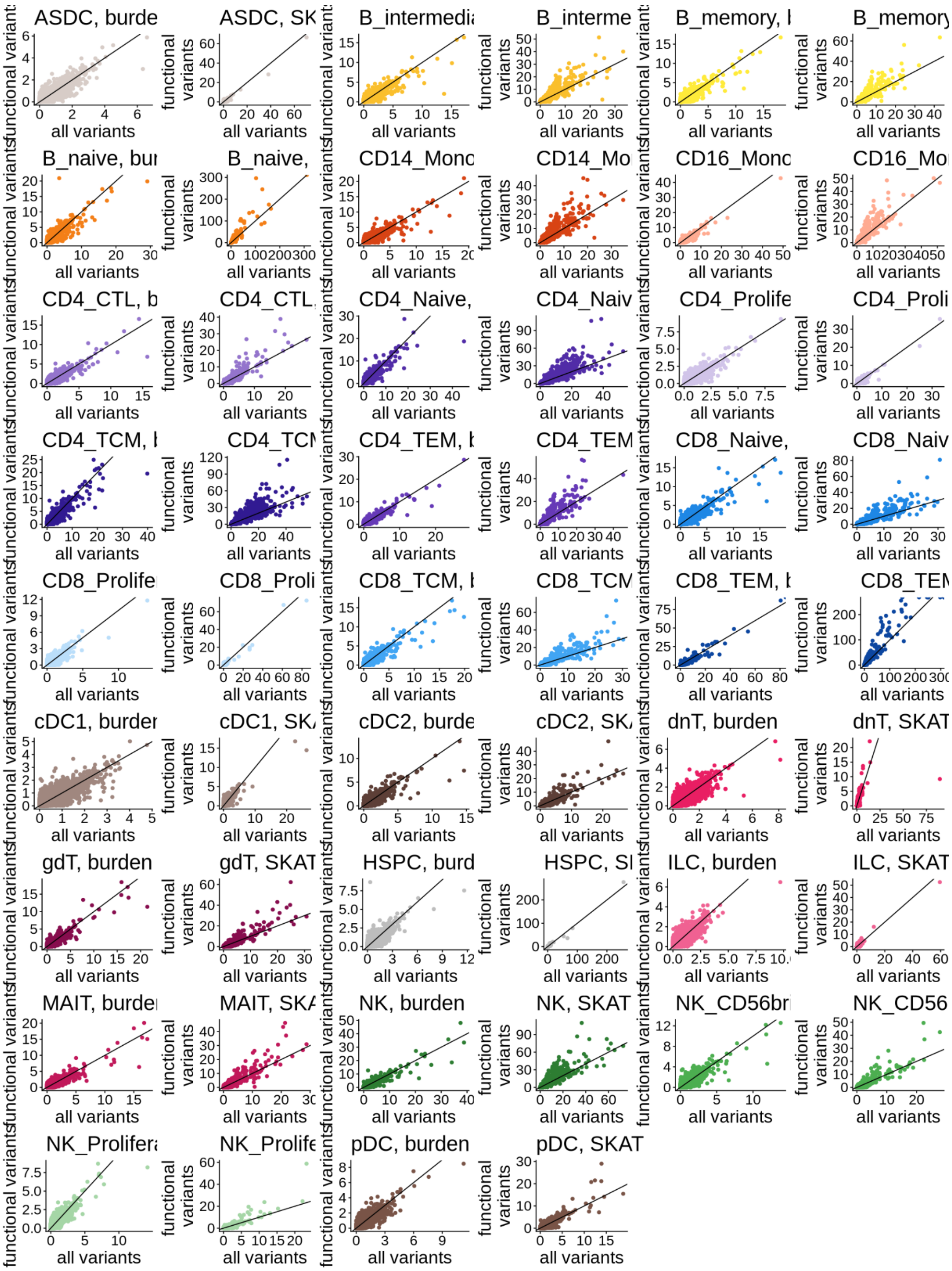


**Supplementary Figure 16. Scatter plots of SKAT and burden *p*-values when testing all variants compared to functional only variants.** Scale is $-\log_{10}$(*p*-values), and all variant results are on the x axis, functional variants on the y. Dots are coloured by cell type. Weights used: dTSS.

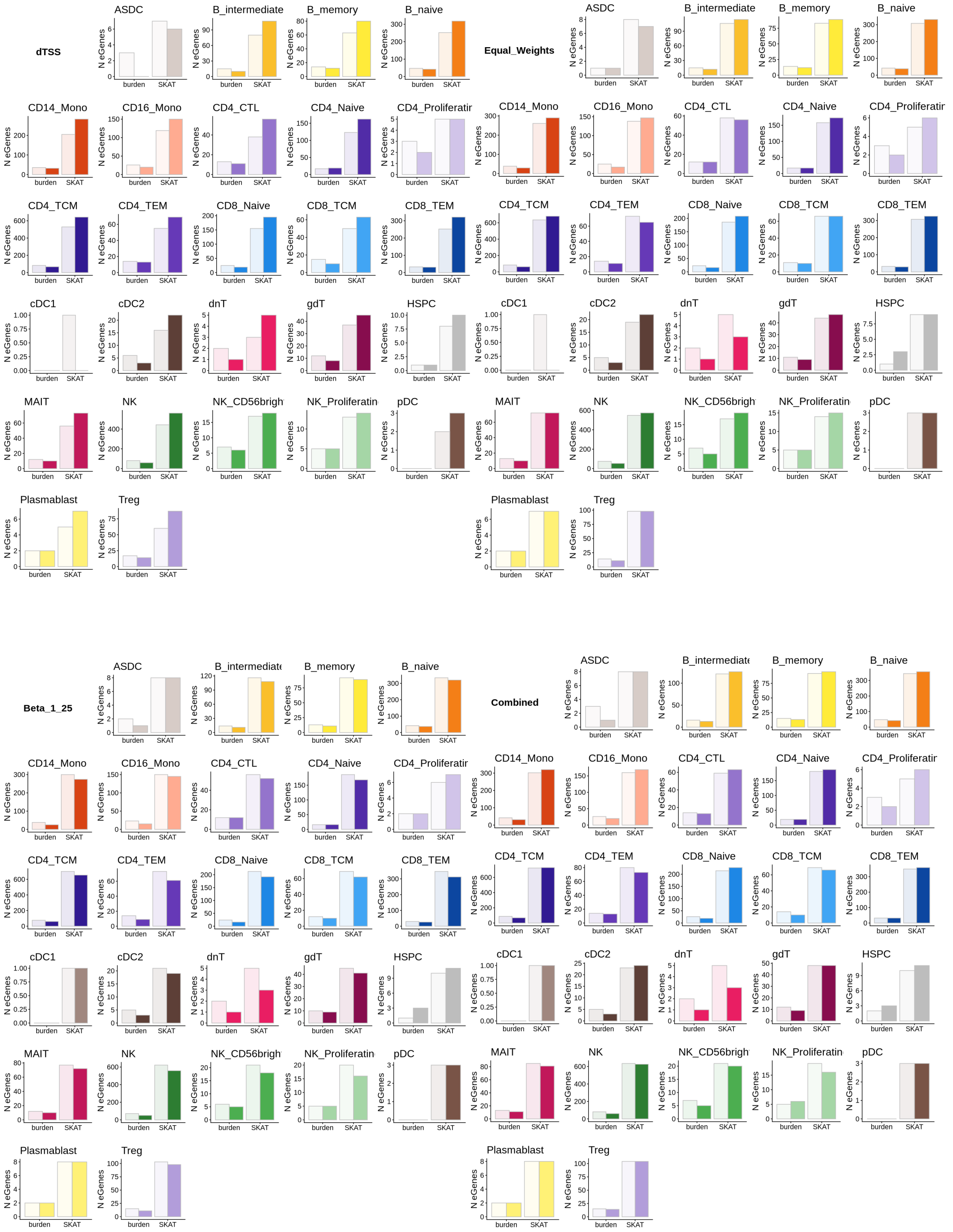


**Supplementary Figure 17. Number of eGenes from SKAT and burden tests when testing all variants compared to functional only variants.** Similar to 3c but with different weight settings, dTSS (top left), equal weights (top right), Beta(1,25) (bottom left), all combined (bottom right).

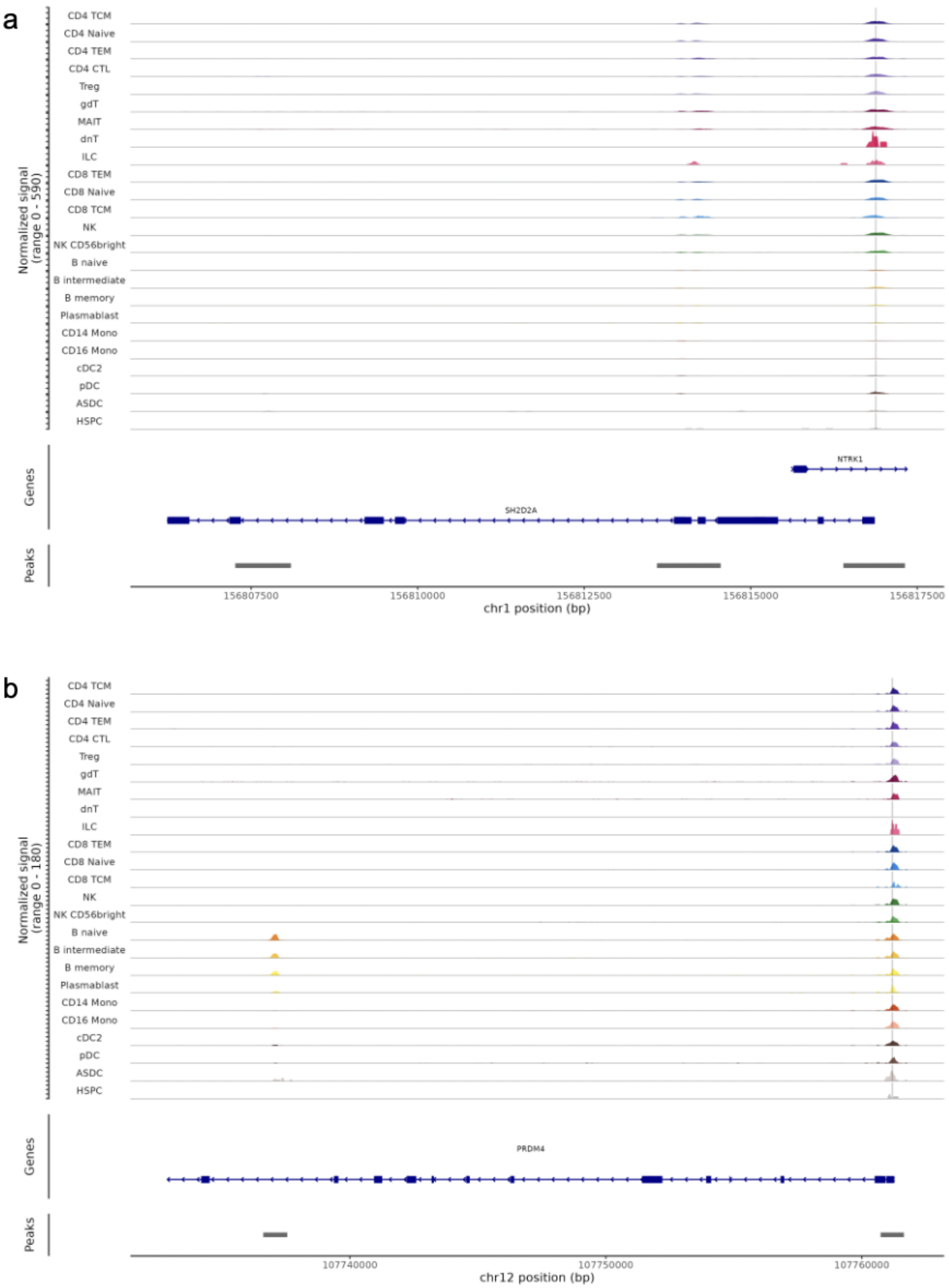


**Supplementary Figure 18. Open chromatin peaks around example rare variant signals for *SH2D2A* and *PRDM4*.** Similar to **Fig. 3** panel **c** and **d**, but for all cell types. Top tracks: scATAC–seq for each cell type in the region surrounding each gene, coloured by cell type. Middle track: gene(s) in the area. Bottom track: peak(s) location. The variants' locations are marked by a vertical line (1:156816831:A:AC and 12:107761146:T:G).

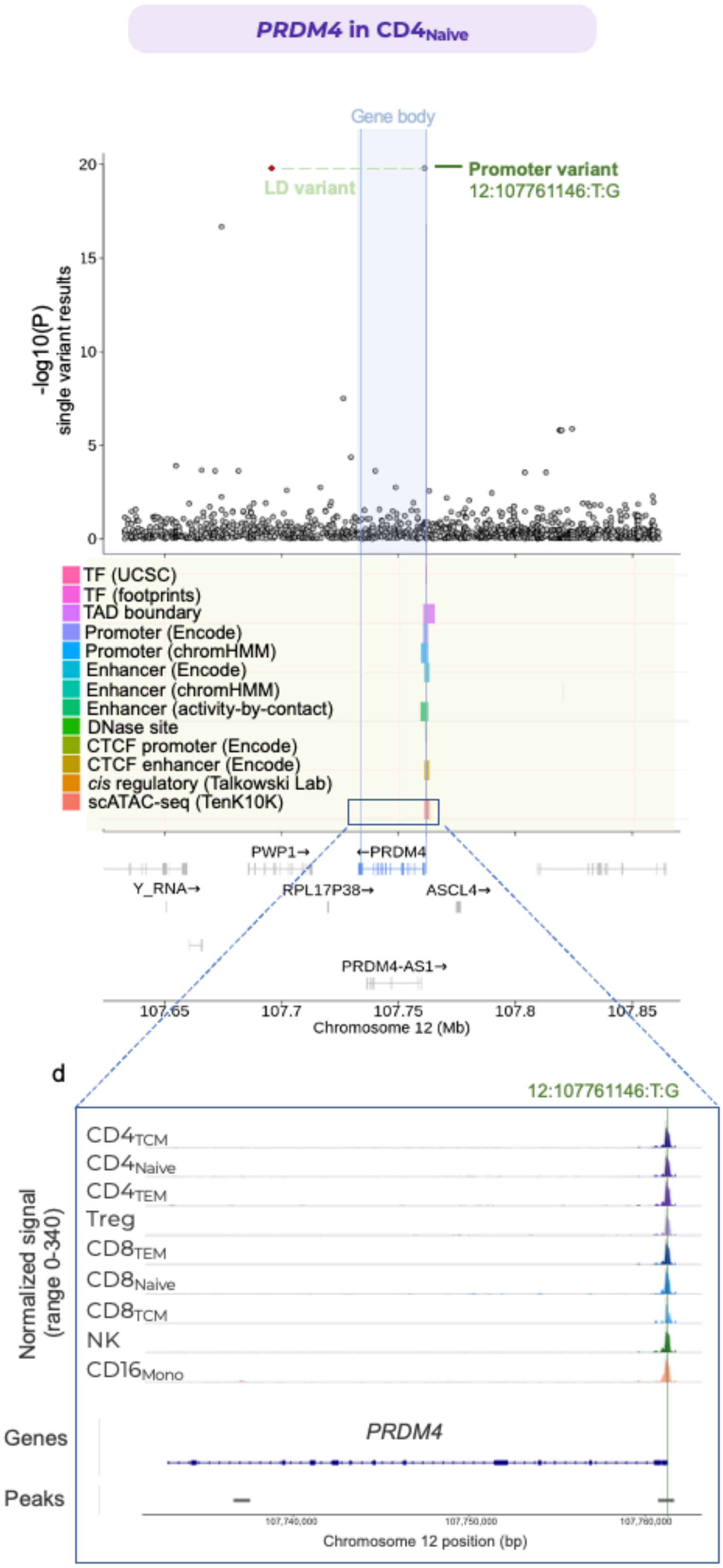


**Supplementary Figure 19. Rare variant eGene example: *PRDM4*.** Locus zoom plot of rare genetic variants associated with the *PRDM4* gene on chromosome 12 in $CD4_{Naive}$ cells.

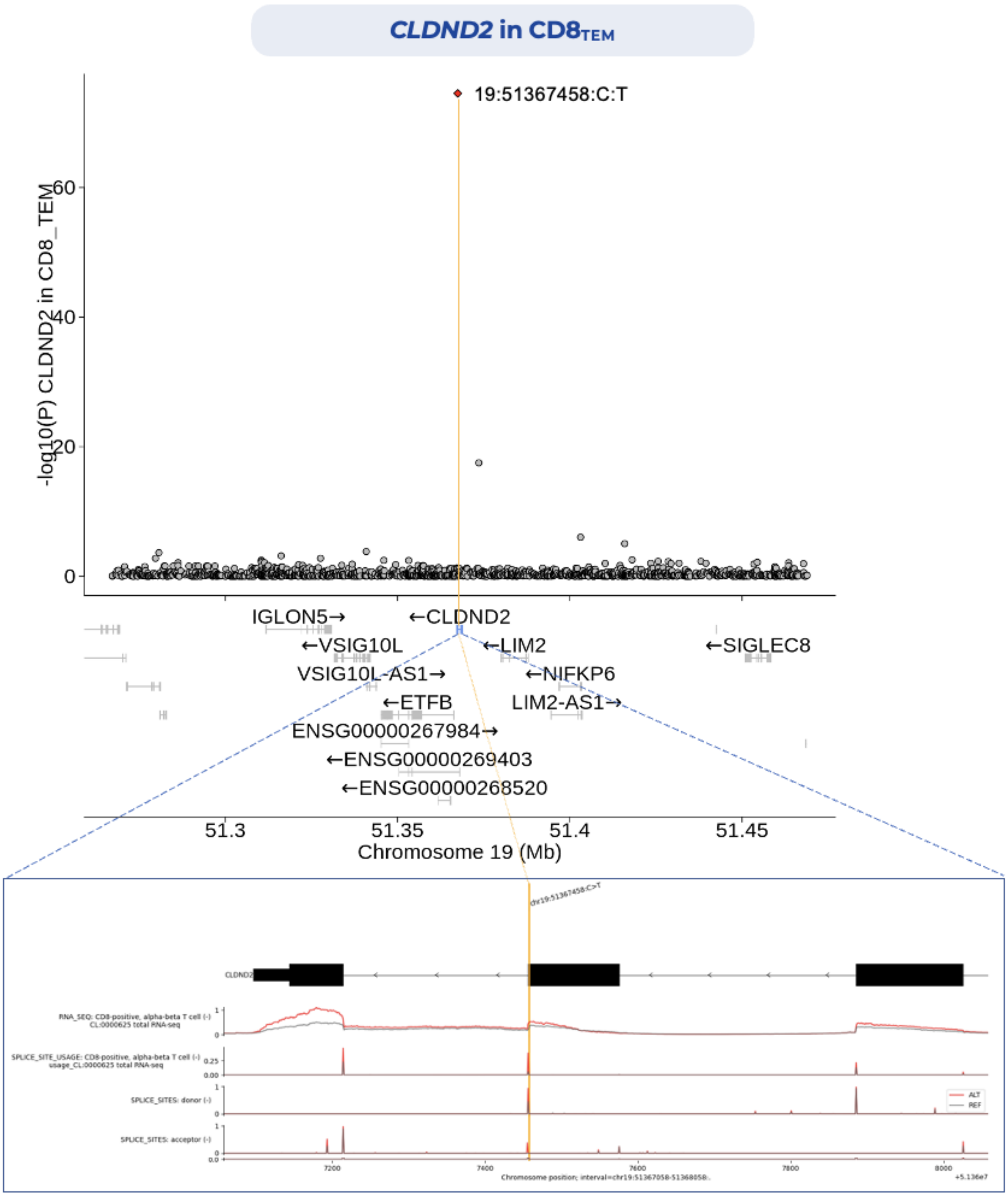


**Supplementary Figure 20. Rare variant eGene example: 19:51367458:C:T affecting splicing of *CLDND2*.** Locus zoom plot of rare genetic variants associated with the *CLDND2* gene on chromosome 19 in CD8TEM cells, with the leading splice-altering genetic variant (19:51367458:C:T) highlighted. Bottom plot shows AlphaGenome[9]-predicted RNA-seq coverage, splice sites usage, and splice sites tracks for exon 2, 3 and 4 of the *CLDND2* gene for the 19:51367458:C:T variant (ALT, red) and reference (REF, grey) alleles. Tracks are predicted for CD8-positive alpha-beta T cells (CL:0000625).

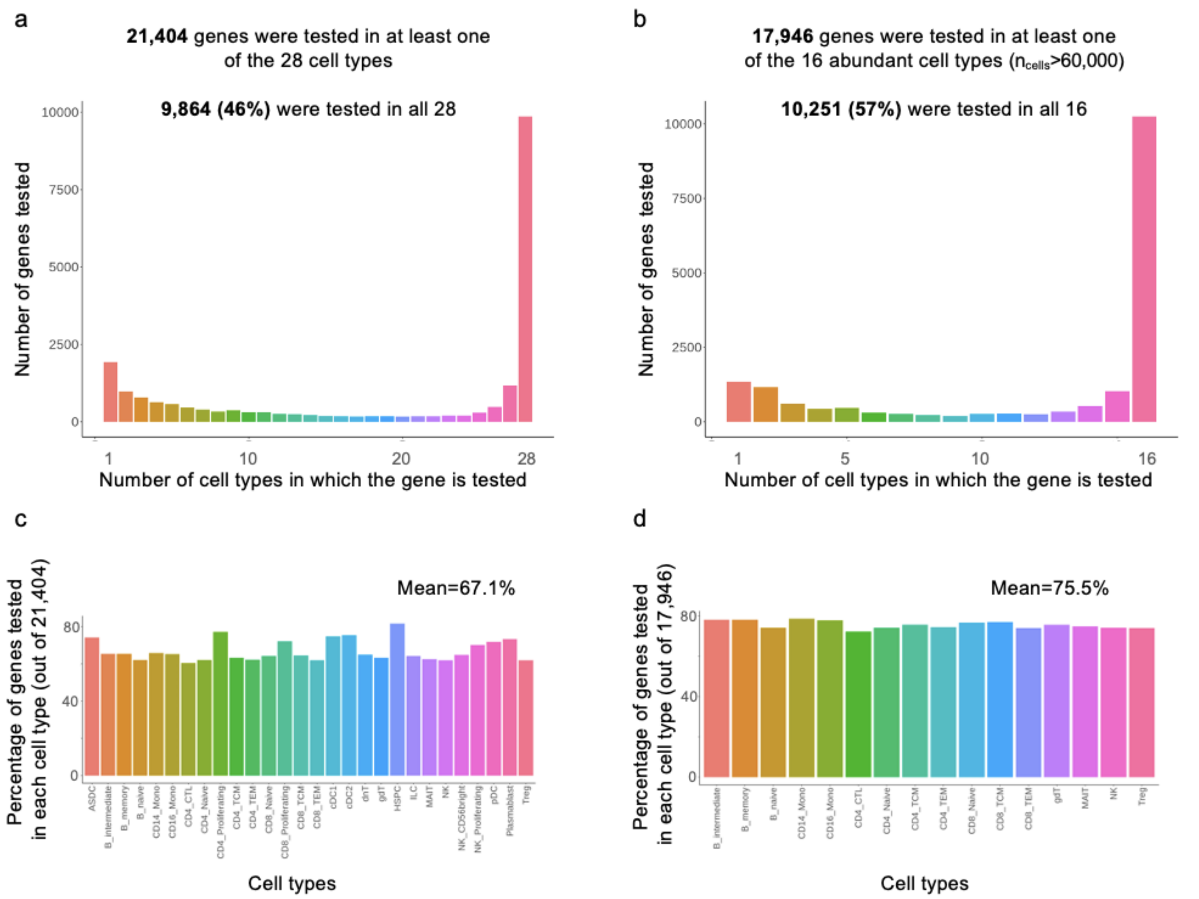


**Supplementary Figure 21. The overlap in number of genes tested across cell types. a)** Bar plot showing the number of cell types each gene is tested in: less than half of genes are tested in all 28 cell types. b) as **a,** but when only considering the 16 more abundant cell types ($n > 60{,}000$). **c)** Percentage of genes tested in each cell type (out of all of the genes tested in at least one of the cell types). **d)** as in c, but excluding rare cell types (as in b).

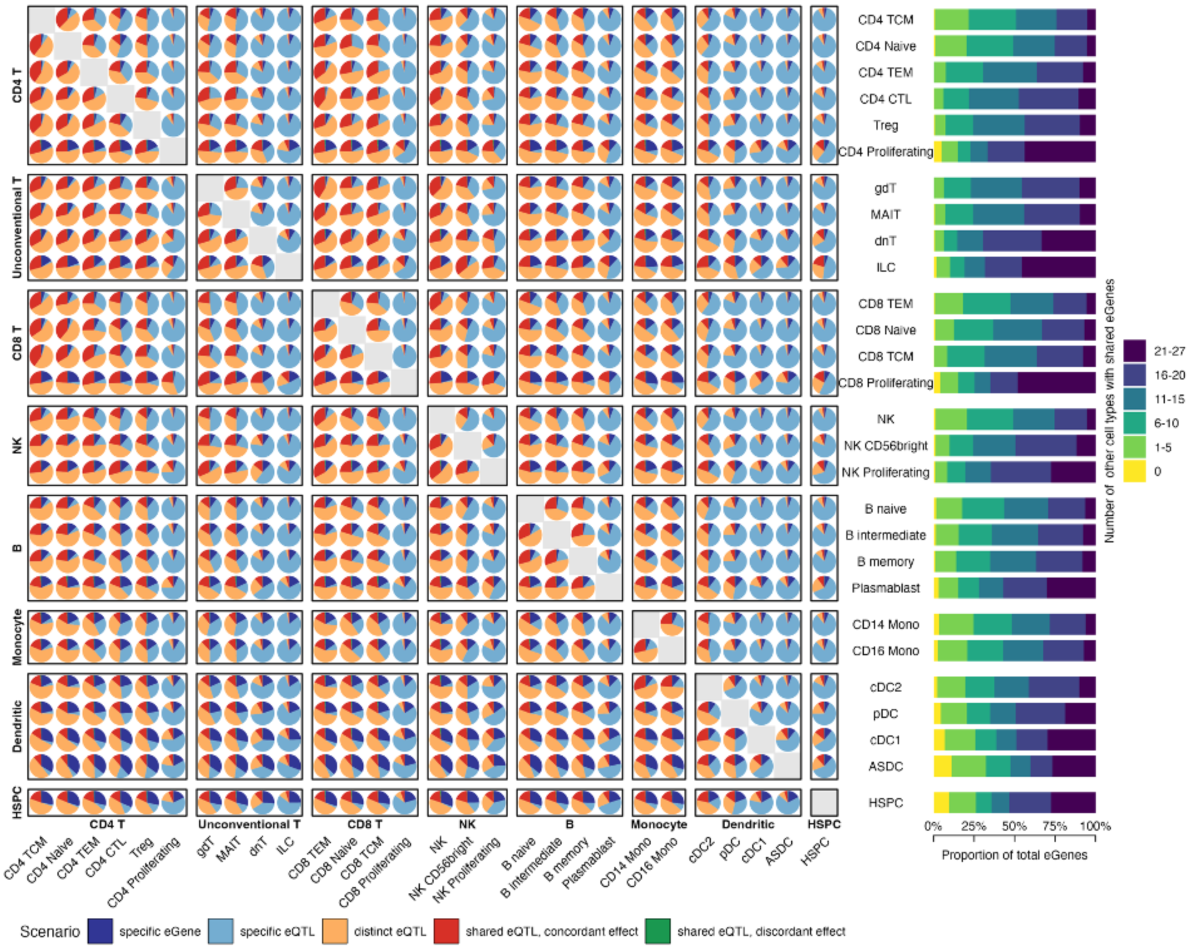


**Supplementary Figure 22. Cell type specificity patterns of common eQTL variants.** The pie charts on the left represent the proportion of cell type-specificity scenarios (see Methods) between pairs of cell types (primary cell type / cell type A in vertical axis, secondary cell type / cell type B in horizontal axis). Bar plots to the right show the proportion of eGenes binned by the number of cell types the effect is shared (scenarios 3 to 5) with. Cell types are grouped by major cell type categories.

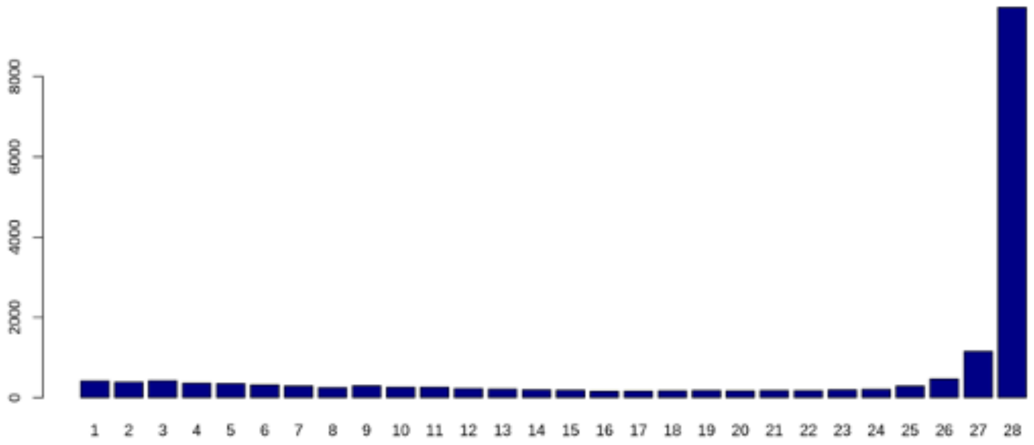


**Supplementary Figure 23. eGenes expression pattern across cell types.** The number of cell types in which each eGene is sufficiently expressed in (>1% all cells), from 1 to 28.

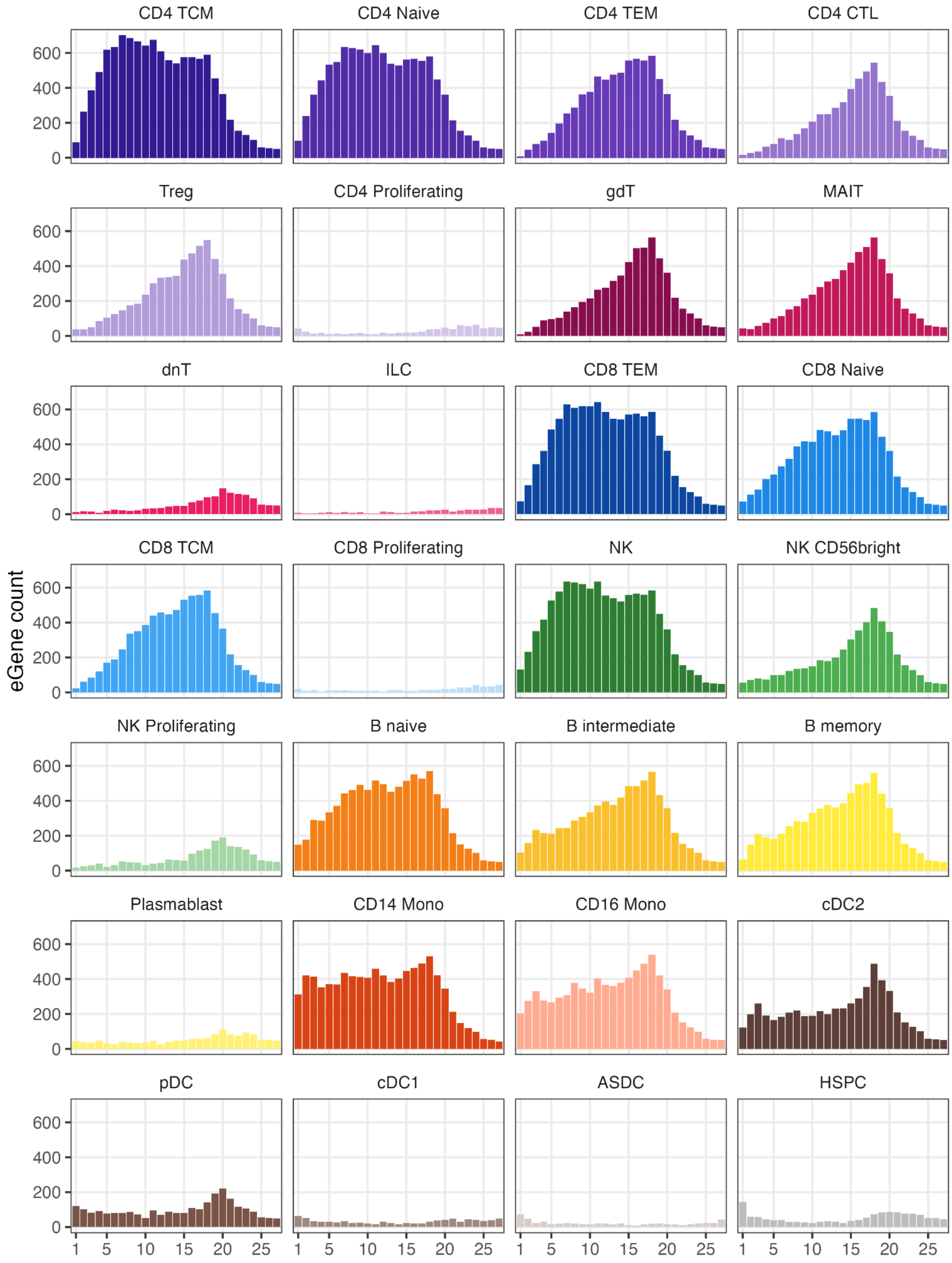


**Supplementary Figure 24. Distribution of shared eGene across cell types.** Each panel shows the number of eGenes (y-axis) identified in a given cell type (panel label, colour code) across number of other cell types wherein the same eGene was found (x-axis, ranging from 1 to 27).

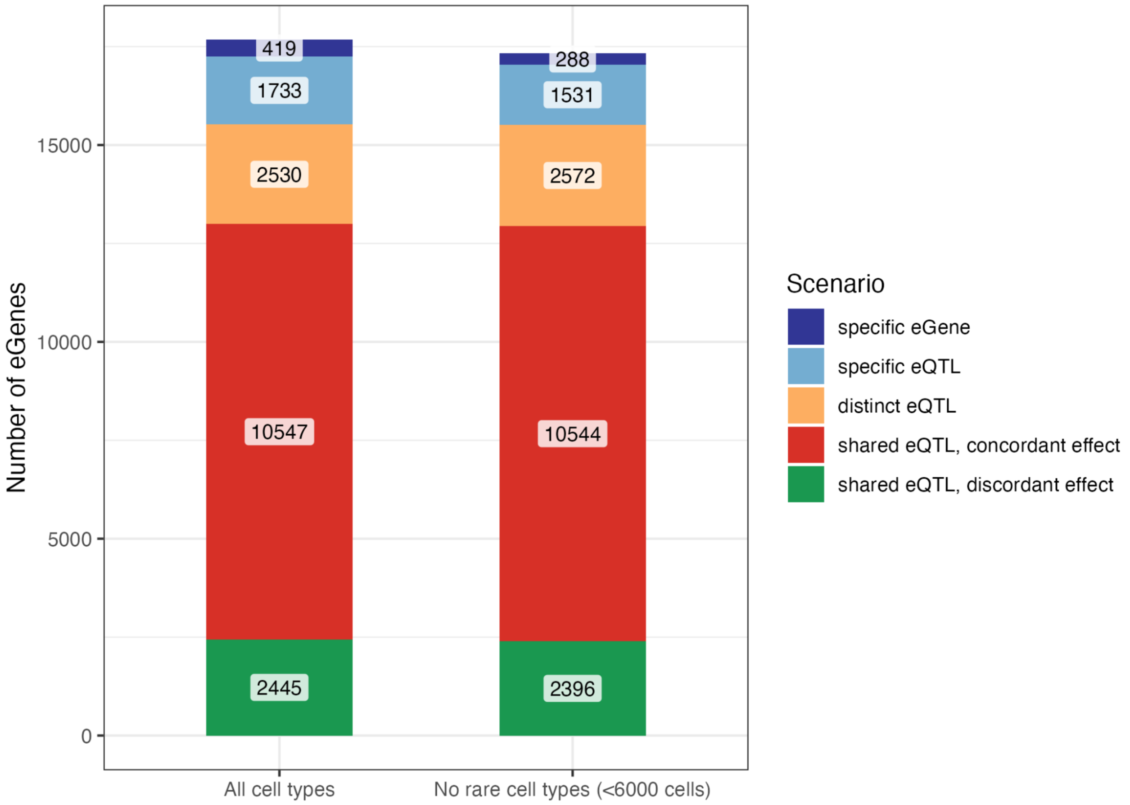


**Supplementary Figure 25. Number of eGenes by cell type specificity scenario when excluding rare cell types.** Number of eGenes stratified by cell-type specificity scenarios using gene-level aggregation strategy across all 28 cell types (left bar chart, same as panel 4b) and an alternative aggregation across 21 most abundant cell types (removing 7 cell types with <6000 cell count).

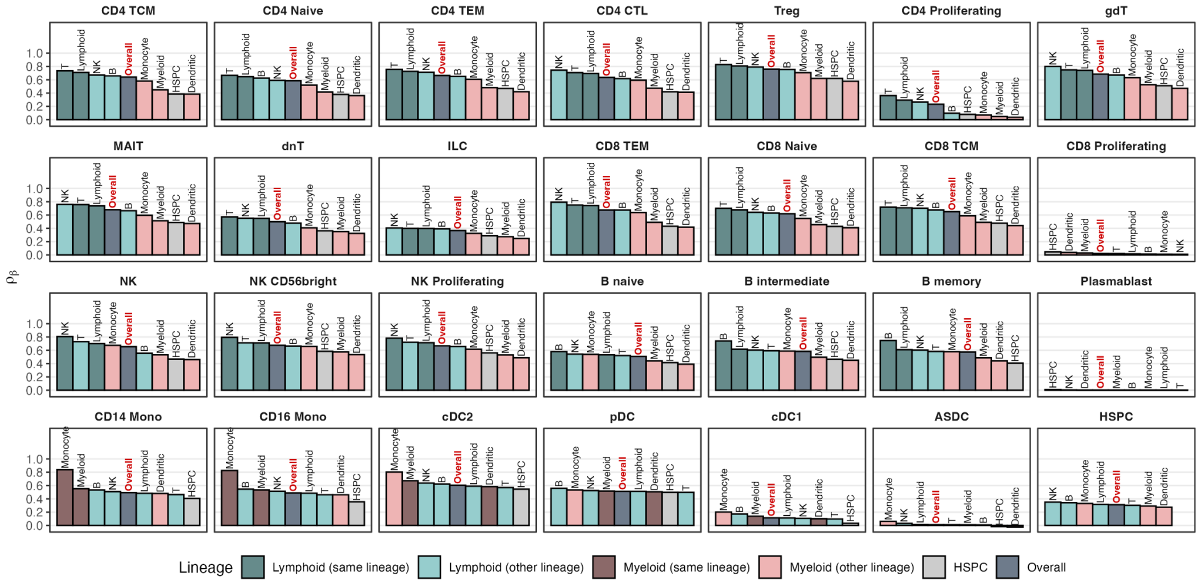


**Supplementary Figure 26. Correlation of eQTL effect across groups of cell types.** The correlation coefficient (shown on the y-axis) was calculated using Pearson's estimator per pair of cell types by taking the absolute effect size (regression coefficient / beta) for top eQTL variants in a given cell type (shown on the panel label) against every other cell type, averaged across groups of cell type (shown on the x-axis). Overall groups represent average across all other cell types. Lineage-based grouping of cell types is described on Supplementary Table 2.

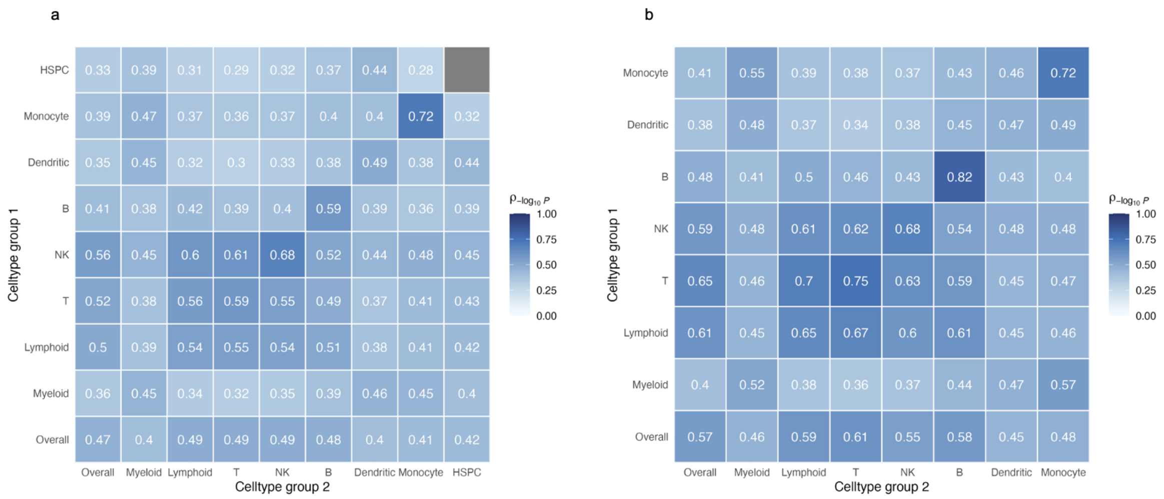


**Supplementary Figure 27. Pairwise Pearson's correlation of -log$_{10}$(*p*-value) eQTL association between major groups of cell types: a)** including all cell types **b)** excluding cell types with *n* cells <6,000.

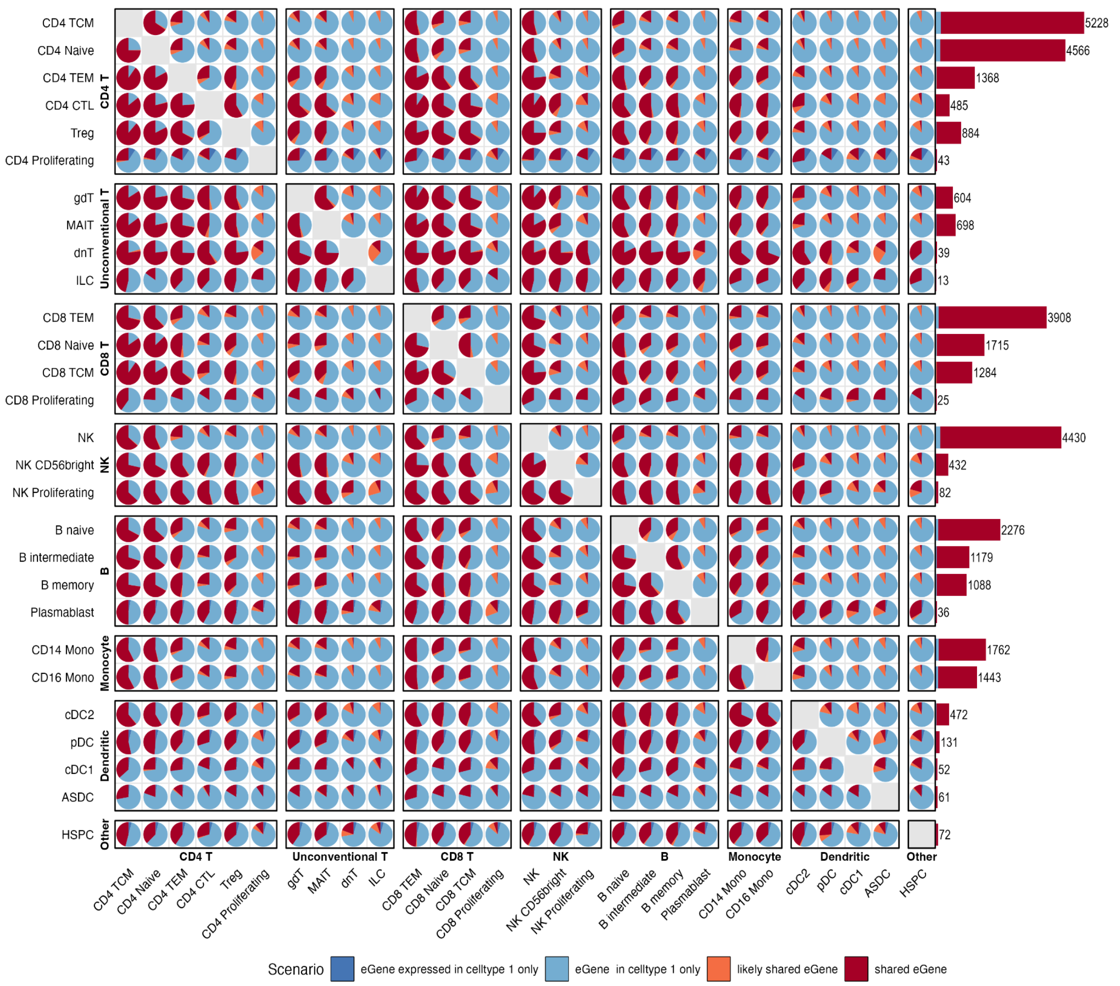


**Supplementary Figure 28. Cell type specificity patterns of rare variant eGenes.** The pie charts on the left represent the proportion of cell type-specificity scenarios (see **Methods**) between pairs of cell types (primary cell type / cell type A in vertical axis, secondary cell type / cell type B in horizontal axis). Bar plots on the right margin show counts of aggregated maximum scenarios per rare eGene in the primary cell type, annotated with the total count of eGenes for each cell type.

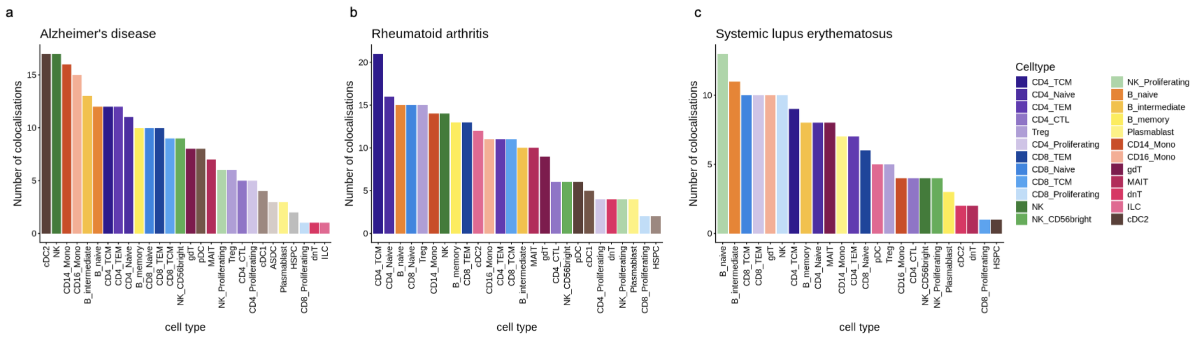


**Supplementary Figure 29. Cell type specificity of colocalizations for selected diseases.** Number of colocalization for each cell type for Alzheimer's disease, rheumatoid arthritis, and systemic lupus erythematosus.

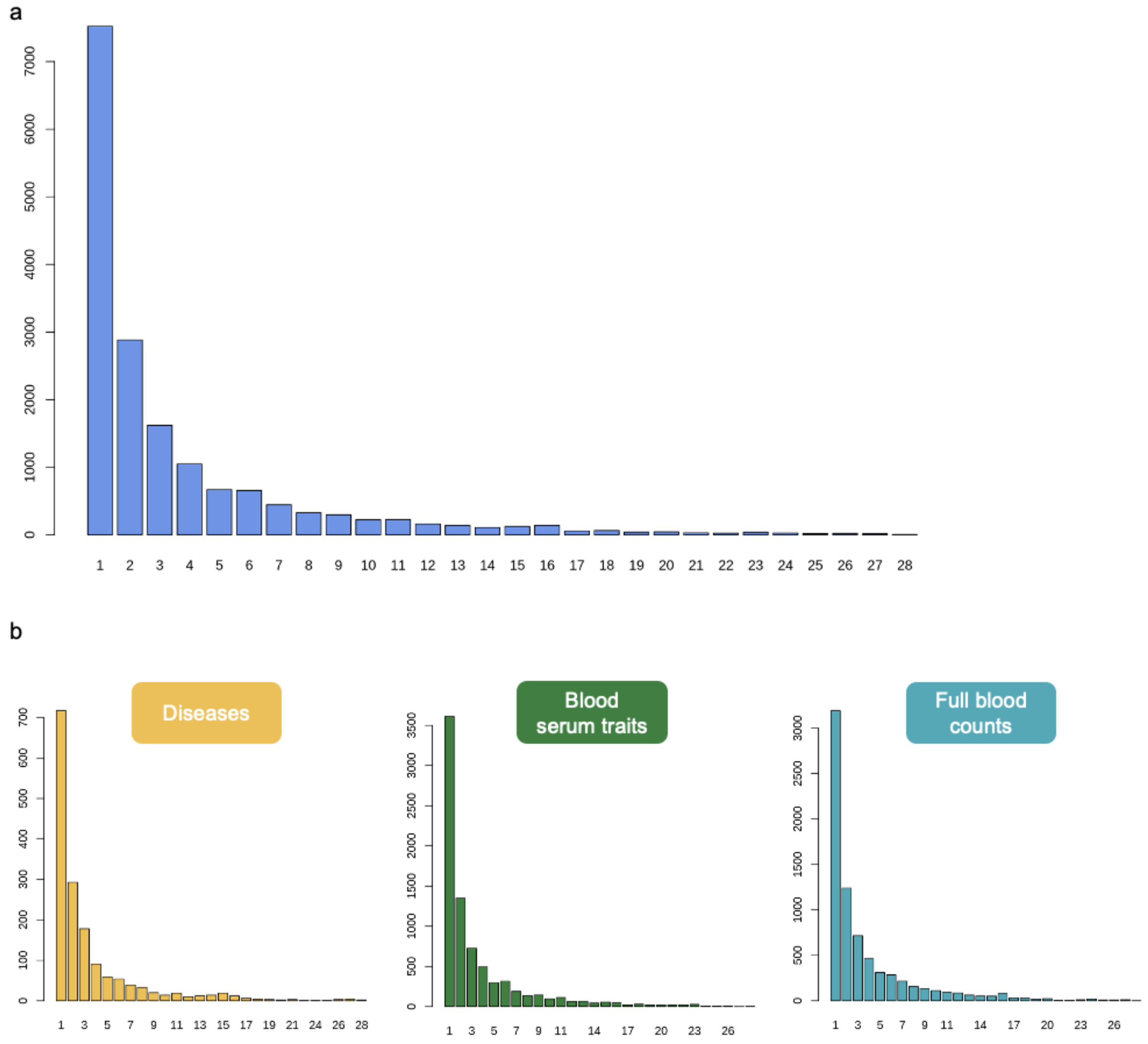


**Supplementary Figure 30. Cell type specificity of colocalization events. a)** Number of cell types each unique gene-trait colocalization pair was identified in at PP4>0.8, from 1 to 28. **b)** As in a, but split by trait category.

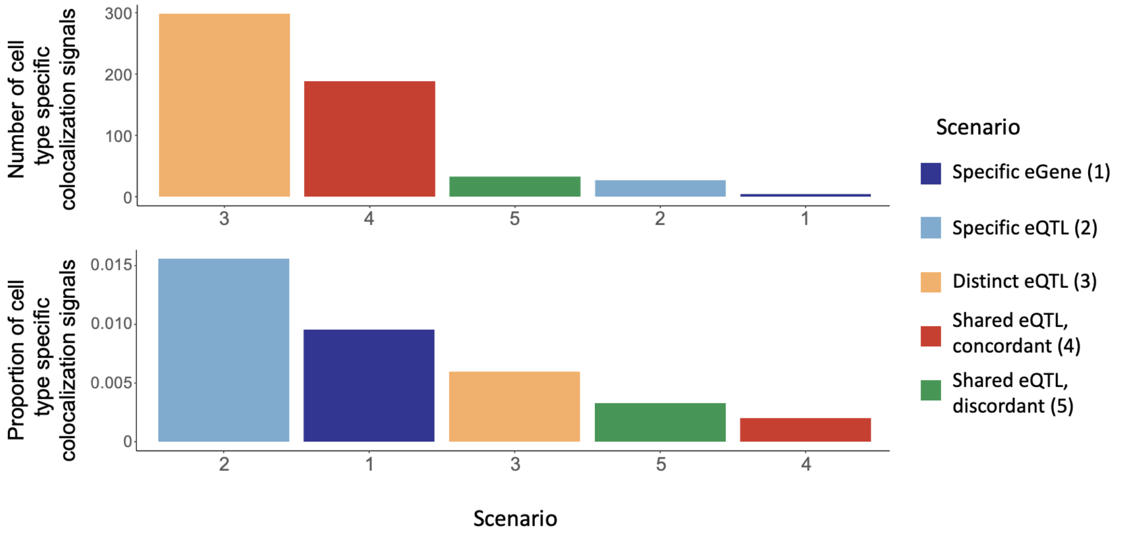


**Supplementary Figure 31. Cell type specificity of eGenes with evidence of colocalization. Top:** Number of colocalisation eGenes by cell type specificity scenario. Bottom: Proportion of colocalisation eGenes by cell type specificity scenario (divided by total number of eGenes in each scenario). Scenarios are as in Figure 4.

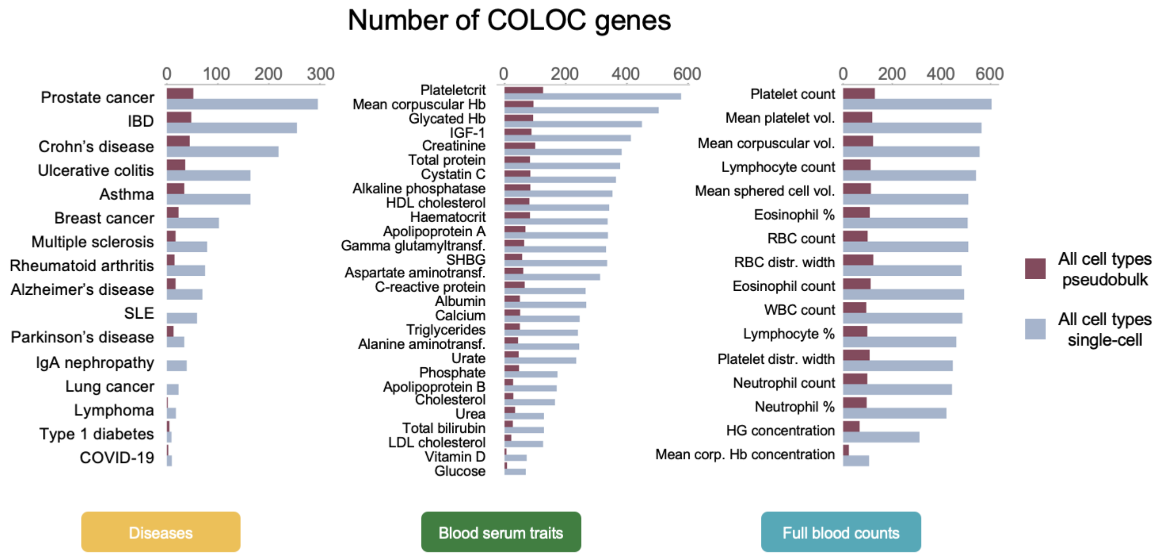


**Supplementary Figure 32. Number of eGenes with at least one colocalisation event found using an all-cell-type pseudobulk compared to single-cell results stratified by cell type.** Similar to Supplementary Figure 11 but quantifying the number of unique eGenes colocalising with each trait, divided by the type of trait (diseases, blood serum traits, full blood counts, as in Figure 5). For the "All cell type single-cell" results (blue bars), colocalisations identified in at least one of the 28 cell types were counted.

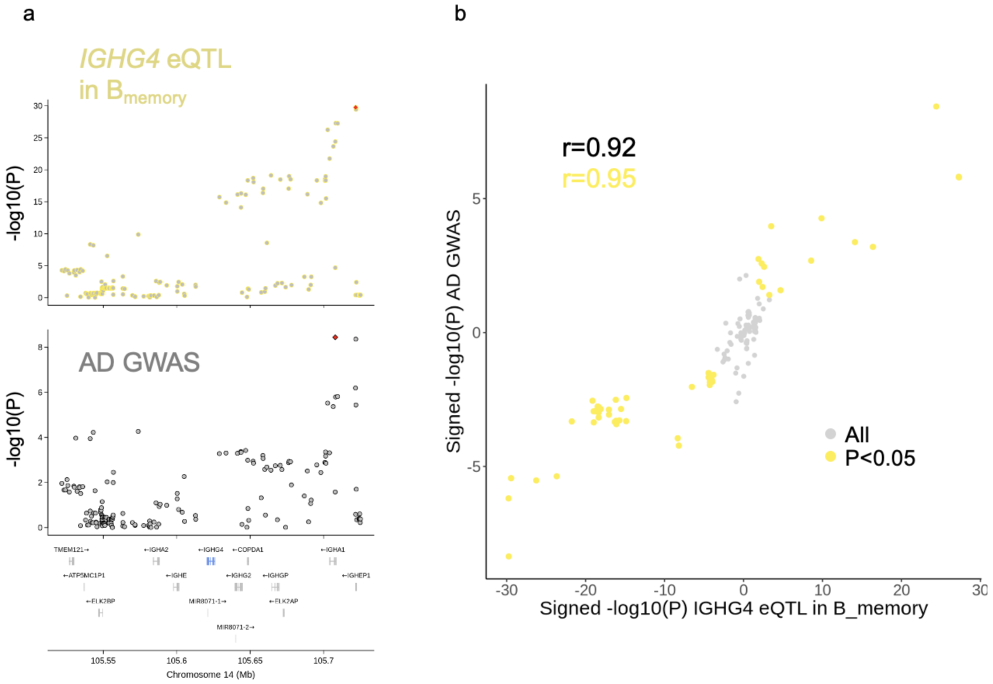


**Supplementary Figure 33. Colocalisation between Alzheimer's disease and *IGHG4* expression in memory B cells (PP4=0.99). a)** Locus zoom plot, -$\log_{10}$($p$-value) for all variants tested in the cis window around the *IGHG4* gene. Top: *IGHG4* eQTL in $B_{memory}$ cells; bottom: Alzheimer's disease (AD) GWAS. **b)** Scatter plot of -$\log_{10}$($p$-value) signed by effect size (multiplied by -1 if effect size is negative, 1 if positive). All in grey, $p$-value <0.05 in yellow.

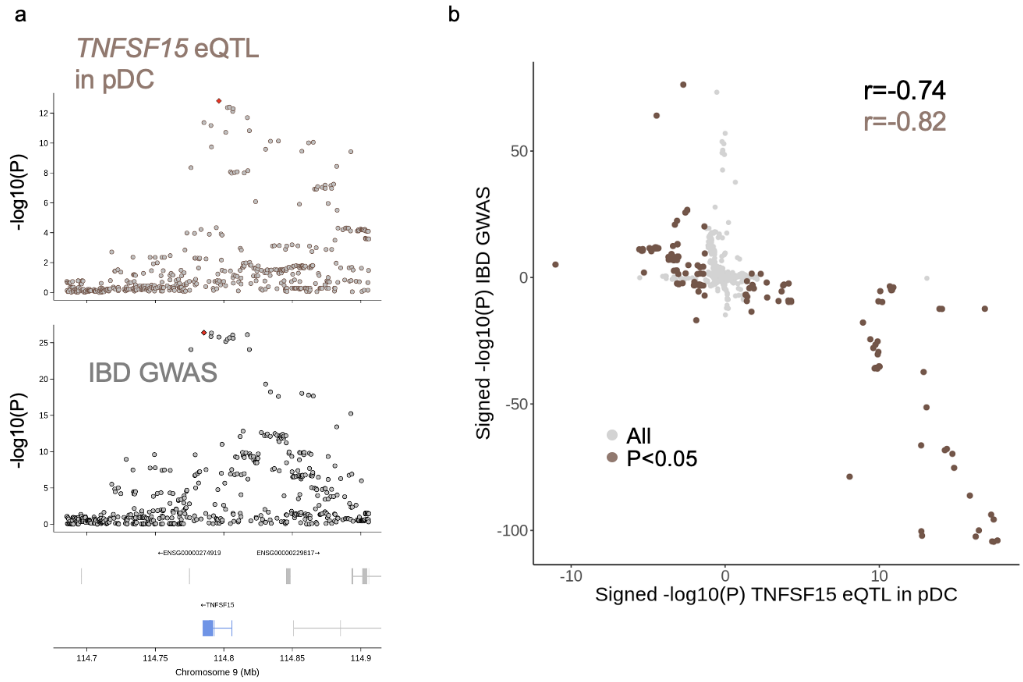


**Supplementary Figure 34. Colocalisation between inflammatory bowel disease and *TNFSF15* expression in memory pDC (PP4=0.99). a)** Locus zoom plot, -$\log_{10}$($p$-value) for all variants tested in the cis window around the *TNFSF15* gene. Top: *TNFSF15* eQTL in pDC cells; bottom: inflammatory bowel disease (IBD) GWAS. **b)** Scatter plot of -$\log_{10}$($p$-value) signed by effect size (multiplied by -1 if effect size is negative, 1 if positive). All in grey, $p$-value <0.05 in brown.

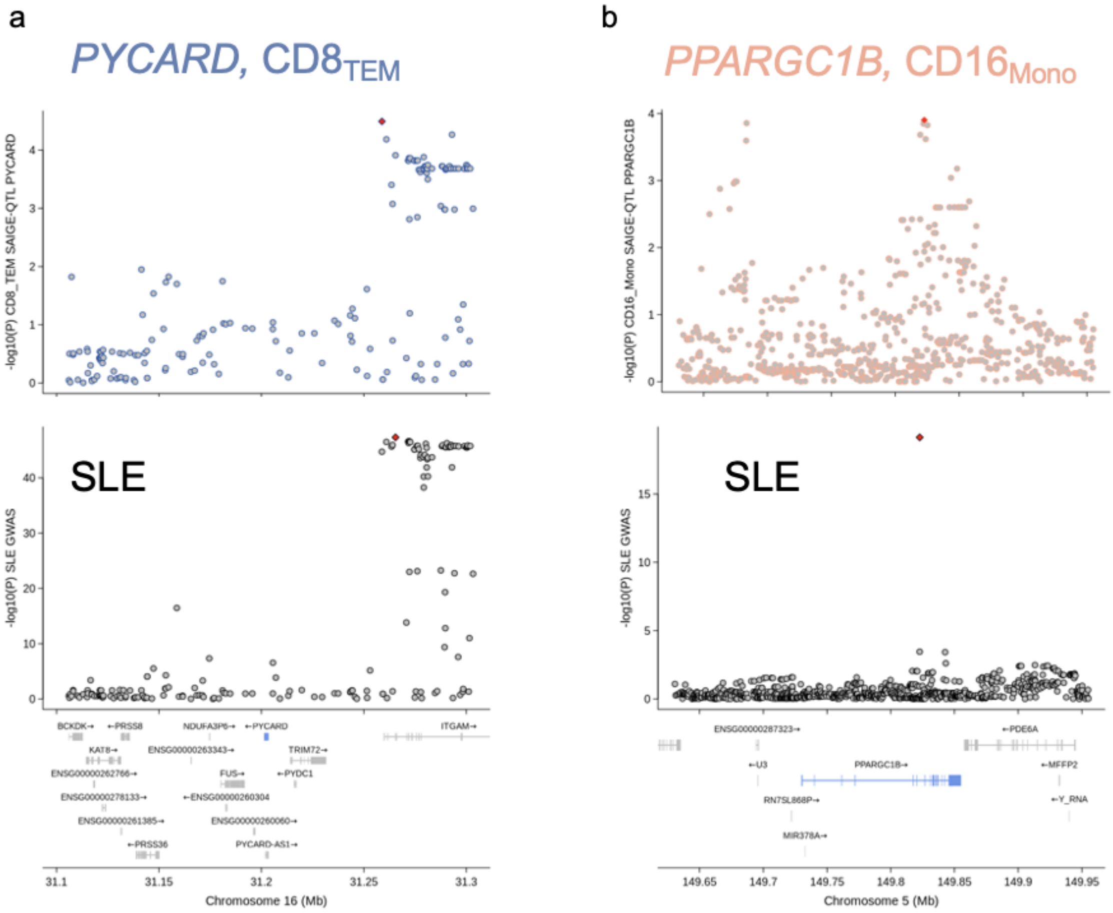


**Supplementary Figure 35. More colocalisation event examples. a)** *PYCARD* gene in $CD8_{TEM}$ and both SLE (PP4=0.91); **b)** colocalisation between a non-classical monocyte ($CD16_{Mono}$)-specific eQTL for *PPARGC1B* and SLE (PP4=0.94).

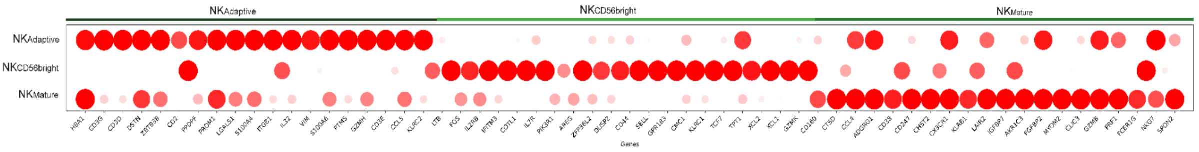


**Supplementary Figure 36. Relative marker expression of three representative sub-NK cell types.** Following the label transfer from the reference dataset[10], we select the representative markers the same as the original publication, and calculate the average expression of the markers within each of the 5 cell sub cell types. Then the relative expression is defined by the min-max normalisation of the average expression of each subtype.

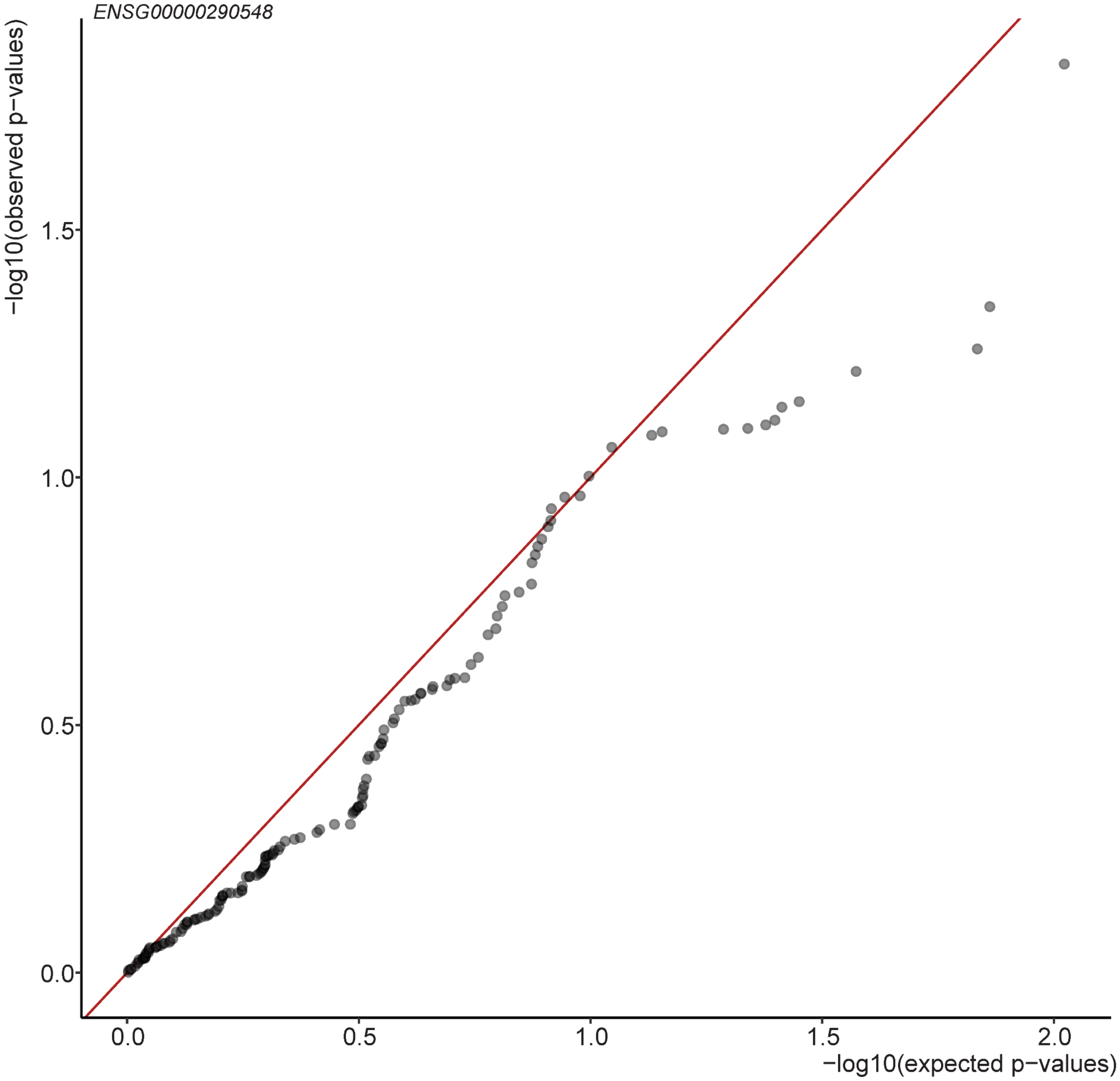


**Supplementary Figure 37.** Cell state eQTL tests are calibrated. We randomly select the *RPS26* gene together with 150 random SNPs, and mapped cell state eQTLs using a linear mixed model with an interaction term and the cell state scores of the cell killing function.

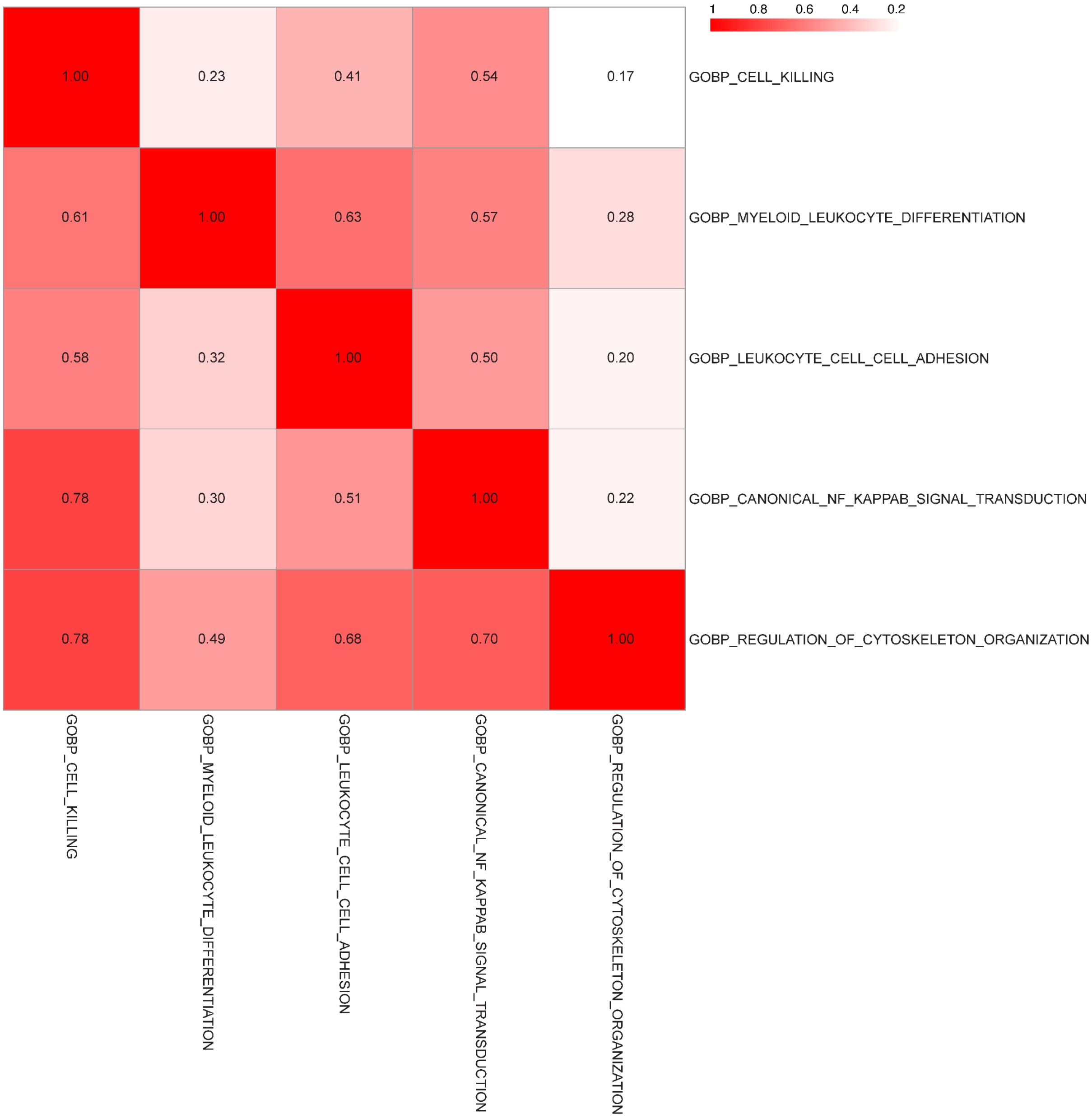


**Supplementary Figure 38.** Overlap of cell state eQTL mapping in NK cells across cell functions. The proportions of the shared eQTLs are calculated by the shared number of eQTLs (between the cell states in each row and each column) divided by the number of eQTLs in the column.

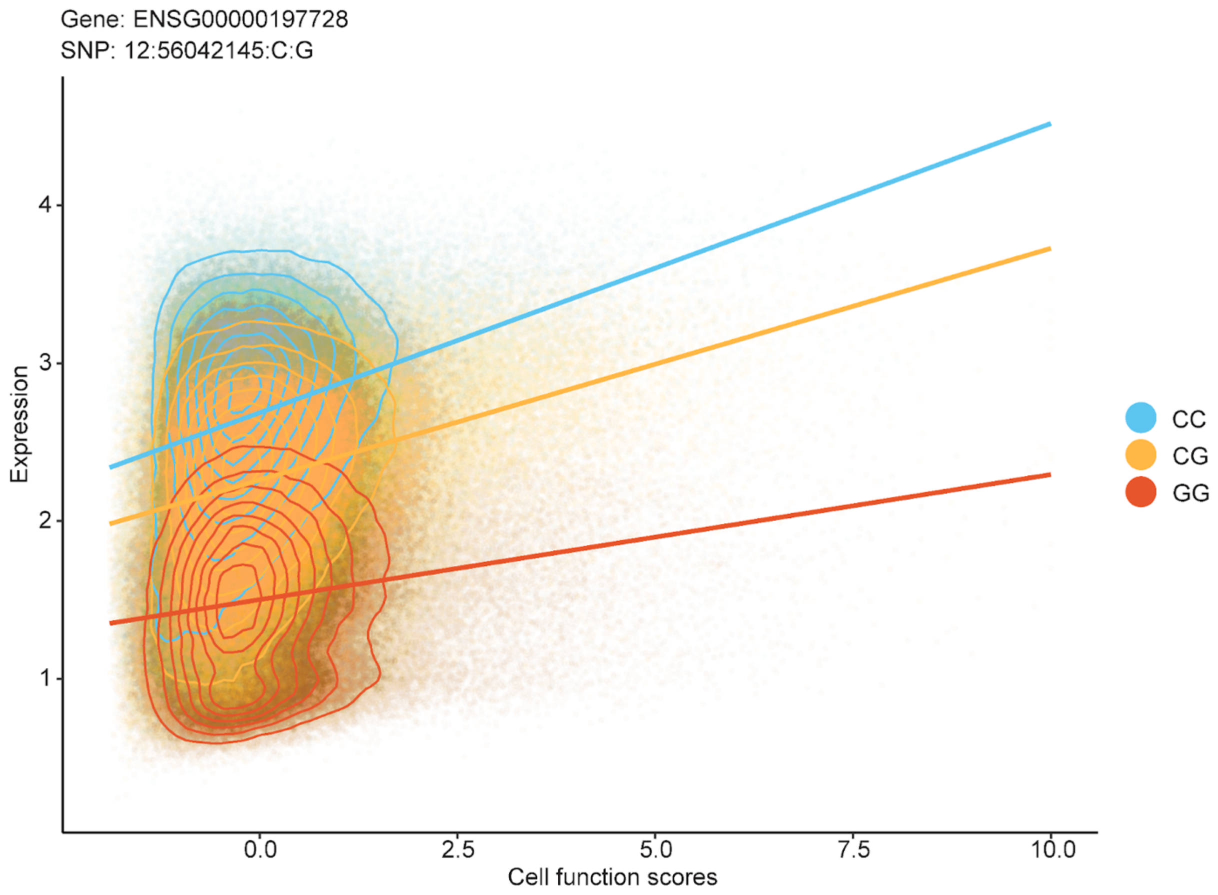


**Supplementary Figure 39. NK dynamic eQTL example: *RPS26* eQTL is modulated by the cell killing function.** Similar to Fig. 6d and e but for *RPS26* (ENSG00000197728), 12:56042145:C:G (rs1131017) and cell killing.

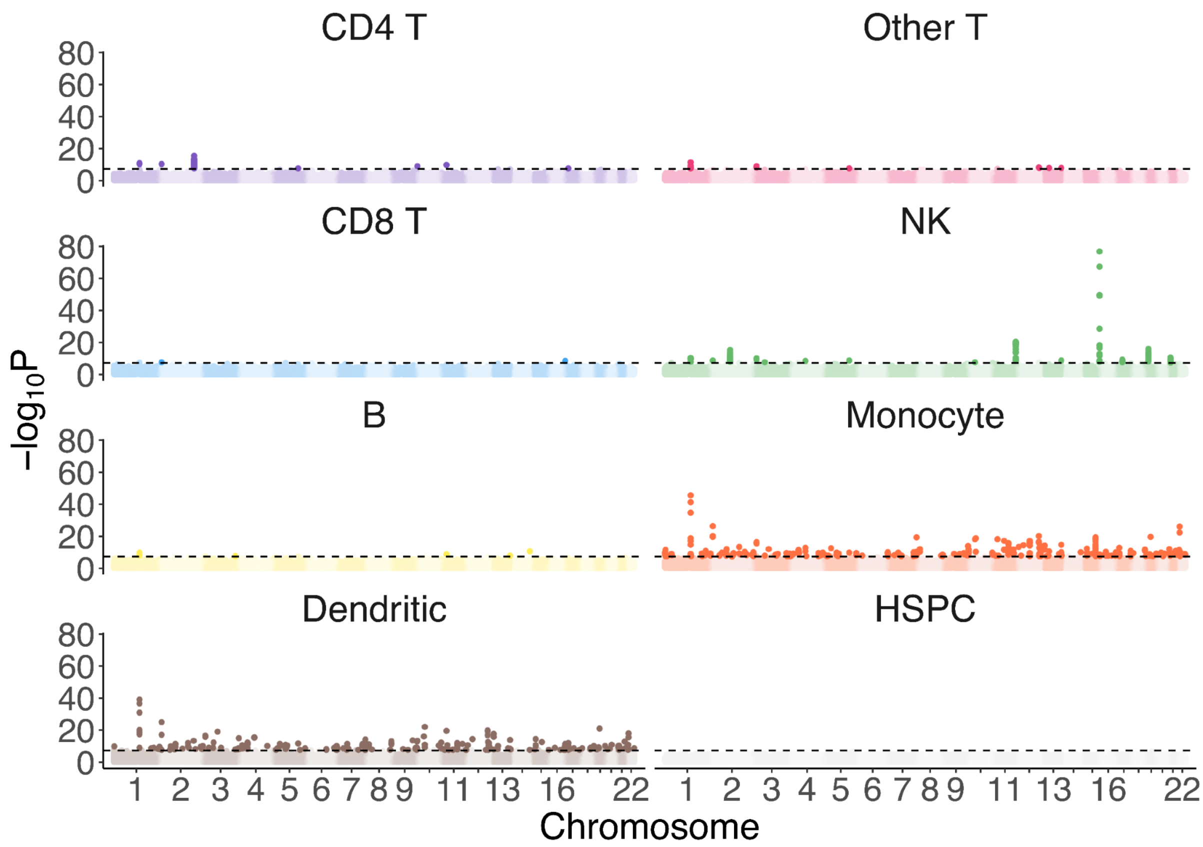


**Supplementary Figure 40. Cell state abundance Manhattan plots.** Manhattan plot illustrating csaQTLs identified with GeNA[11] for all major cell types. Dashed lines indicate the genome-wide significance threshold (*p*-value < $5\times10^{-8}$), and loci passing this threshold are coloured according to the cell type they were significant in.

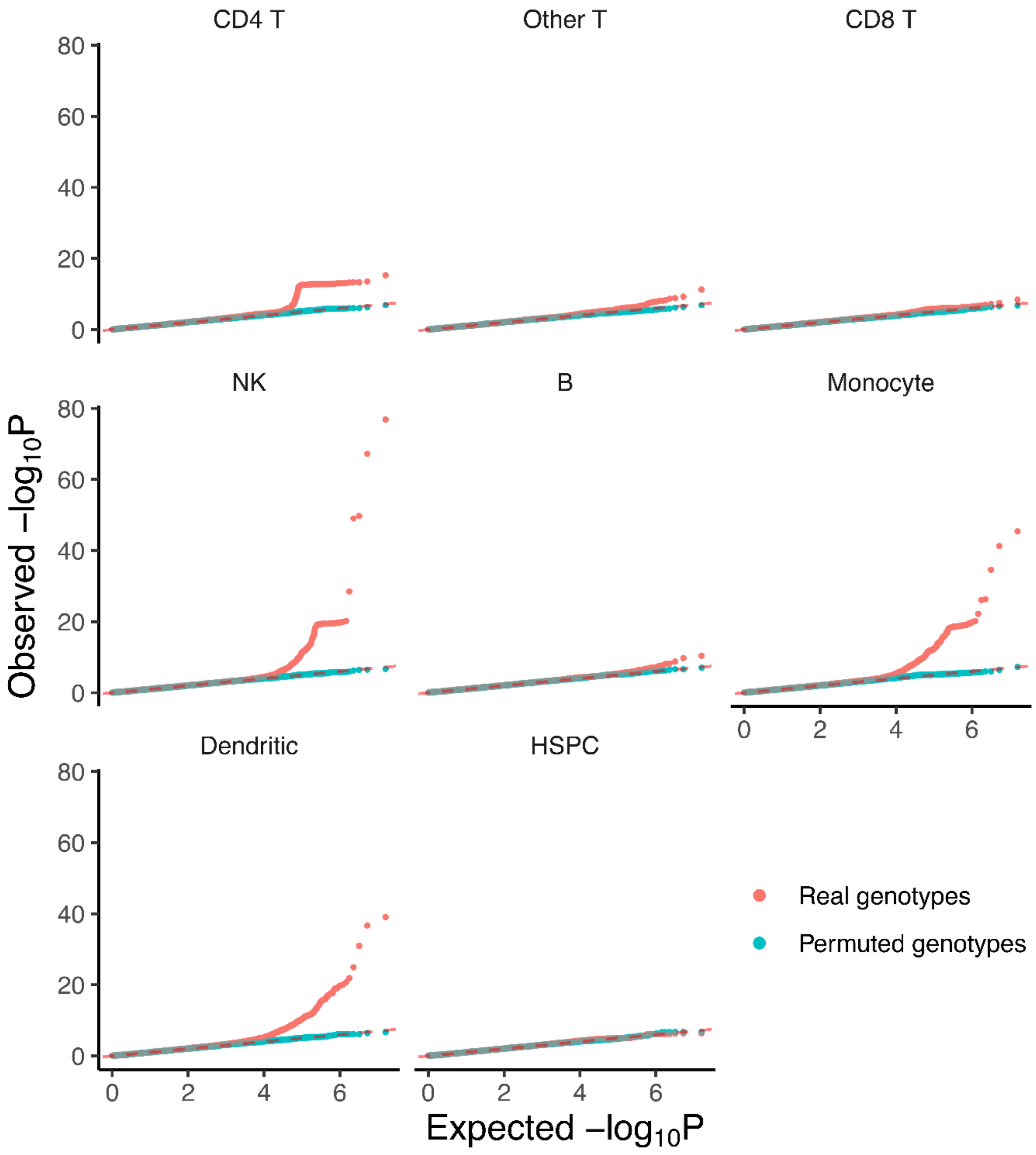


**Supplementary Figure 41. Cell state abundance QQ plots for variants with MAF>=5%.** GeNA csaQTL *p*-values. Permuted *p*-values were generated by running GeNA with genotype data with randomly shuffled sample labels. Permuted genotypes closely follow a uniform (null) distribution, indicated by the dashed red line.

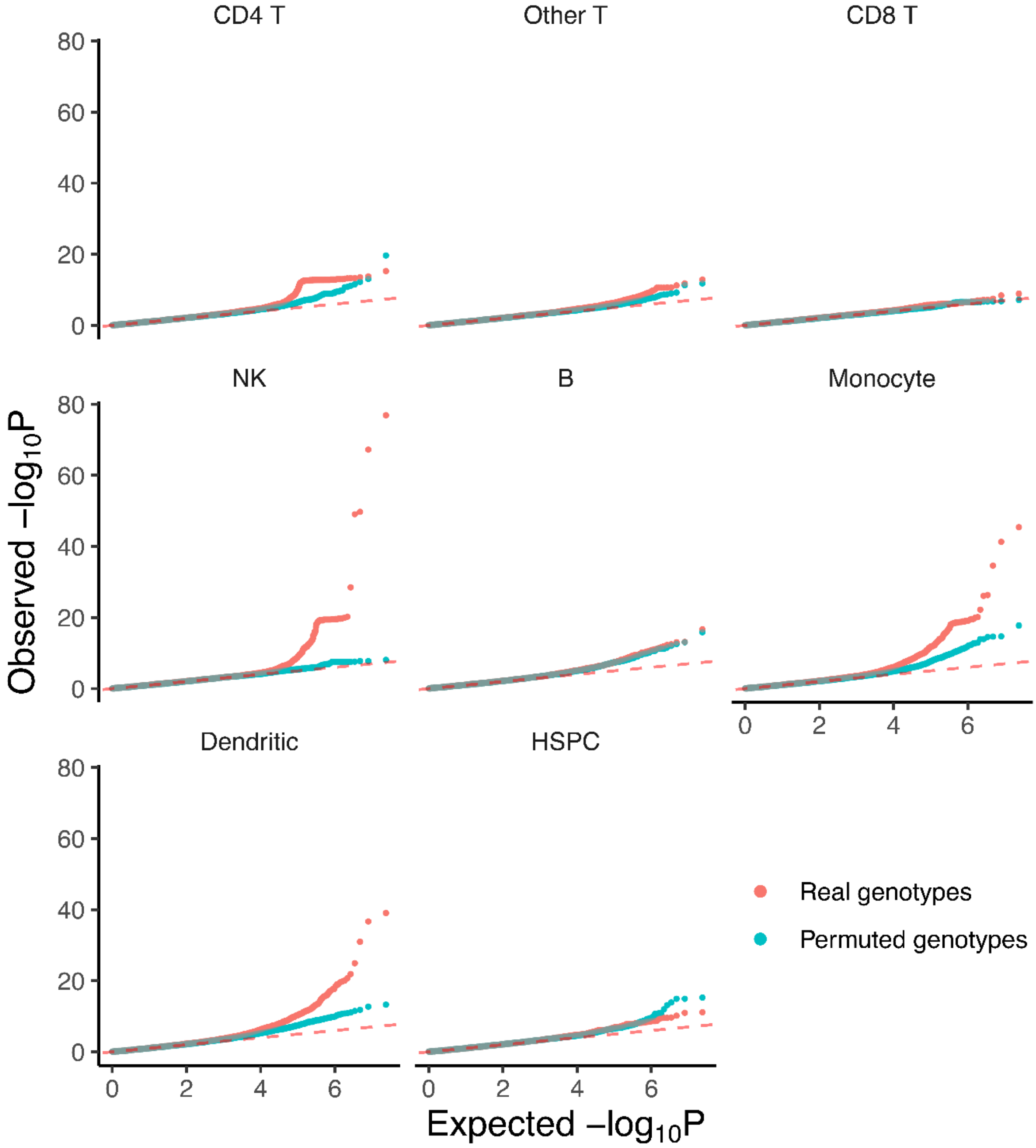


**Supplementary Figure 42. Cell state abundance QQ plots for variants with MAF>=1%.** QQ plots for GeNA csaQTL *p*-values. Permuted *p*-values were generated by running GeNA with genotype data with randomly shuffled sample labels. Permuted genotypes deviate from the uniform (null) distribution, indicated by the dashed red line. This indicates poor calibration for variants with 1-5% MAF, which were ultimately excluded from the analysis.

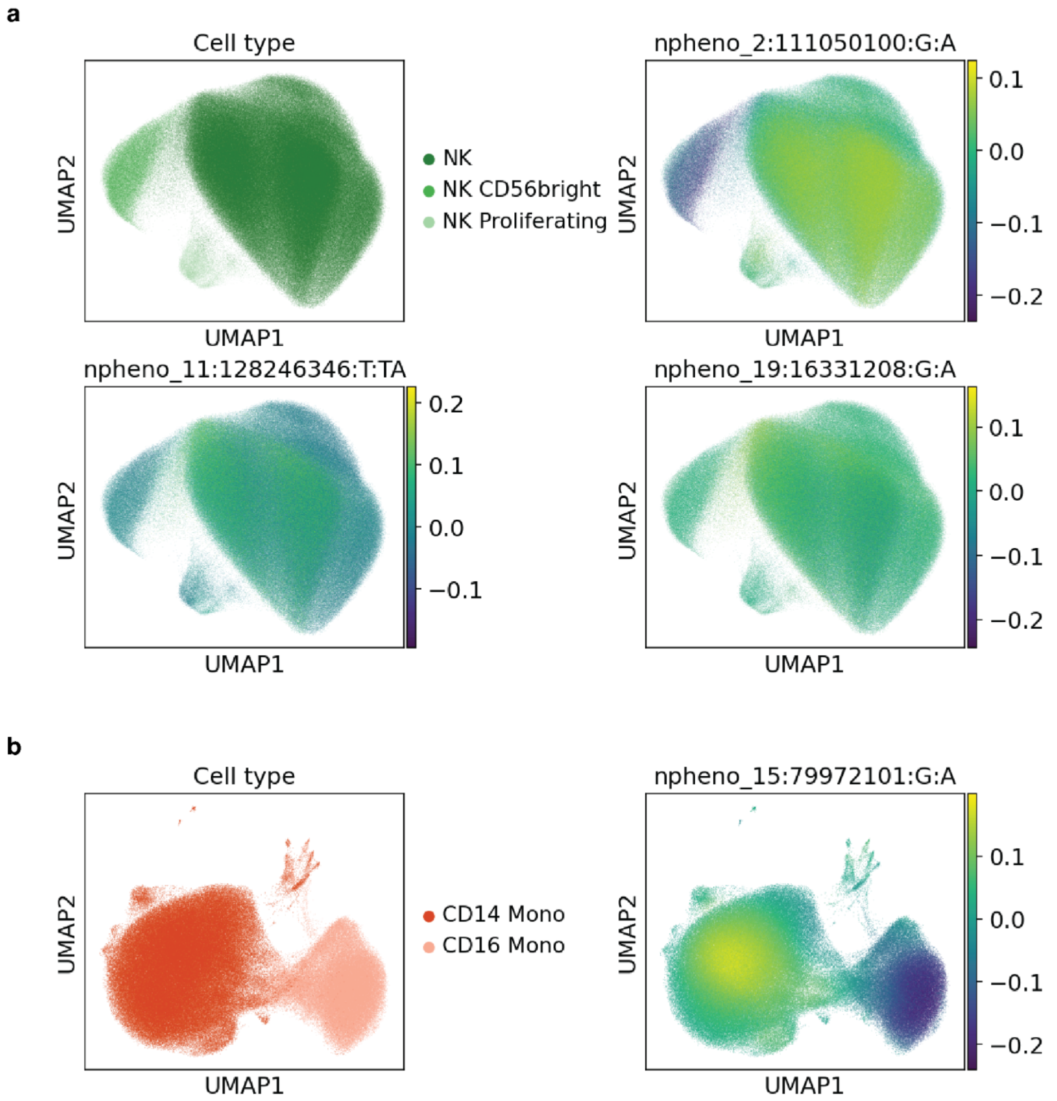


**Supplementary Figure 43. Validation of csaQTLs from GeNA original publication.** UMAP projections for the replicated csaQTL previously identified by Rumker *et al.*[11] with GeNA in the OneK1K cohort. The loci visualised are the lead variants from the TenK10K phase 1 (this paper) analysis within 100Kb of the lead SNPs identified in the OneK1K analysis (outlined in Supplementary Table 5). UMAPs are coloured by the sample-level phenotype for each lead SNP.

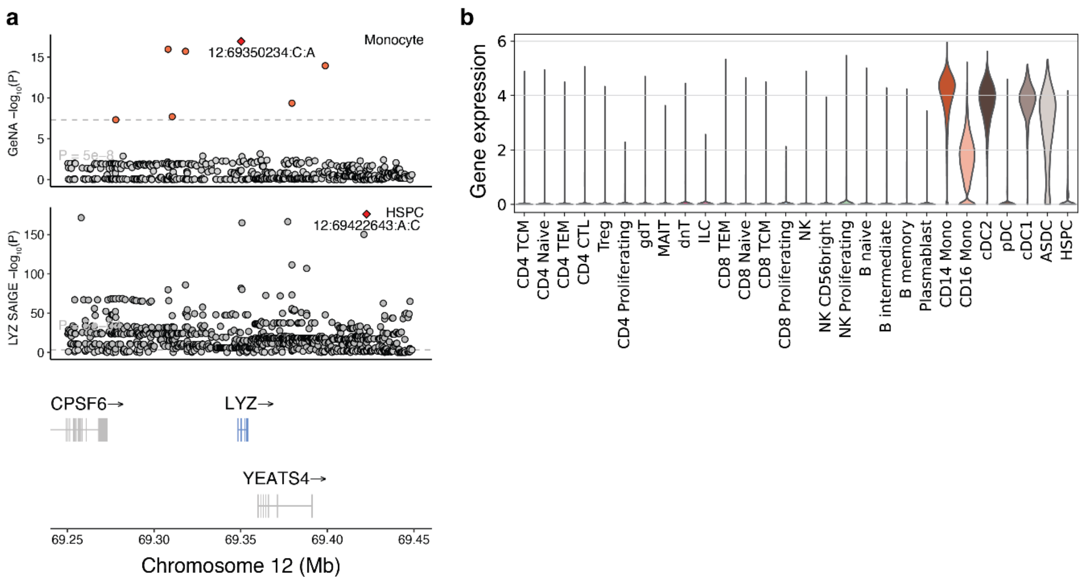


**Supplementary Figure 44. Supplementary info about *LYZ* csaQTL. a)** Locuszoom plot for csaQTL 12:69350234:C:A (Fig. 7b-f), which colocalised with *LYZ* in HSPCs (PP4 = 0.92). The upper track shows GeNA $-\log_{10}$($p$-values), lower tracks show SAIGE-QTL $p$-values in HSPC. **b)** Violin plot showing *LYZ* gene expression in each cell type.

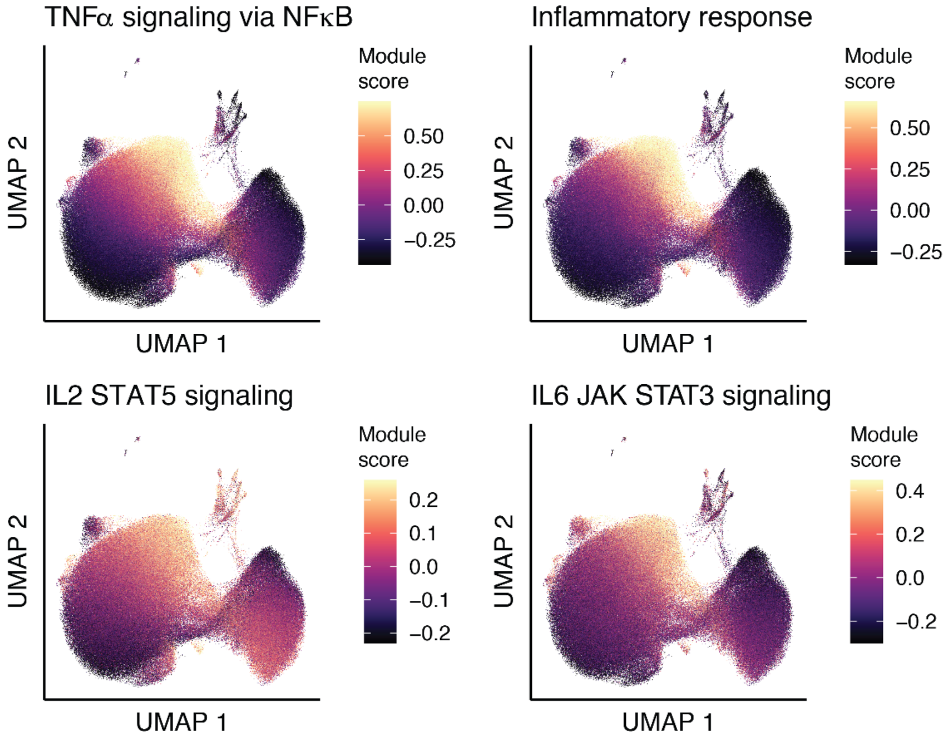


**Supplementary Figure 45. Characterisation of csaQTL-associated cell states with gene set enrichment analysis.** UMAP of Monocytes coloured by gene set module scores for Hallmark gene sets that were enriched within expanded cell states (Methods) associated with 12:69350234:C:A (Fig. 7b-f) and 12:9953308:T:TG (Supplementary Fig. 46). TNFα signalling via NFκB was significantly enriched within the phenotypes associated with both 12:69350234:C:A (BH-adjusted $p$-value = $7.87\times10^{-8}$) as well as 12:9953308:T:TG (BH-adjusted $p$-value = $6.51\times10^{-44}$). Inflammatory response pathways were also enriched within both 12:69350234:C:A (BH-adjusted $p$-value=$9.03\times10^{-8}$) and 12:9953308:T:TG (BH-adjusted $p$-value = $1.70\times10^{-22}$) associated phenotypes. The 12:9953308:T:TG csaQTL was also associated with increased IL2 STAT5 signalling (BH-adjusted $p$-value = $1.74\times10^{-4}$) and IL6 JAK STAT3 signaling (BH-adjusted $p$-value = $2.46\times10^{-4}$).

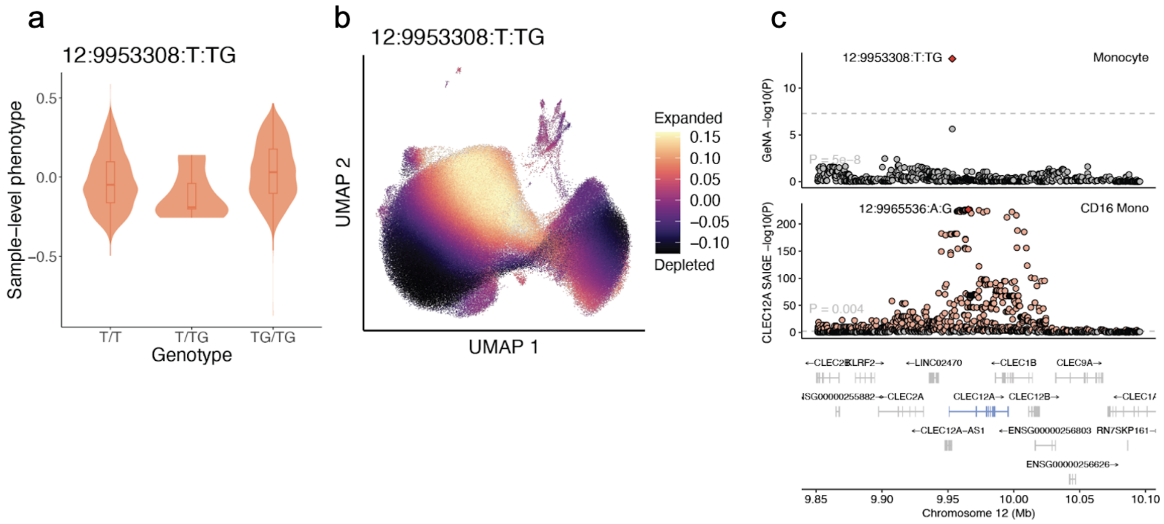


**Supplementary Figure 46. Monocyte csaQTL example: *CLEC12A*. a)** GeNA sample-level phenotype values for csaQTL variant 12:9953308:T:TG by genotype. **b)** UMAP projection of monocytes, coloured by GeNA neighborhood-level phenotype values for csaQTL variant 12:9953308:T:TG. **c)** Locus zoom plot for 12:9953308:T:TG, which was also intersecting a significant eQTL for *CLEC12A*. The upper track shows GeNA -log10(*p*-values), lower tracks show SAIGE-QTL -log10(*p*-values) in CD16Mono.

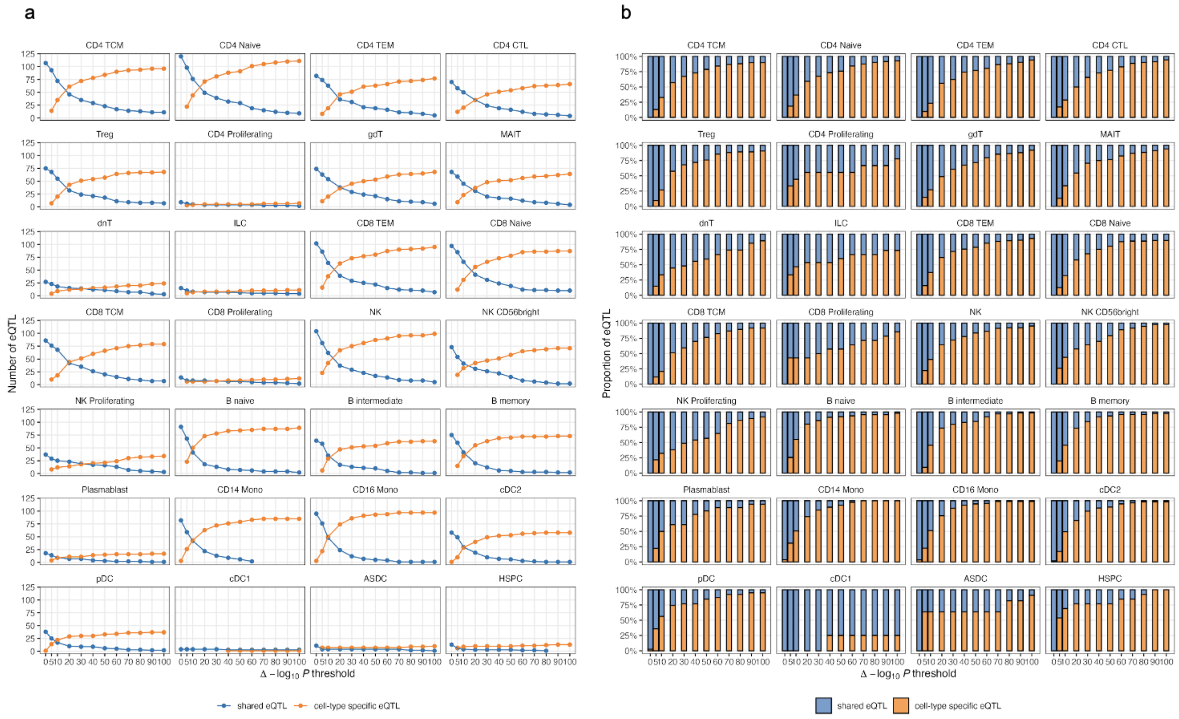


**Supplementary Figure 47. Shared and cell type specific eQTLs using different meta-analysis *p*-value to the primary *p*-value delta thresholds.** Number (a) and proportion (b) of eQTL classified as shared or cell-type specific across a range of possible threshold for -log10(*p*-value) difference between the original eQTL association analysis and meta-analysis approach to identify sharedness (delta -log10(*p*-value) threshold, x-axis). The detail of the meta-analysis approach is described in the Methods section of the main text.

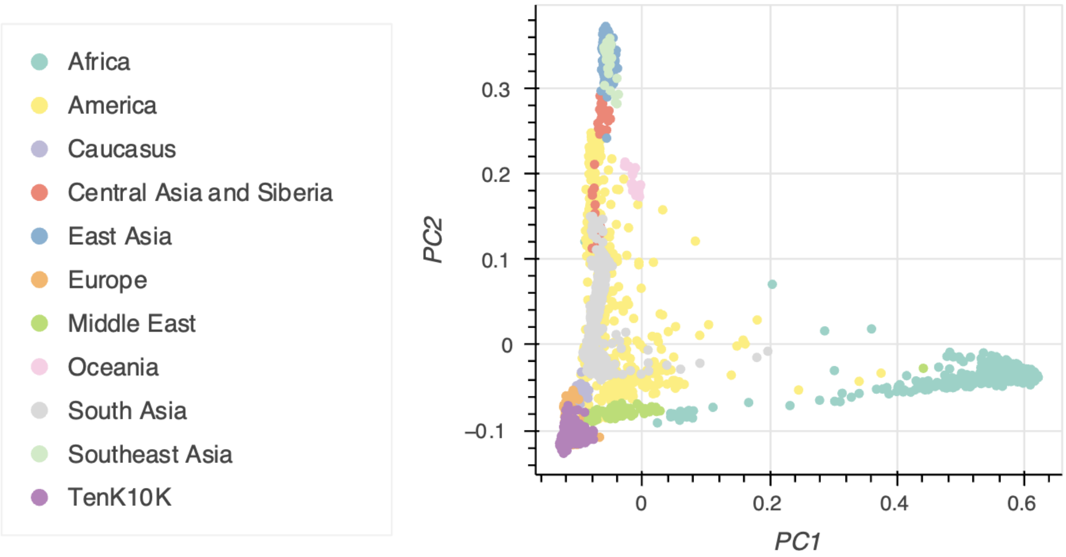


**Supplementary Figure 48. Principal component analysis (PCA) of TenK10K Phase 1 cohort samples overlaid with global reference populations.** Scatter plot of the first two principal components derived from SNVs, showing genetic variation among study participants (purple) in the context of reference samples from the Human Genome Diversity Project (HGDP) and 1000 Genomes Project (1KGP). Reference samples are labelled by major continental ancestry groups. Study cohort samples cluster in genetic principal component space primarily with participants self-described as of European ancestry. Identical to Supplementary Fig. 1 from accompanying manuscript Tanudisastro *et al*, 2025.

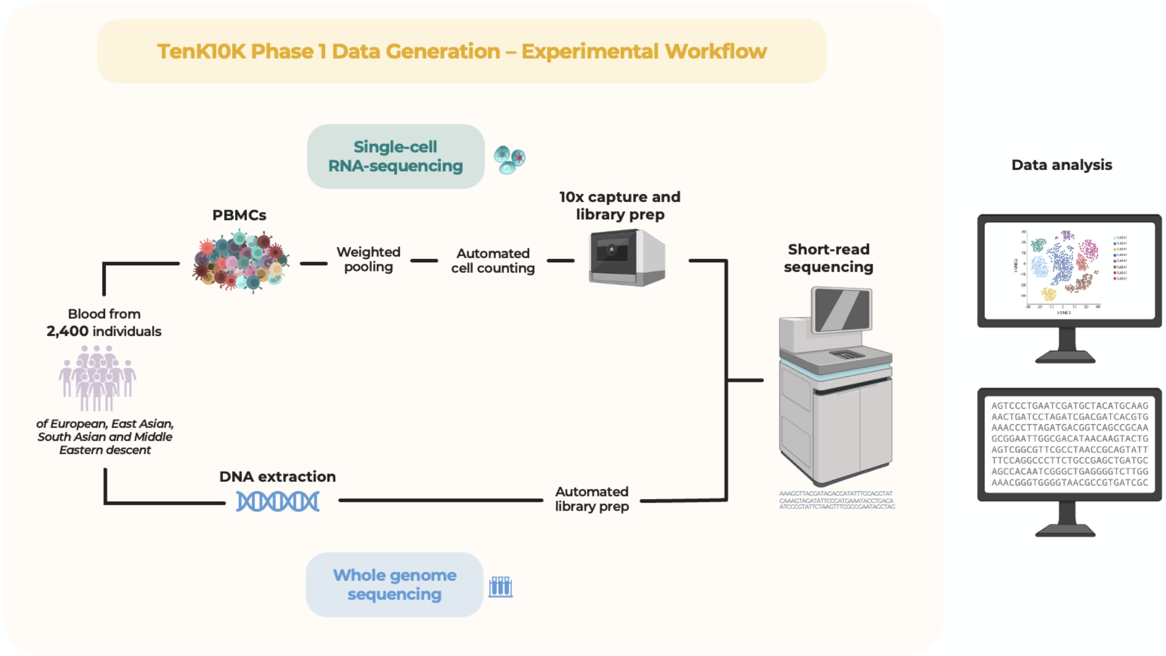


**Supplementary Figure 49. Experimental workflow.** For scRNA-seq, cryopreserved PBMC were thawed, counted and multiplexed (weighted by live cell content) into pools of 8-14 donors. To maximise throughput, these pools were cryopreserved and thawed once more, before batch captures were performed on the ChromiumX analyser. WGS libraries were generated from matched extracted DNA. Both scRNA-seq libraries and WGS libraries were sequenced on the NovaSeq6000 platform. More detail can be found in Methods.

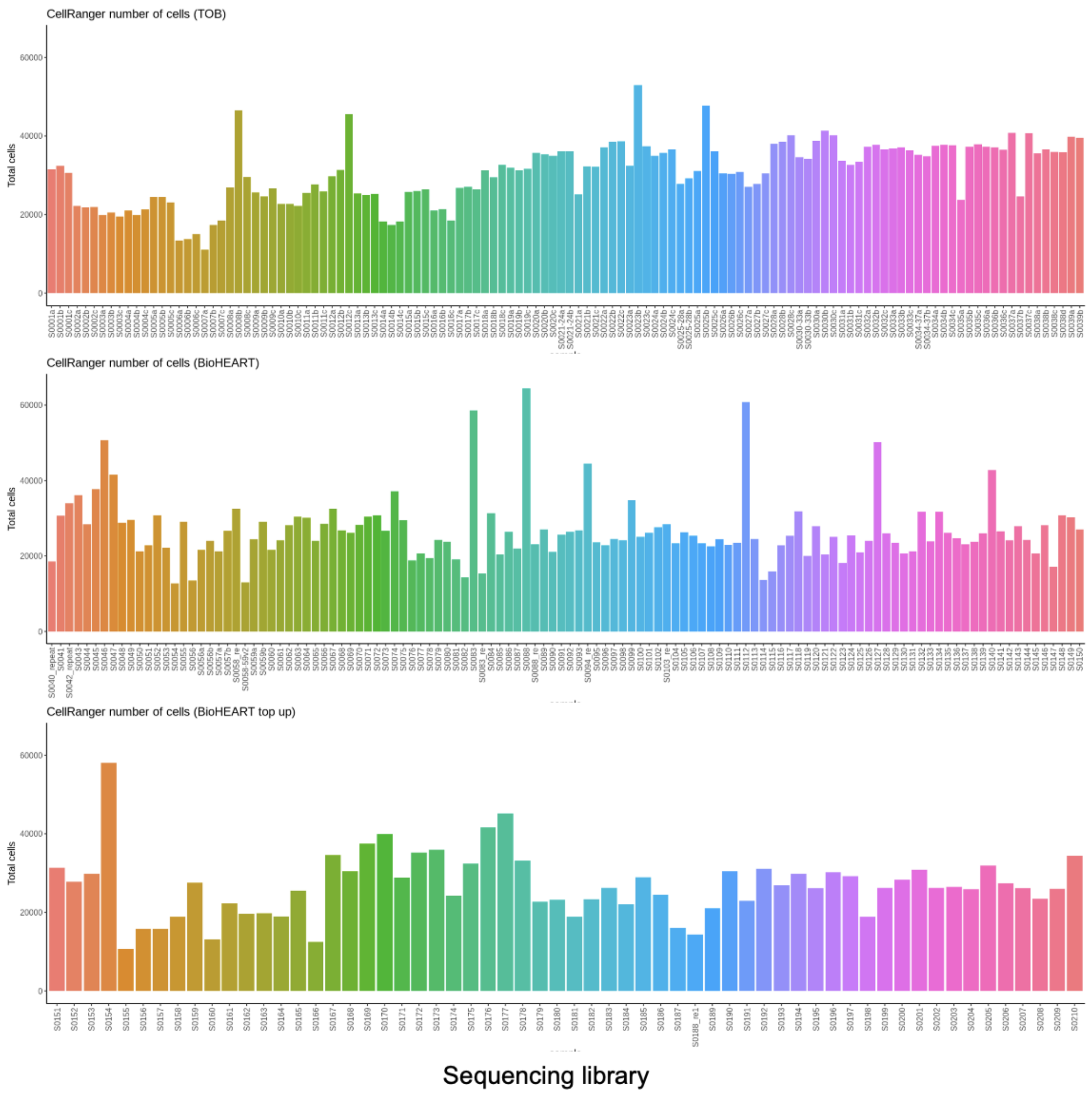


**Supplementary Figure 50. Number of cells identified by CellRanger.** Each bar represents one sequencing library.

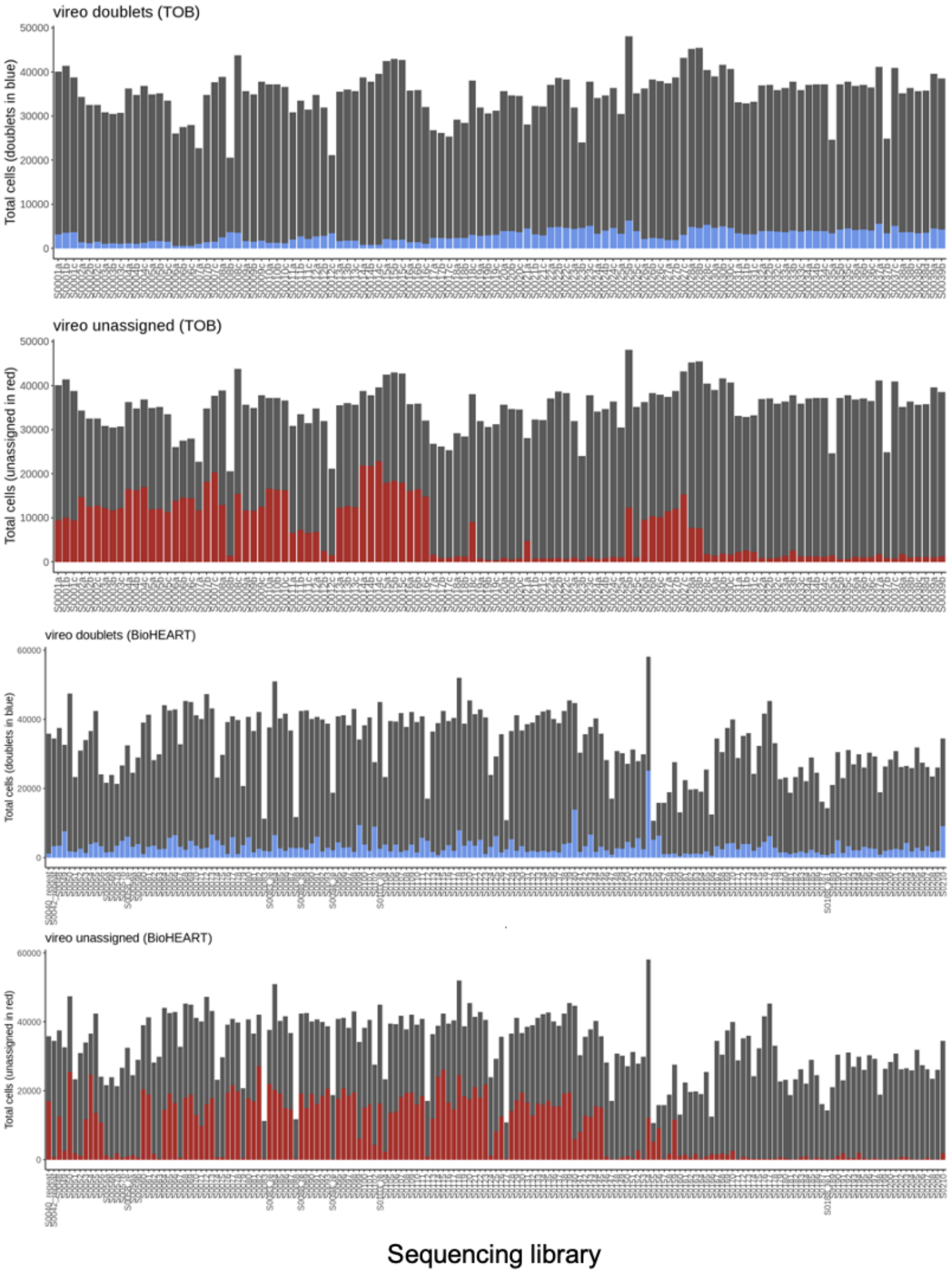


**Supplementary Figure 51. Number of cells identified as doublets or unassigned.** Out of the total number of cells identified; each bar represents one sequencing library.

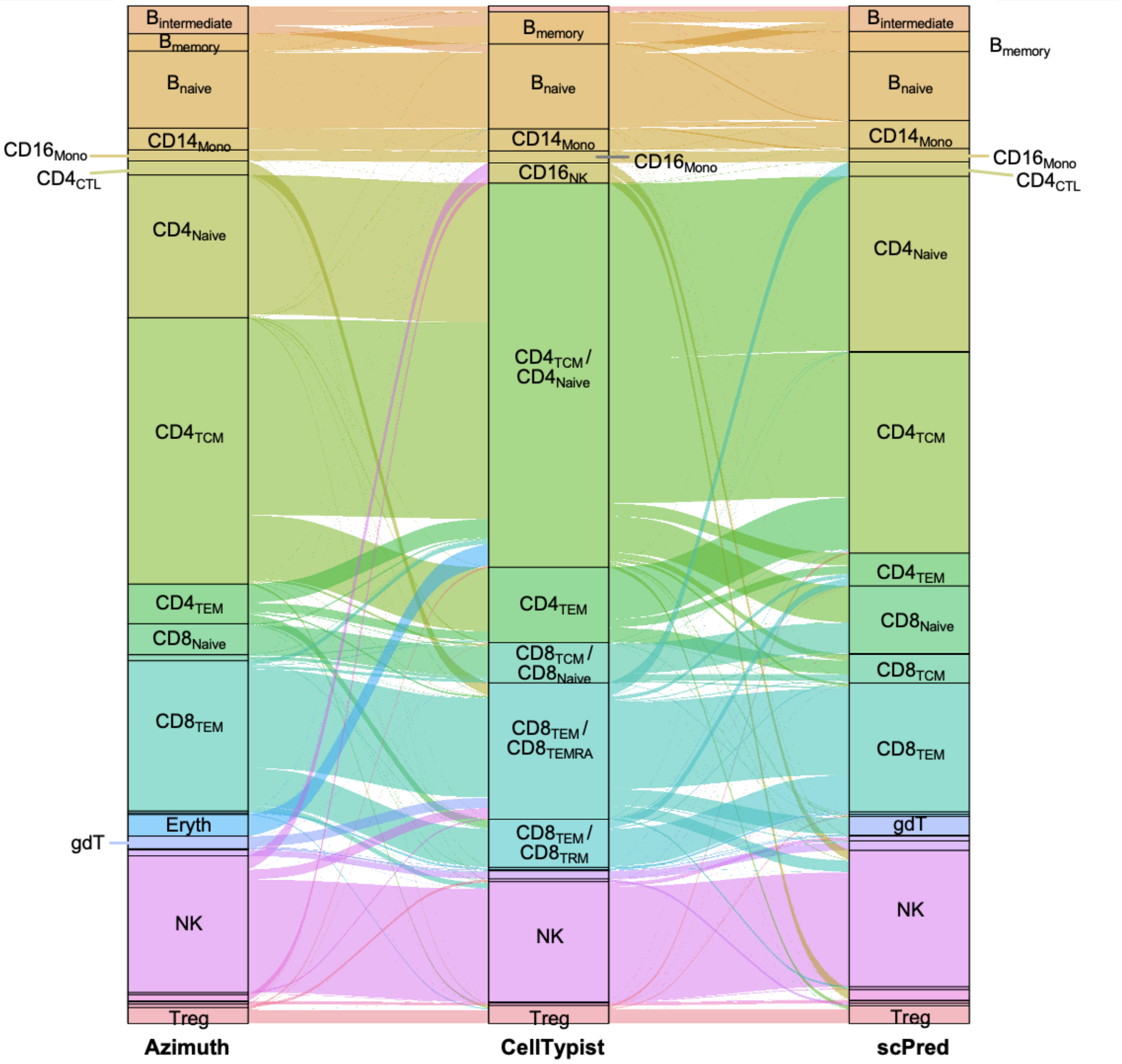


**Supplementary Figure 52. Cell typing concordance between methods.** For a representative sequencing library, agreement between three cell typing methods: scPred, Azimuth and Celltypist (only the more abundant cell types are labelled for clarity).

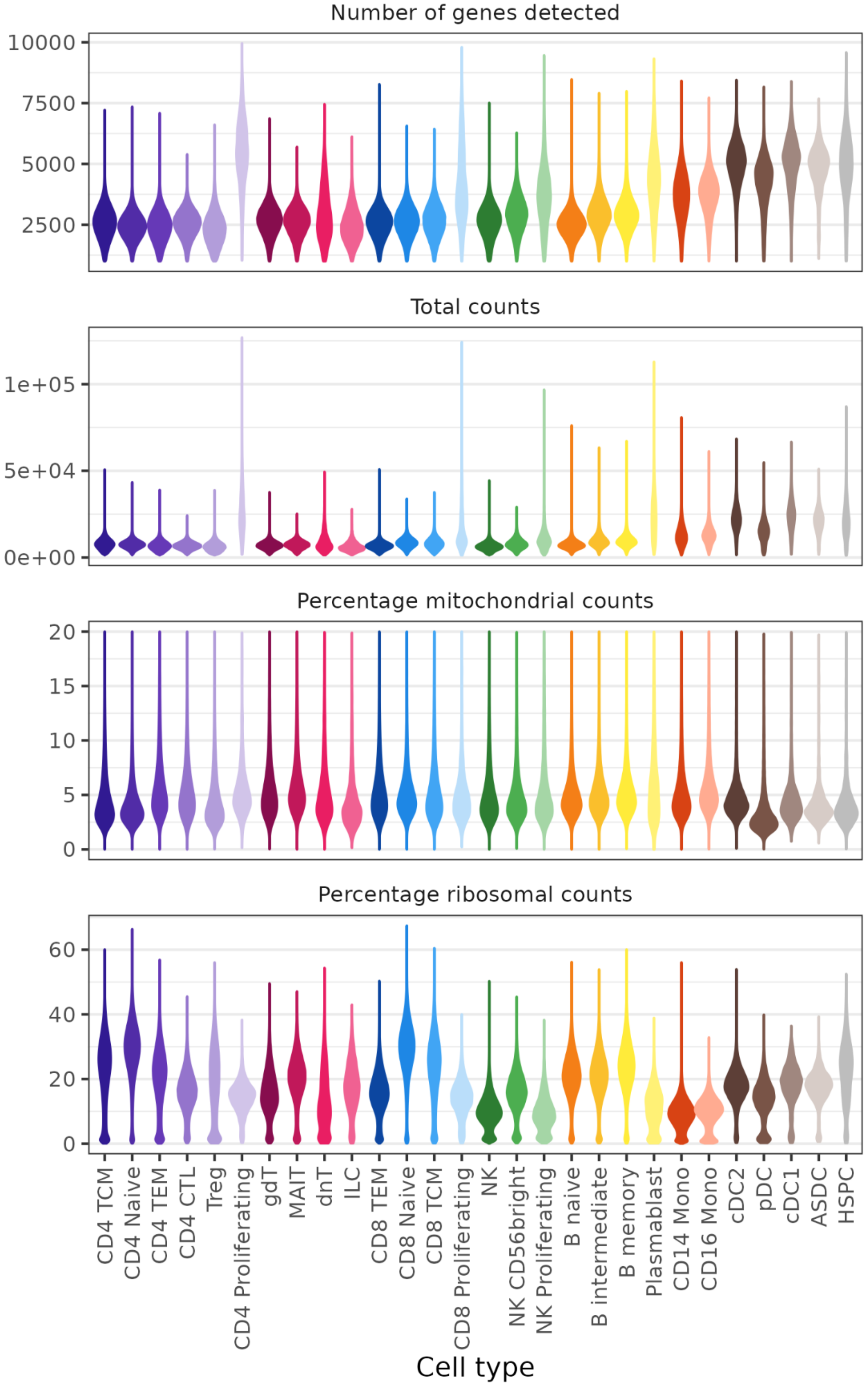


**Supplementary Figure 53. Cell QC metric distributions.** For each cell type.

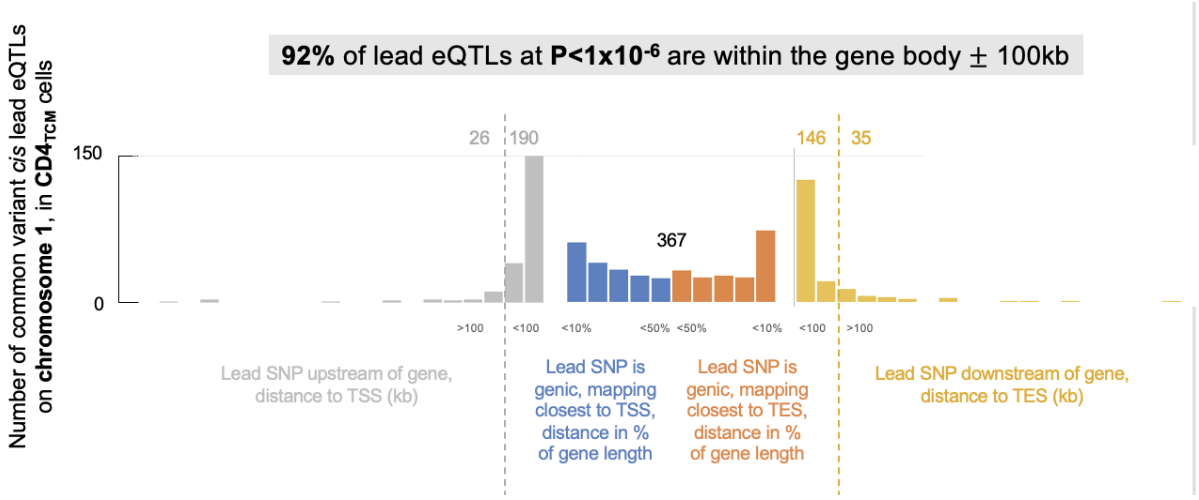


**Supplementary Figure 54. Window size comparison 100kb vs 1Mb.** Location of lead common variant eQTLs with respect to the gene body (similar to Vosa *et al*, 2021[2]). When considering variants with *p*-value<$10^{-6}$, 92% are found within the +/-100kb window size around the gene body.

### Supplementary Tables

*All supplementary tables are provided as supplementary files.*

### Supplementary Note

**Supplementary Note 1. Monocyte proportion differences across cohorts.**

Monocyte proportion differences were noted between TOB and BioHeart cohorts: together CD14+ and CD16+ monocytes made up approximately 9.4% of all BioHEART cells (*n*=184,639 and 68,605 for the two subtypes respectively, out of a total of 2,690,210 cells), and only 2.4% of all TOB cells (*n*=41,384 and 24,326 out of 2,748,469). This is likely to be an artefact of different sample handling between batches, rather than true biological variation. Monocyte proportions are demonstrated to change dynamically with long-term cryopreservation[12].

Processing differed for cryopreserved PBMC in each cohort. Samples from the TOB cohort were collected prior to 2018 and had one previous freeze-thaw cycle before thawing again in 2023 for TenK10K scRNA-seq. BioHEART PBMC samples were collected from 2015 to 2023 and thawed for the first time prior to pooling.

We also compared the monocyte proportion between scRNA-seq and scATAC-seq and the monocyte proportion from scATAC-seq are more consistent with the public PBMC datasets[13]. Given the scATAC-seq data were processed from samples more recently collected, it further supports the hypothesis that the monocytes proportion differences are due to sample handling rather than biological variations.

**Supplementary Note 2. Cellbender tool limitations with high-throughput scRNA-seq.**

We originally ran CellBender[14] (v0.3.0) to measure ambient RNA in our data. However, the estimates of cell numbers were not well aligned with those estimated by Cellranger (especially for the TOB cohort), likely because of the new high-throughput kit we used, as shown in Supplementary Fig. 55.

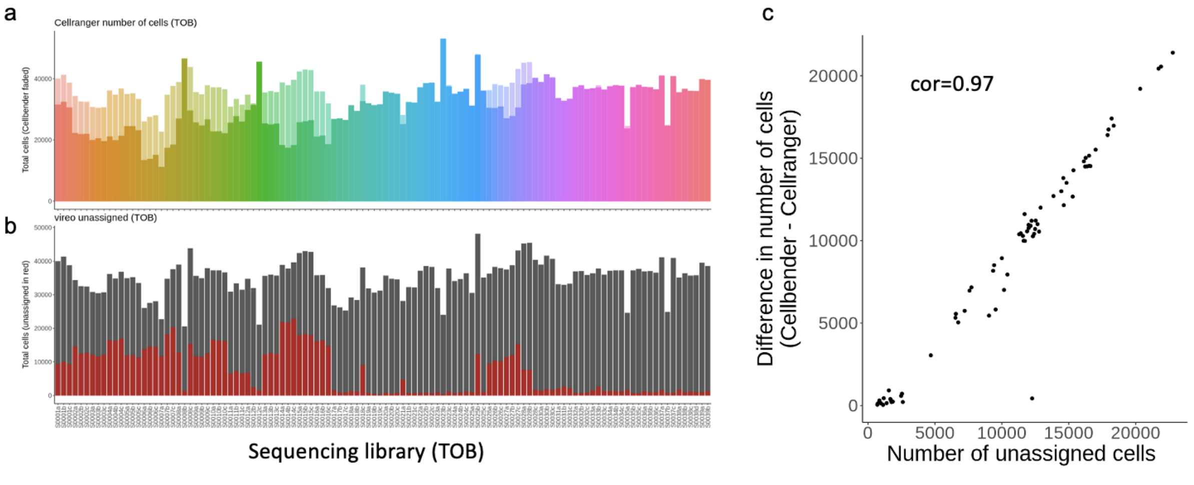


**Supplementary Figure 55. Cellbender-Cellranger mismatch in the estimated number of cells.** **a)** Cellbender number of cells in faded colour and Cellranger number of cells in solid colour for a few sequencing libraries, showing Cellbender often overestimates the number of cells by as much as 2-fold. **b)** Similar to the above but now stratified by whether the droplets were assigned to an individual (by vireo, in grey) or deemed "unassigned" (could not confidently be assigned to any donors, in red) when using all droplets called as cells by Cellbender, showing how the extra cells are likely not real as they cannot be assigned to any donor. **c)** Correlation between the difference in number of cells between Cellbender and Cellranger (y-axis) and the number of unassigned cells (x-axis), excluding *n*=3 point for which the estimate in number of cells was higher in Cellranger

We still measured the ambient RNA %, but did not remove any cells as we could not be sure the estimates were robust given the mismatch outlined above. Supplementary Fig. 56 shows the cellbender background fraction was <80% for all QC-ed cells, and only 2.1% of cells had Cellbender background fraction > 20%.

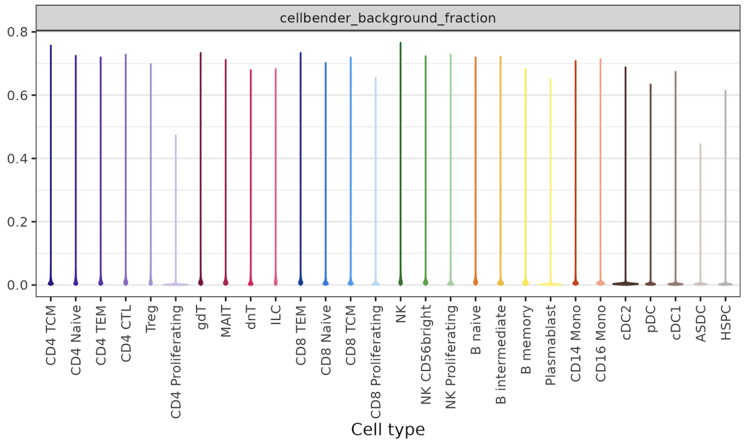


**Supplementary Figure 56. Cellbender background fraction distributions.**
Violin plots for each cell type, coloured by cell type.

**Supplementary Note 3. Convergence fails of SAIGE-QTL.**
For a subset of gene-cell type combinations (0.27%) the SAIGE-QTL[15] model (fitting the null) failed to converge and these were excluded from the analysis. These convergence failures were likely due to numerical errors due to the covariate matrix structure. A solution we found to reduce the number of convergence errors was to change the 'isCovariateOffset' parameter to TRUE, which regresses out the covariates from the phenotype before running the model. However, this setting results in slightly inflated *p*-values, thus was not a working solution for the main results. We still provide summary statistics for this run in Zenodo as a separate file. Supplementary Fig. 56 shows the results when setting 'isCovariateOffset=TRUE' for *ITGA4* in CD14+ monocytes. The QQ plot shows some inflation in the permuted control, which we see consistently using this setting.

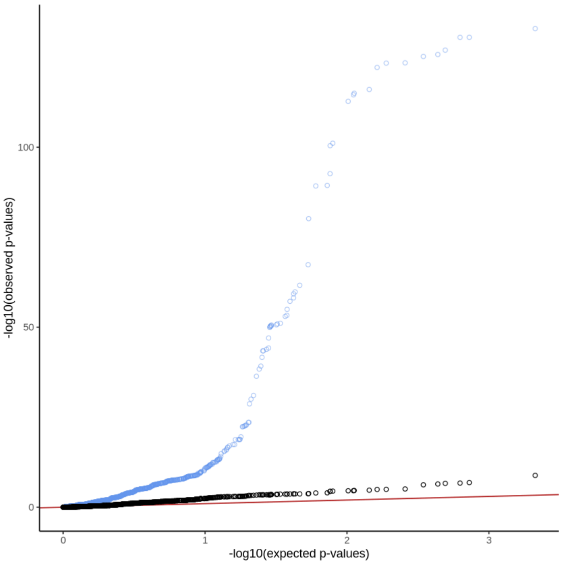


**Supplementary Figure 57. Inflated *p*-values when regressing out covariates.**
As demonstrated testing all common *cis* variants for *ITGA4* (ENSG00000115232) in CD14+ monocytes.
QQ plot of *p*-values, blue: true association and black: permuted control

**Supplementary Note 4. ACAT-V is not calibrated in our data.**

SAIGE-QTL[15]'s set-test implementation includes not only the burden and SKAT tests throughout this manuscript, but also the ACAT-V test[16]. However, in our results we noticed some inflation in the *p*-values when shuffling or individuals which we did not observe for the other two tests (Supplementary Fig. 58). As a consequence, we do not present these results and when reporting a single combined *p*-value we use the Cauchy aggregation *p*-values for burden and SKAT only.

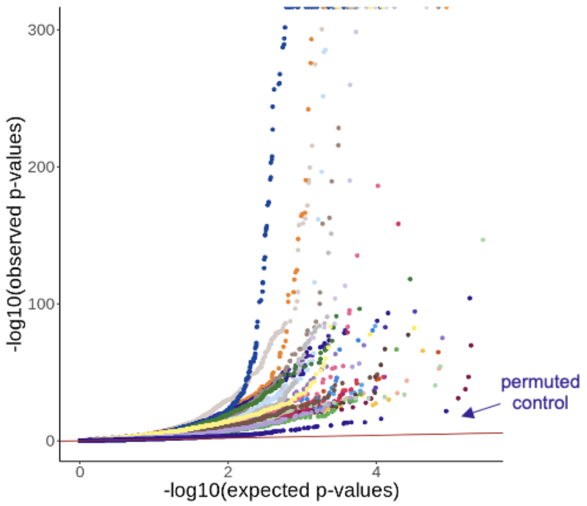


**Supplementary Figure 58. Inflated *p*-values for ACAT-V results.**

QQ plots of ACAT-V *p*-values, across all cell types included the indicated permuted control ($CD4_{TCM}$ after shuffling individual IDs). Similar to Fig. 2a but this time showing some inflation.

**Supplementary Note 5. SuSie limitations with SAIGE-QTL outputs.**

SuSie[2] uses a linear assumption, which is violated by the Poisson model used in SAIGE-QTL[15]. This leads to some inconsistencies, for example in the number of primary eQTLs identified using the original method (while qualitatively retaining the same rank across cell types, Supplementary Fig. 59).

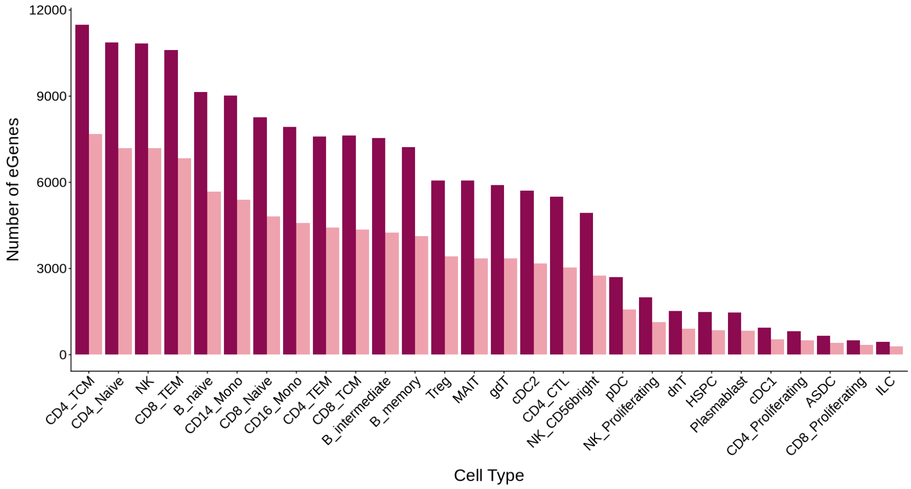


**Supplementary Figure 59. Fewer eGenes identified by SuSie.**

Bar plots showing the number of eGenes identified using the main approach used here (FDR<5% after ACAT gene-level *p*-value correction) compared to the equivalent number using SuSie (number of eGenes with at least one 95% credible set). While the overall trend of numbers of eGenes across cell types is similar, the FDR approach identifies on average 1.68 more eGenes across cell types.
